# Pleiotropic genetic architecture linking schizophrenia and substance use disorders

**DOI:** 10.64898/2026.08.31.26361799

**Authors:** Selena Aranda, Dora Koller, Sergi Papiol, María Soler Artigas, Ana M Pérez-Gutiérrez, Javier González-Peñas, Monika Budde, Pablo Jácome-Ferrer, Celso Arango, Kristina Adorjan, Elisabet Vilella, Gerard Muntané, Lourdes Martorell, Maria Heilbronner, M. Dolores Moltó, Olga Rivero, Alba Navarro-Flores, Julio Bobes, Motjaba Oraki Kohshour, Benedicto Crespo-Facorro, Daniela Reich-Erkelenz, Ana González-Pinto, Eva C Schulte, Manuel Arrojo, Gerardo Flórez, Fanny Senner, Ion-George Anghelescu, Volker Arolt, Detlef E Dietrich, Andreas J Fallgatter, Christian Figge, Markus Jäger, Fabian U Lang, Georg Juckel, Carsten Konrad, Jens Reimer, Eva Z Reininghaus, Max Schmauß, Andrea Schmitt, Carsten Spitzer, Jens Wiltfang, Jörg Zimmermann, Lourdes Fañanás, Urs Heilbronner, Bárbara Arias, Peter Falkai, Javier Costas, Thomas G Schulze, Marina Mitjans, Bru Cormand

**Author notes:** **Corresponding authors:** Bru Cormand and Marina Mitjans. Department of Genetics, Microbiology and Statistics, Faculty of Biology, Universitat de Barcelona, Av. Diagonal 643, Prevosti Building, 3rd floor, 08028 Barcelona, Spain. and. Equally contributed (Selena Aranda, Dora Koller). Equally supervised (Marina Mitjans, Bru Cormand).

## Abstract

Schizophrenia (SCZ) frequently co-occurs with substance use disorders (SUDs), yet the genetic basis of this comorbidity remains unclear. Using the latest European-ancestry genome-wide association studies (GWAS) for SCZ, cannabis use disorder (CanUD), opioid use disorder (OUD), problematic alcohol use (PAU), tobacco use disorder (TUD), and a general addiction factor (AF), together with two SCZ and one SUD case-control samples with individual-level genotype data, we applied multiple complementary genomic approaches to characterize their shared genetic architecture. Significant positive genome-wide genetic correlations were observed across all SCZ–SUD pairs. Local genetic correlation analyses identified multiple genomic regions contributing to this shared architecture, with both positive and negative correlations, and evidence of genomic regions shared across multiple SCZ–SUD pairs. Polygenic overlap analyses indicated substantial sharing (25-50%) of trait-associated variants between SCZ and SUDs. Genomic structural equation modelling supported a common latent factor underlying SCZ and all SUDs, accounting for approximately 23% of SCZ variance. Cross-trait polygenic risk score (PRS) analyses showed bidirectional associations between SCZ and SUD genetic liability. Mendelian randomization analyses provided evidence for a bidirectional causal relationship between SCZ and CanUD. Horizontal pleiotropy analyses identified numerous loci with concordant and discordant effects across traits, including loci shared among multiple SCZ–SUD pairs. Gene mapping and enrichment analyses indicated pathways related to neuroplasticity, synaptic transmission, immune system, metabolism and proteolysis, including both shared and SCZ–SUD-specific biological processes. Overall, these findings suggest that part of SCZ liability reflects genetic susceptibility to SUDs with potential implications for patient stratification and clinical management.

## Introduction

Schizophrenia (SCZ) is a complex psychiatric disorder affecting approximately 1% of the population [1]. Both genetic and environmental factors contribute to its aetiology [2]. The genetic architecture of SCZ is highly complex and heterogeneous. Twin-based heritability (h^2^) for SCZ has been estimated at 60-80% [2], while single nucleotide polymorphisms (SNPs) collectively explain 24% of the variance, according to the most recent genome-wide association study (GWAS) [3].

Individuals with SCZ are highly vulnerable to developing substance use disorders (SUDs) [4,5]. SUDs are complex heritable conditions (twin-based h^2^ ∼ 30-80%; SNP-based h^2^ ∼ 6.6-11.8%) characterized by impaired control over the use of psychoactive substances despite their adverse consequences [6]. The lifetime prevalence of SUDs among individuals with SCZ has been reported to reach up to 43.1%–65.0% for alcohol use disorder [7,8], 50.8% for cannabis use disorder (CanUD) [8], 28.5% for tobacco use disorder (TUD) [9], and 13.0% for opioid use disorder (OUD) [10]. These rates exceed those observed in non-SCZ controls. Importantly, the co-occurrence of SUDs in individuals with SCZ is associated with worse clinical outcomes, cognitive impairment, reduced global functioning, violence, poor treatment adherence, and increased rates of rehospitalization and mortality [11–13].

Multiple hypotheses have been proposed to explain the high co-occurrence and clinical burden of SCZ and SUDs, yet no definitive conclusion has been reached [13]. From a genetic perspective, several studies using conjunctional false discovery rate (conjFDR) analyses have identified pleiotropic loci jointly associated with SCZ and substance use-related traits. For example, Wiström et al. identified 52 pleiotropic loci shared between SCZ and alcohol use, of which 19 were replicated in an independent sample [14]. Similarly, a cross-trait genomic analysis of SCZ, CanUD, and tobacco smoking identified 202 genomic loci with convergent effects across all three traits, as well as 37 loci jointly associated with SCZ and CanUD and 46 jointly associated with SCZ and tobacco smoking [15]. In addition, Hole et al. identified two shared genomic risk loci for SCZ and OUD [16]. Beyond shared loci, recent studies investigating the relationship between SCZ and cannabis-related phenotypes have reported a causal effect of genetic liability to SCZ on CanUD [17] and vice versa [18]. These studies also reported a positive genome-wide genetic correlation between the two phenotypes [17,18], although local analyses revealed a mixture of positively and negatively correlated genomic regions [17,18]. More recently, a cross-disorder genomic analysis encompassing fourteen disorders, including SCZ and SUDs, characterized broad common genetic mechanisms underlying their co-occurrence [19].

Despite these advances, further analyses using complementary genomic approaches in increasingly powered GWAS are needed to refine our understanding of the shared genetic architecture of SCZ and SUDs. Such insights may inform disease sub-classification, clinical trajectories, and treatment strategies. In the present study, we applied a comprehensive set of genomic approaches, including multivariate modelling, polygenic overlap analyses, polygenic risk scoring, Mendelian randomization and pleiotropy mapping to characterize the genetic relationships between SCZ and multiple SUDs. We also investigated the biological pathways underlying pleiotropic loci and examined the convergence of annotated genes across SCZ-SUD pairs to identify shared biological mechanisms.

## Material and methods

### GWAS summary statistics

We used the most recent European-ancestry GWAS meta-analyses for SCZ [3], CanUD [20], PAU [21], TUD [22], and OUD [23] (Table 1). Clinical diagnoses in these studies were derived from DSM (Diagnostic and Statistical Manual of Mental Disorders) and ICD (International Classification of Diseases) criteria and self-reported information. For PAU, cases were defined based on alcohol use disorder, alcohol dependence, and items assessing problematic alcohol-related consequences using the Alcohol Use Disorders Identification Test (AUDIT) [21].

**Table 1.** GWAS summary statistics and individual-level genetic datasets used in the present study.

| Dataset | Diagnosis | N<sub>cases</sub> | N<sub>controls</sub> | N<sub>effective</sub> | SNP-h<sup>2</sup> (%) | Associated loci | Ethnicity | Reference |
| --- | --- | --- | --- | --- | --- | --- | --- | --- |
| GWAS summary statistics |  |  |  |  |  |  |  |  |
| SCZ GWAS | SCZ | 53,386 | 77,258 | 58,749 | 24.0 | 287 | European | Trubetskoy et al., 2022 |
| CanUD GWAS | CanUD | 42,281 | 843,744 | 161,053 | 6.7 | 22 | European | Levey et al., 2023 |
| ODU GWAS | ODU | 15,040 | 282,607 | 57,120 | 10.7 | 14 | European | Kember et al., 2022 |
| PAU GWAS | PAU | 113,325 | 639,923 | 385,102 | 6.6 | 110 | European | Zhou et al., 2023 |
| TUD GWAS | TUD | 174,021 | 565,874 | 532,367 | 11.7 | 88 | European | Toikumo et al., 2023 |
| AF gSEM* | PAU, PTU, CanUD, OUD | 1,025,550 | - | - | - | 19 | European | Hatoum et al., 2023 |
| Individual-level genetic datasets |  |  |  |  |  |  |  |  |
| CIBERSAM sample | SCZ | 1,826 | 1,372 | - | - | - | European (Spain) | Mitjans et al., 2024 |
| PsyCourse Study | SCZ | 553 | 404 | - | - | - | European (Germany) | Budde et al., 2019 |
| Galician sample | SUD** | 1,176 | 1,001 | - | - | - | European (Spain) | Gurriarán et al., 2019 |
| AF: Addition Factor; CanUD: Cannabis use disorder; gSEM: Genomic structural equation model; GWAS: Genome-wide association study; h<sup>2</sup>: heritability; OUD: Opioid use disorder; PAU: Problematic alcohol use; PTU: Problematic tobacco use; SCZ: Schizophrenia; SUD: Substance use disorder; TUD: Tobacco use disorder. |  |  |  |  |  |  |  |  |
| *Summary statistics from a multivariate genome-wide association meta-analysis integrating published European GWAS of PAU, problematic tobacco use, CanUD and OUD. |  |  |  |  |  |  |  |  |
| **Substances considered were alcohol, tobacco, cannabis, cocaine, opiates, hypnotics, stimulants and hallucinogens. |  |  |  |  |  |  |  |  |

We also included summary statistics from a multivariate GWAS meta-analysis of the addiction factor (AF), which integrates published European-ancestry GWAS of PAU, problematic tobacco use, CanUD and OUD. This analysis partly overlaps with the GWAS datasets analysed in the present study, as it incorporates earlier versions of these datasets, and identified 19 genome-wide significant loci associated with general addiction liability [24] (Table 1).

### Schizophrenia samples

Clinical, sociodemographic and genotypic data from unrelated individuals with European ancestry were obtained from two independent multicenter SCZ samples: a Spanish sample recruited through the Spanish Mental Health Networking Biomedical Research Centre (CIBERSAM) and a German sample belonging to the the PsyCourse Study (Table 1). Detailed genotyping procedures and quality control are described in the Supplementary Methods.

The CIBERSAM sample (N_cases_ = 1,826; N_controls_ = 1,372) was recruited by eight research groups across Spain [25]. Diagnoses were based on DSM-IV criteria for SCZ spectrum disorders. The transdiagnostic PsyCourse Study was recruited through a network of 20 centres in Germany and Austria [26]. For the present study, participants with a diagnosis of SCZ spectrum disorders and healthy controls were selected (N_cases_ = 553; N_controls_ = 404). Diagnoses were established according to DSM-IV or ICD-10 diagnostic criteria. Written informed consent was obtained from all participants in both samples, and the respective studies were approved by the local ethics committees of the participating centres.

### Substance use disorder sample

We retrieved clinical, sociodemographic and genotypic data from 2,177 unrelated individuals with European ancestry (1,176 substance abuse/dependence patients and 1,001 controls) from Galicia, Spain (Table 1). Detailed genotyping procedures and quality control are described in the Supplementary Methods.

Patients were recruited at several Addictive Disorders Assistance Units from Galicia, and the sample was enriched for polydrug users. Diagnoses were based on ICD10 criteria for dependence of a substance and abuse/dependence of another substance. The substances considered were alcohol (F10.1, F10.2; N=779), tobacco (F11.1, F11.2; N=1,009), cannabis (F12.1, F12.2; N=606), cocaine (F13.1, F13.2; N=672), opiates (F14.1, F14.2; N=571), hypnotics (F15.1, F15.2; N=195), stimulants (F16.1, F16.2; N=76) and hallucinogens (F17.1, F17.2; N=41). Because many patients presented abuse/dependence on multiple substances, these categories are not mutually exclusive and the sum of cases across substances exceeds the total number of patients. Controls were recruited from blood donors at Santiago de Compostela, Galicia, and those who self-reported chronic alcoholism and use of illicit drugs were excluded from the sample. Substance use was not assessed in the control group; therefore, some controls may exhibit SUD symptoms. All participants provided written informed consent to take part in the study.

### Genetic correlation analyses

Genome-wide genetic correlations among SCZ, the different SUDs, and AF were estimated using linkage disequilibrium score regression (LDSC) [27]. Linkage disequilibrium (LD) estimates were obtained from the Phase 3 European reference panel of the 1000 Genomes Project [28]. To ensure inclusion of well-imputed, common variants, LDSC analyses were restricted to SNPs present in the HapMap 3 reference panel [29], and the highly polymorphic major histocompatibility complex (MHC) region (chr6: 26,000,000–34,000,000, GRCh37/hg19) was excluded.

Local genetic correlations between SCZ and each SUD and AF were assessed using Local Analyses of [co]Variant Association (LAVA) across a predefined set of 2,495 genomic regions [30]. Only regions displaying significant heritability were retained for downstream analyses. Genes within significantly correlated regions were identified using positional mapping implemented in the topR R package [31].

### Polygenic overlap

Polygenic overlap between SCZ and each SUD was estimated using bivariate causal mixture models implemented in MiXeR [32]. The extend of polygenic overlap was summarized using the Dice coefficient (DC), which represents the proportion of shared causal variants relative to the total number of variants with nonzero effects across both traits [32]. Genome-wide genetic correlations (r_g_) and genetic correlation within the shared polygenic component (r_gs_) were also calculated. Variants located within the MHC region (chr6: 26,000,000–34,000,000, GRCh37/hg19) were excluded.

Conditional quantile–quantile (Q–Q) plots were generated to visualise polygenic enrichment across traits, and log-likelihood surfaces were inspected to assess parameter identifiability and convergence of the optimization procedure. Model fit was further assessed by comparing the Akaike Information Criterion (AIC) of the best-fitting bivariate MiXeR model against reference models representing minimum and maximum possible polygenic overlap. Differences in AIC were used to evaluate whether the best-fitting solution provided a superior fit relative to these boundary scenarios, with positive AIC values indicating that the best-fitting MiXeR estimates were statistically distinguishable from the corresponding reference models.

### Genomic structural equation modelling

Shared genetic architecture between SCZ and SUDs was further investigated using Genomic Structural Equation Modelling (gSEM) [33]. The number of latent factors was first evaluated using the Kaiser criterion and exploratory factor analysis (EFA) implemented in the psych R package [34]. Model convergence and factor loadings were then assessed through a confirmatory factor analysis (CFA) using the lavaan R package [35]. Model fit for both EFA and CFA was evaluated using standard fit indices provided by the GenomicSEM R package [33]. All analyses were restricted to HapMap 3 variants, and LD estimates were derived from the Phase 3 European populations of the 1000 Genomes Project [28].

### Polygenic risk scoring

We evaluated the association between CanUD-, OUD-, PAU-, TUD-, and AF-derived polygenic risk scores (PRSs) and SCZ diagnosis in the CIBERSAM and the PsyCourse samples. The association between SCZ-derived PRS and SUD diagnosis was evaluated in the Galician sample. Posterior SNP effect sizes were inferred under continuous shrinkage priors from each GWAS and AF meta-analysis using the 1000 Genomes European LD reference panel [28]. The global shrinkage parameter (φ) was estimated with PRS-CS [36], and posterior SNP effect size estimates were used to compute the corresponding PRSs in the target samples with PLINK [37]. Associations were tested using logistic regression models, including sex, the first ten principal components (PCs), and the recruitment centre as covariates. For each PRS, a baseline logistic regression model including only covariates and a full model including the PRS and covariates were fitted. The variance explained by the PRS was estimated as the increase in Nagelkerke’s pseudo-R² using the rcompanion R Package [38]. P-values were adjusted for multiple testing using the Benjamini–Hochberg false discovery rate (FDR) procedure, and associations with FDR-adjusted p-values < 0.05 were considered statistically significant.

### Genetically informed causal inference analysis

Bidirectional two-sample Mendelian randomization (MR) analyses were conducted using the TwoSampleMR [39] and MR-PRESSO [40] R packages to investigate potential causal genetic relationship between SCZ and SUDs. The multiplicative random-effects inverse variance weighted (IVW) method was used as the primary approach to infer causality [41]. When evidence of a potential causal relationship was detected (IVW p-adjusted < 0.05), additional sensitivity analyses were performed, including weighted median [42], weighted mode [43], MR-PRESSO global test [40] and Steiger filtering [44].

Additionally, the I^2^ statistic was calculated to quantify the strength of the violation of the NO Measurement Error (NOME) assumption [45]. In cases where the NOME assumption was not violated (I^2^ > 0.9), the presence of pleiotropic instruments was tested using the MR-Egger intercept, and the MR-Egger sensitivity analyses was also taken into consideration when there was evidence of pleiotropy (p-value of MR-Egger intercept < 0.05) [46]. When the deviation from the NONE assumption was weak (0.6 < I^2^ < 0.9), we evaluated the presence of pleiotropic instruments using the simulation extrapolation (SIMEX) intercept, and the SIMEX sensitivity analysis was also performed if there was evidence of pleiotropy (p-value of SIMEX intercept < 0.05) [45]. In cases of strong deviation from the NONE assumption (0.6 > I^2^), data was considered to be unpowered to detect causality.

### Horizontal pleiotropy analyses

Genomic loci underlying horizontal pleiotropy between SCZ and each SUD or AF were identified using the PolarMorphism R package [47]. SNPs with a theta q-value < 0.05 were considered pleiotropic and subsequently clumped to remove variants in high LD (R^2^ > 0.9) within a 5 Mb window using PLINK [37]. Subsequently, we evaluated whether the identified pleiotropic loci or variants in high LD (R^2^ > 0.9) had been previously reported as pleiotropic between SCZ and SUDs, including alcohol use [14], CanUD [15], OUD [16], and tobacco smoking [15].

### Biological characterization of pleiotropic variants shared between schizophrenia and substance use disorders

Pleiotropic loci identified by PolarMorphism for each SCZ-SUD pair were annotated to coding genes using positional mapping and brain expression quantitative trait loci (eQTL) mapping implemented in the Functional Mapping and Annotation (FUMA) tool [48]. The annotated genes were then subjected to non-redundant Gene Ontology (GO) enrichment analyses of biological processes using the clusterProfiler R package [49].

To identify convergent biological pathways across SCZ-SUD pairs, we examined the overlap of genes annotated by FUMA for each SCZ–SUD comparison. Genes shared across multiple SCZ– SUD pairs were grouped according to their overlap pattern and GO enrichment analyses of biological processes were subsequently performed for each shared gene set using the clusterProfiler R package [49].

## Results

### Genetic correlation analyses

Positive genome-wide genetic correlations were observed between SCZ and all examined SUDs and AF, with estimates ranging from 0.10 ± 0.02 for TUD (p = 5.34×10^−6^) to 0.37 ± 0.02 for CanUD (p = 1.38×10^−60^) (Figure 1A, Table S1), indicating a moderate but consistent positive genome-wide genetic correlation between SCZ and SUDs.

**Figure 1.**
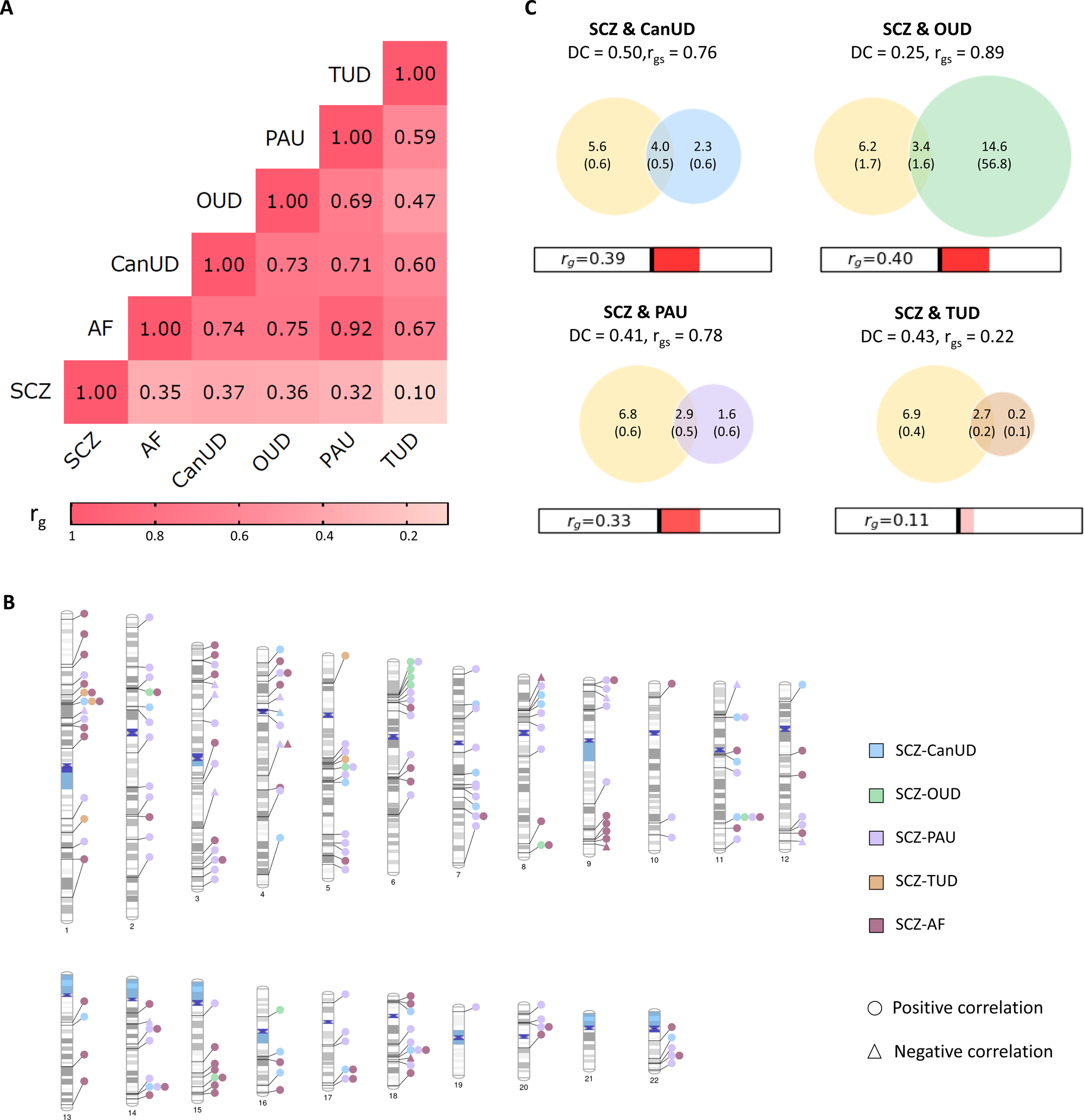
Genetic correlations and polygenic overlap between schizophrenia and substance use disorders. (A) Genome-wide genetic correlations among schizophrenia and substance use disorders estimated by LDSC. (B) Genomic regions locally correlated between schizophrenia and substance use disorders according to LAVA. (C) Polygenic overlap and genome-wide genetic correlation between schizophrenia and substance use disorders estimated by MiXeR. The numbers within the Venn diagrams indicate the estimated quantity of causal variants (in thousands) per component, followed by the standard error. The size of the circles reflects the degree of polygenicity. AF: Addiction factor; CanUD: Cannabis use disorder; DC: Dice coefficient; OUD: Opioid use disorders; PAU: Problematic alcohol use; r_g_: Genetic correlation; rgs: Genetic correlation of the shared component; TUD: Tobacco use disorder; SCZ: Schizophrenia.

Local genetic correlation analyses further identified specific genomic regions contributing to this shared architecture. Significant local genetic correlations were detected for SCZ-AF (N_regions_ = 69; 65 positive, 4 negative), SCZ-CanUD (N_regions_ = 23; 22 positive, 1 negative), SCZ-OUD (N_regions_ = 10; all positive), SCZ-PAU (N_regions_ = 102; 92 positive, 10 negative), and SCZ-TUD (N_regions_ = 5; all positive) (Figure 1B, Table S2). Notably, a subset of these regions was shared across multiple cross-trait pairs (Figure 1B), indicating convergent genomic loci underlying the shared architecture.

### Polygenic overlap

MiXeR analysis revealed moderate polygenic overlap between SCZ and all examined SUDs. Estimated Dice coefficients (DC) ranged from 0.25 to 0.50, with the largest overlap observed for SCZ-CanUD (DC = 0.50; ∼4,000 shared variants), followed by SCZ-TUD (DC = 0.43; ∼2,700 variants), SCZ-PAU (DC = 0.41; ∼2,900 variants), and SCZ-OUD (DC = 0.25; ∼3,400 variants) (Figure 1C). Genome-wide genetic correlations (r_g_) estimates were consistent with those obtained using LDSC. In all cases, the genetic correlation within the shared polygenic component (r_gs_) was higher than the genome-wide one (Figure 1C).

Evidence for polygenic enrichment was further supported by the successive leftward deviations from the null line observed in the conditional Q-Q plots between SCZ and CanUD, PAU, and TUD, and vice versa, indicating increasing enrichment of association signals as a function of cross-trait association strength (Figure S1).

The robustness of overlap estimates was evaluated using log-likelihood profiles (Figure S1) and AIC comparisons against minimum- and maximum-overlap reference models (Table S3). PAU showed the strongest support for an intermediate overlap model, with a well-defined likelihood maximum and positive AIC values relative to both minimum and maximum boundaries. CanUD also displayed a clear likelihood peak, although AIC was near the lower-overlap boundary, indicating a low overlap estimate. In contrast, OUD and TUD exhibited likelihood profiles constrained to the boundaries of the parameter space, without a well-defined internal maximum, suggesting reduced identifiability of the overlap parameter.

### Genomic structural equation modelling

The Kaiser criterion indicated that the one-factor model provided the best-fitting representation of the genetic covariance structure across SCZ and the examined SUDs (Figure S2). The EFA showed that this single-factor solution accounted for 54% of the total variance, with a sum of squared loadings (SS loadings) of 2.71. The proportion of variance for each trait explained by the model ranged from 0.14 for SCZ to 0.80 for CanUD (Table S4).

Confirmatory factor analysis (CFA) supported this structure, with statistically significant standardized loadings observed for all traits: 0.88 ± 0.11 for CanUD, 0.82 ± 0.13 for OUD, 0.83 ± 0.13 for PAU, 0.39 ± 0.28 for SCZ, and 0.65 ± 0.20 for TUD (Figure 2, Table S4). The proportion of trait variance explained by the common latent factor ranged from 22.6% for SCZ to 78.14% for CanUD (Table S4).

**Figure 2.**
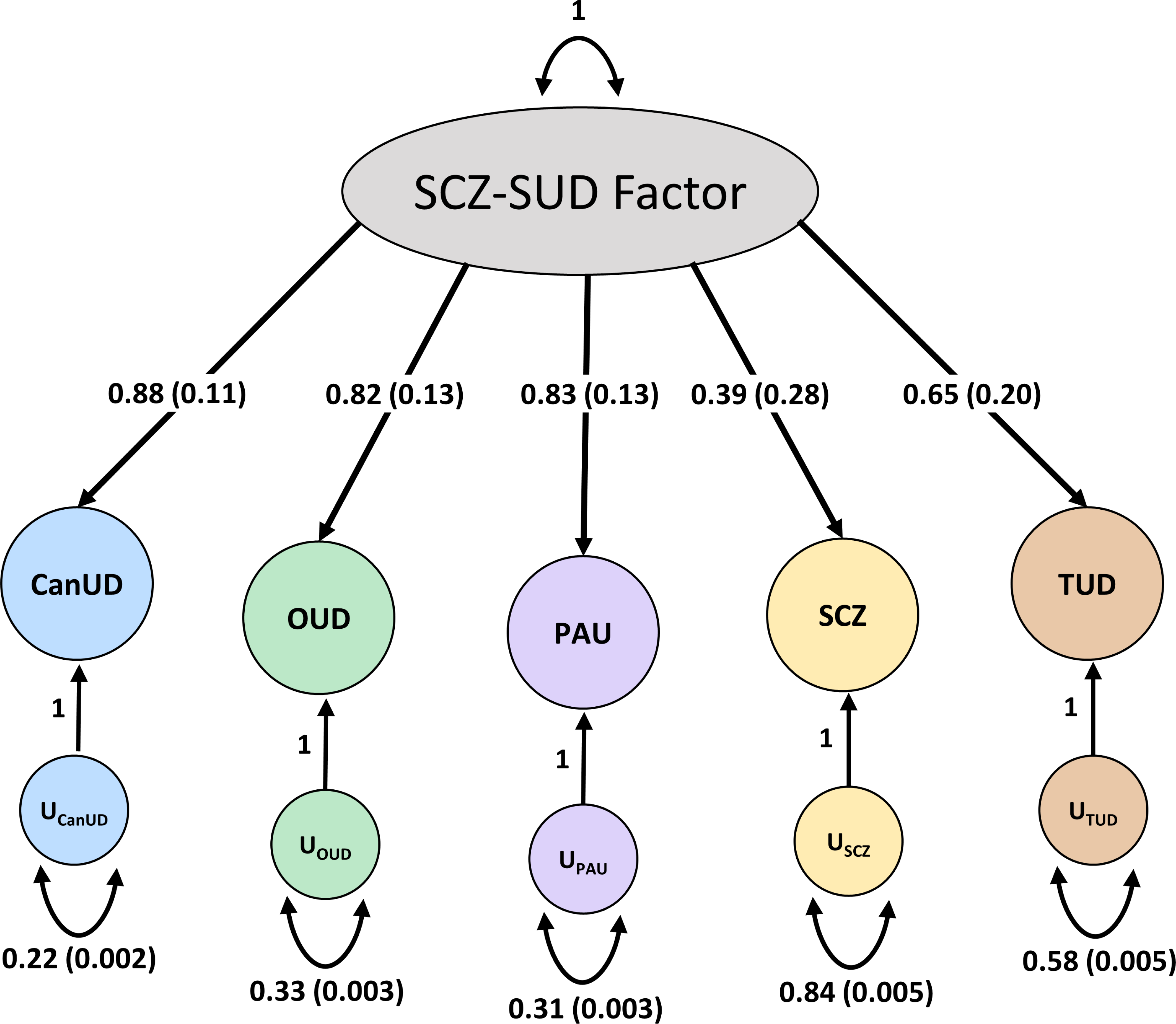
Genomic structural equation modelling of schizophrenia and substance use disorders. Values correspond to standardized loadings with standard error. Arrows pointing out from the general SCZ-SUD factor indicate the loadings attributable to each disorder. U-shaped bidirectional arrows indicate the proportion of variance unexplained by the general factor according to the EFA results. AF: Addiction factor; CanUD: Cannabis use disorder; OUD: Opioid use disorders; PAU: Problematic alcohol use; TUD: Tobacco use disorder; SCZ: Schizophrenia; U: Unexplained variance.

Model fit indices indicated that the one-factor CFA model provided a good fit to the data (χ² (5) = 5850.86, p < 0.001; comparative fit index (CFI) = 0.96; Tucker-Lewis index (TLI) = 0.91; standardized root mean square residual (SRMR) = 0.05). However, the root mean square error of approximation (RMSEA) remained elevated (RMSEA = 0.14; 90% CI: 0.14–0.15), likely reflecting the low number of degrees of freedom of the model.

### Polygenic risk scoring

PRS analyses revealed significant cross-trait associations between genetic liability to several SUDs and SCZ diagnosis (Figure 3A, Table S5). In both the CIBERSAM and the PsyCourse SCZ samples, CanUD- and PAU-derived PRS were significantly associated with SCZ diagnosis. In the CIBERSAM sample, the corresponding estimates were: CanUD-PRS (OR = 1.13, FDR = 5.07×10^−3^, Nagelkerke R^2^ = 0.004) and PAU-PRS (OR = 1.23, FDR = 2.72×10^−6^, Nagelkerke R^2^ = 0.012). In the sample from the PsyCourse Study, the following estimates were observed: CanUD-PRS (OR = 1.32, FDR = 5.70×10^−4^, Nagelkerke R^2^ = 0.02) and PAU-PRS (OR = 1.24, FDR = 6.07×10^−3^, Nagelkerke R^2^ = 0.014). OUD-PRS was positively associated with SCZ diagnosis in the CIBERSAM sample (OR = 1.11, FDR = 1.90×10^−2^, Nagelkerke R^2^ = 0.003) but not in the PsyCourse one (OR = 1.16, FDR = 5.69×10^−2^, Nagelkerke R^2^ = 0.006). On the contraty, TUD-PRS was not associated with SCZ diagnosis in either sample (CIBERSAM: OR = 1.01, FDR = 0.834; PsyCourse: OR = 1.15, FDR = 5.92×10^−2^). A lack of association was also revealed between AF-derived PRS and SCZ diagnosis (CIBERSAM: OR = 1.05, FDR = 0.262; PsyCourse: OR = 0.99, p-value = 0.874) (Figure 3A, Table S5).

**Figure 3.**
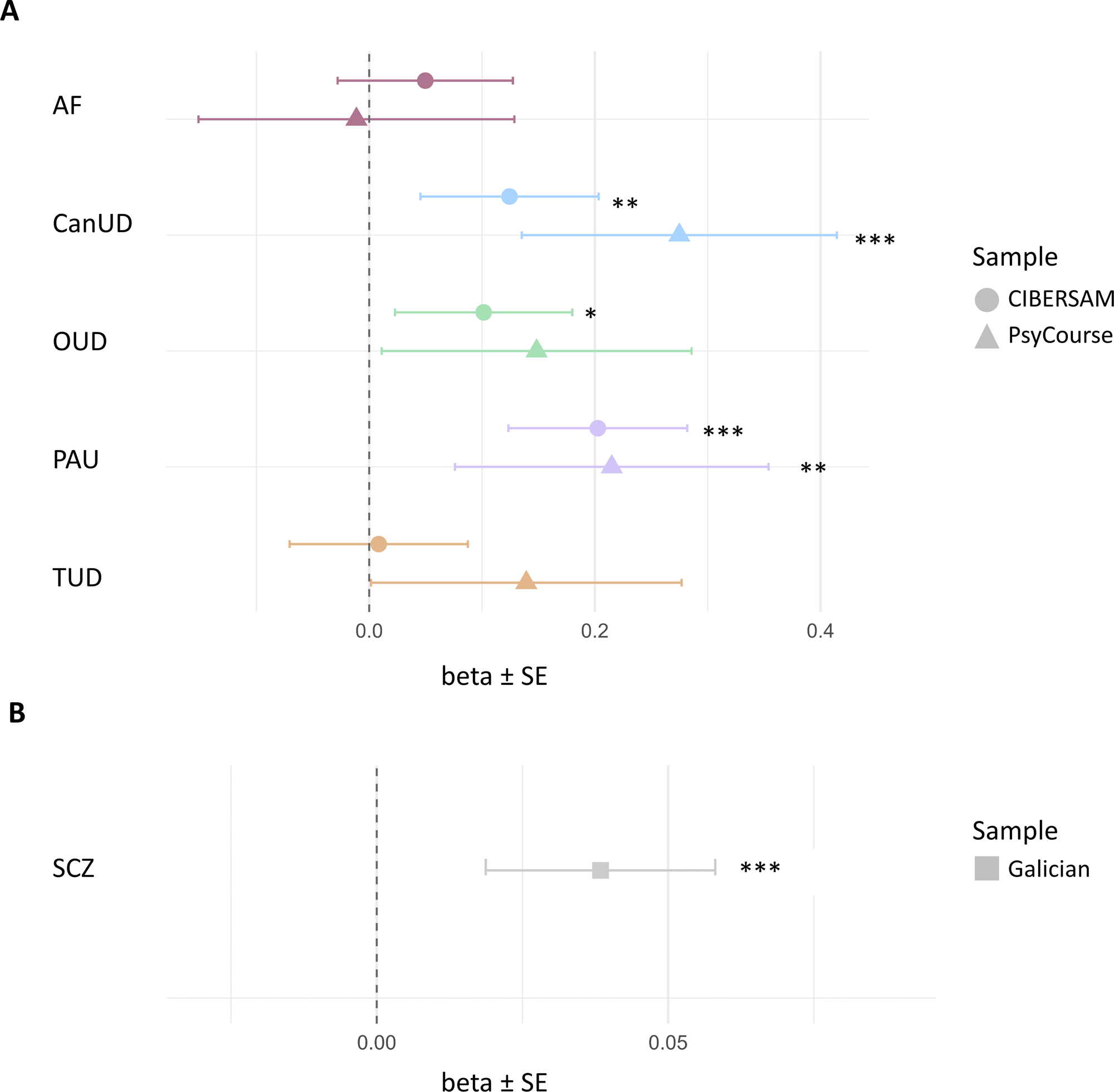
Cross-trait polygenic risk score association analyses. (A) Association of substance use disorder-informed polygenic risk scores with schizophrenia diagnosis in the CIBERSAM and PsyCourse samples. (B) Association of schizophrenia-informed polygenic risk scores with substance use disorder diagnosis in the Galician sample. The x-axis represents the model estimates and standard error of SCZ- and polysubstance use disorder-PRS in A and B, respectively. Asterisks denote the level of significance: *p-value <0.05, **p-value <0.01, ***p-value <0.001. PRS: Polygenic risk score; AF: Addiction factor; CanUD: Cannabis use disorder; OUD: Opioid use disorders; PAU: Problematic alcohol use; TUD: Tobacco use disorder; SCZ: Schizophrenia.

In the reverse direction, SCZ-derived PRS showed a significant positive association with SUD diagnosis in the Galician SUD sample (OR = 1.19, p-value = 1.42×10^−4^, Nagelkerke R^2^ = 0.009), indicating that genetic liability to SCZ also contributes to SUD risk (Figure 3B, Table S6).

### Genetically informed causal inference analysis

Mendelian randomization analyses using the IVW approach revealed significant positive bidirectional causal relationship between SCZ and CanUD, OUD and PAU, as well as a unidirectional causal effect of SCZ on TUD (Table S7). However, after the weighted median, weighted mode, MR-PRESSO global test and Steiger filtering sensitivity analyses, only the bidirectional causal relationship between SCZ and CanUD remained statistically robust (Figure 4, Table S7). The deviation from the NONE assumption was weak in either direction SCZ → CanUD: I^2^ = 0.62; CanUD → SCZ: I^2^ = 0.61), and thus the SIMEX strategy was applied to further assess sensitivity. However, no evidence of pleiotropy was found among the instrument used, as revealed by the lack of significancy of the SIMEX intercept test in either direction (SCZ → CanUD: −0.007 ± 0.008, p-value = 0.362; CanUD → SCZ: 0.017 ± 0.013, p-value = 0.224), therefore, results from the SIMEX slope were not considered to evaluate causality.

**Figure 4.**
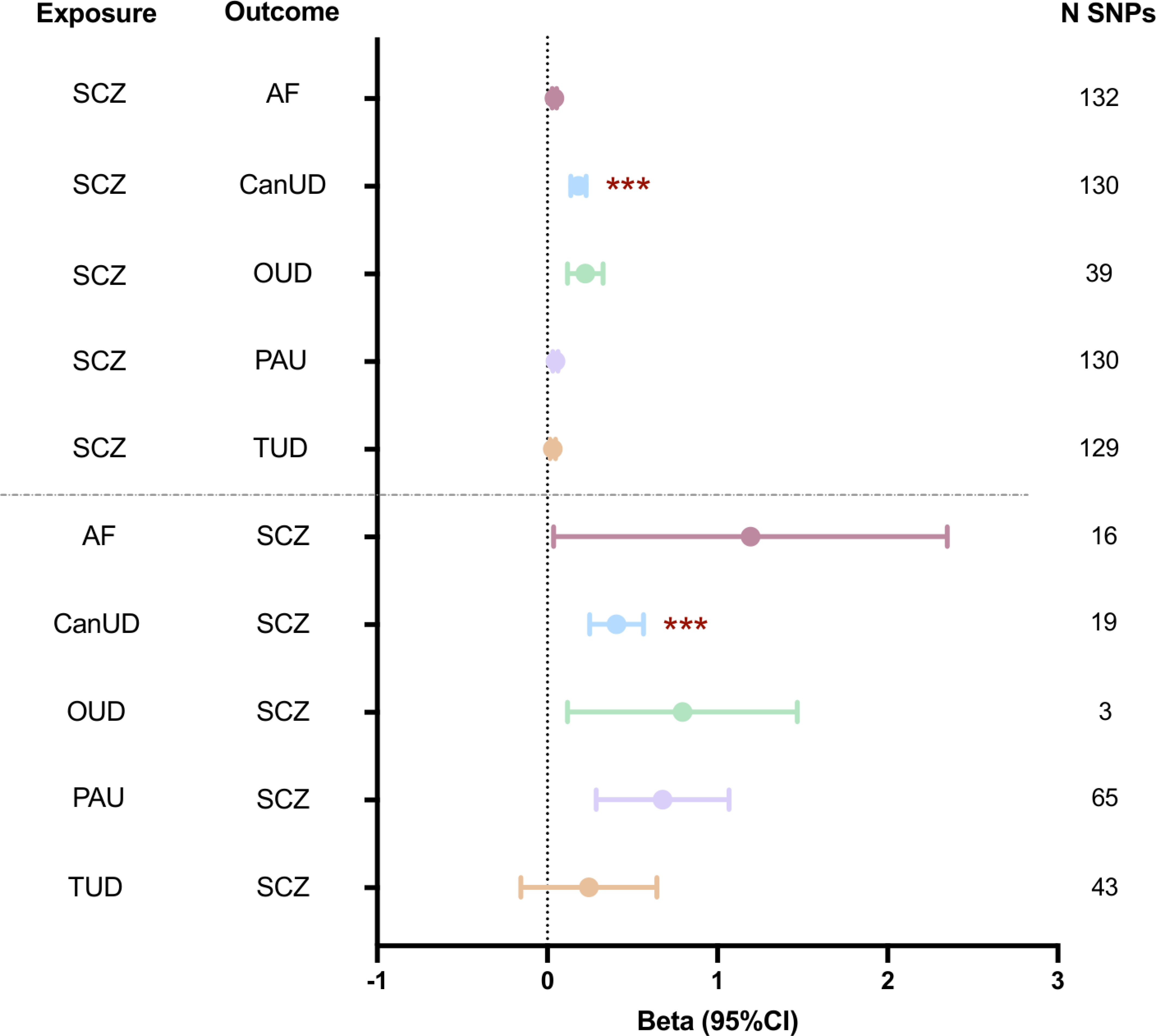
Bi-directional two-sample Mendelian randomization analyses using schizophrenia as both exposure and outcome. Effect size and 95 % confidence intervals are reported for the inverse variance-weighted (IVW) effect estimates. Evidence of causal relationship was considered if the same direction of effect as the inverse variance-weighted approach and a p-value <0.05 was obtained all sensitivity analyses. Asterisks denote the level of significance in the IVW test: *p-value <0.05, **p-value <0.01, ***p-value <0.001. AF: Addiction factor; CanUD: Cannabis use disorder; CI: confidence interval; OUD: Opioid use disorders; PAU: Problematic alcohol use; TUD: Tobacco use disorder; SCZ: Schizophrenia; SNP: single nucleotide polymorphism.

### Horizontal pleiotropy analyses

Horizontal pleiotropy analyses performed using PolarMorphism identified pleiotropic loci between SCZ and all examined traits. Specifically, 199 loci were identified for SCZ-AF, 64 for SCZ-CanUD, 2 for SCZ-OUD, 128 for SCZ-PAU, and 115 for SCZ-TUD (Tables S8-S12). Among them, 27 loci identified for SCZ-AF, 6 for SCZ-CanUD, 1 for SCZ-OUD, 22 for SCZ-PAU and 1 for SCZ-TUD converged in statistically significant correlated regions between the same trait pairs according to LAVA analyses (Tables S8-S12).

Several previously reported pleiotropic loci were replicated in the present study, including 14 independent loci jointly associated with SCZ and alcohol use disorder [14] (Table S11); 7 independent loci jointly associated with SCZ and CanUD [15] (Table S9), and 6 independent loci jointly associated with SCZ and tobacco smoking [15] (Table S12).

Some pleiotropic loci were shared across multiple trait pairs. These included loci common to SCZ-AF and SCZ-CanUD (lead SNP: rs9420), SCZ-AF and SCZ-PAU (lead SNP: rs1796518), and SCZ-AF, SCZ-CanUD and SCZ-PAU (lead SNP: rs4702), all showing concordant pleiotropic effects across trait pairs. By contrast, two loci (lead SNPs: rs11210201 and rs11649759) showed opposite pleiotropic effects between SCZ-AF and SCZ-PAU.

### Biological characterization of pleiotropic variants shared between schizophrenia and substance use disorders

Functional annotation of the pleiotropic loci identified by PolarMorphism revealed hundreds of candidate genes potentially contributing to the co-occurrence of SCZ-CanUD, SCZ-PAU and SCZ-TUD; whereas only one gene, *TSNARE1*, was annotated for SCZ-OUD (Table S13). GO enrichment analyses showed that genes uniquely annotated for SCZ-CanUD were enriched in pathways related to neuronal plasticity and chemical synapsis, whereas those for SCZ-PAU were enriched in urate transport and negative regulation of cellular catabolic processes, and those genes linked to SCZ-TUD were enriched in pathways involved in cell cycle regulation, neuronal plasticity, and proteostasis (Table S14).

To identify biological mechanisms shared across different SCZ-SUD pairs, we examined the overlap of genes annotated to pleiotropic loci identified for each trait pair. Shared genes were identified between SCZ-CanUD and SCZ-PAU (N=49), SCZ-CanUD and SCZ-TUD (N=35), SCZ-CanUD, SCZ-PAU and SCZ-TUD (N=40), SCZ-OUD and SCZ-PAU (N=1), and SCZ-PAU and SCZ-TUD (N=86) (Figure 5A, Table S15). GO enrichment analyses of these shared gene sets identified biological processes related to neuroplasticity, synaptic transmission, the immune system, metabolism and proteolysis, highlighting biological pathways that may contribute to the SCZ-SUD comorbidity (Figure 5B-F, Table S16).

**Figure 5.**
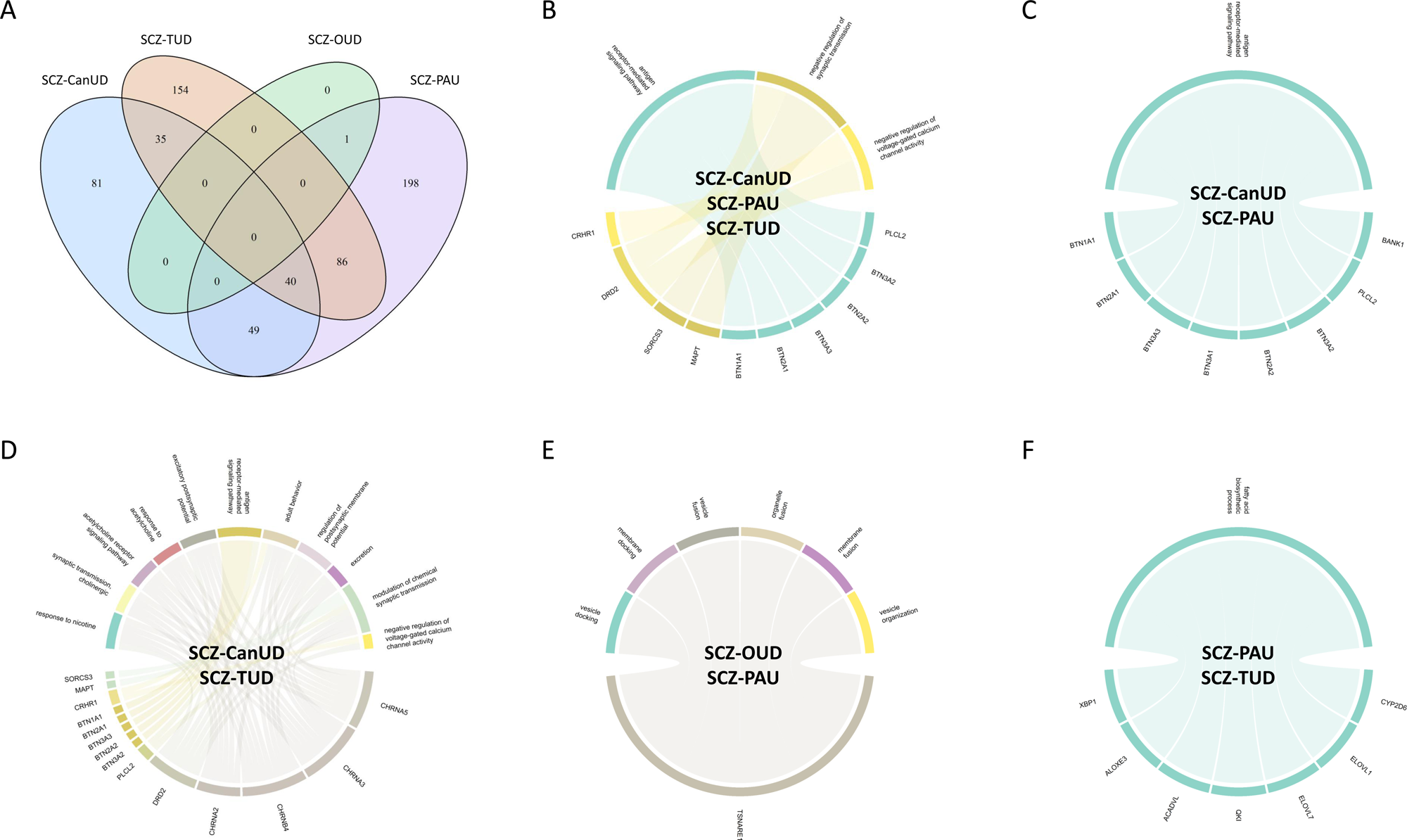
Convergence of genes and biological processes among schizophrenia-substance use disorder pairs. (A) Venn diagram illustrating the overlap of annotated genes for pleiotropic risk loci among schizophrenia-substance use disorder pairs. (B-F) Chord diagrams showing overlapping genes and biological processes between schizophrenia-substance use disorder pairs. CanUD: Cannabis use disorder; OUD: Opioid use disorders; PAU: Problematic alcohol use; TUD: Tobacco use disorder; SCZ: Schizophrenia.

## Discussion

In this study, we applied a comprehensive set of genetically informed approaches to characterize the shared genetic architecture between SCZ and SUDs. Our findings provide converging evidence that SCZ and SUDs share a complex genetic architecture characterized by both common and SCZ-SUD pair-specific genetic mechanisms.

Genome-wide genetic correlation analyses using LDSC revealed moderate positive genetic correlations between SCZ and AF, CanUD, OUD and PAU, whereas the correlation with TUD was substantially lower. MiXeR yielded correlation estimates of similar magnitude and direction, supporting the robustness of these findings. These results are consistent with previous studies reporting lower genome-wide genetic correlation between SCZ and nicotine-related phenotypes than between SCZ and CanUD, OUD, or PAU [18,19].

Local genetic correlation analyses further refined these findings by showing that the shared genetic architecture is not uniformly distributed across the genome. Instead, only a limited number of genomic regions exhibited significant local genetic correlations between SCZ and SUDs. Our results extend previous evidence of the existence of locally correlated regions between SCZ and CanUD [17,18] and between SCZ and PAU [19], while identifying, to our knowledge, the first locally correlated regions between SCZ and OUD, TUD and AF. Consistent with the positive genome-wide genetic correlations, most of these regions showed positive local correlations. However, a small subset displayed negative genetic correlations, suggesting that some genomic regions may exert opposite effects on SCZ and SUD-related phenotypes. These findings highlight that the shared genetic architecture between SCZ and SUDs is not uniformly concordant across the genome, but rather comprises genomic regions with both concordant and discordant effects.

We observed partial overlap of locally correlated regions between SCZ-AF and the individual SCZ-SUDs pairs, consistent with AF capturing genetic factors shared across multiple SUDs. Moreover, the recurrence of specific locally correlated regions across several SCZ-SUD pairs suggests the existence of convergent genomic loci contributing to the shared genetic architecture of these disorders. Nevertheless, the presence of numerous SCZ-SUD pair-specific regions indicates that each pair also retains partially distinct genetic architecture.

MiXeR analyses suggested a moderate polygenic overlap between SCZ and SUDs, exceeding 50% for SCZ-CanUD, 40% for SCZ-PAU and SCZ-TUD, and 25% for SCZ-OUD. For all trait pairs, the genetic correlation within the shared polygenic component (r_gs_) exceeded the genome-wide genetic correlation (r_g_), although the difference was less pronounced for SCZ-TUD. Consistent with the predominance of positively over negatively correlated regions identified by LAVA, these findings suggest that variants contributing to the shared polygenic component tend to exert concordant effects on SCZ and SUDs. However, the MiXeR estimates for SCZ-OUD and SCZ-TUD should be interpreted with caution, as the log-likelihood profiles and AIC comparisons with the reference models indicated limited robustness of these results.

The existence of a common genetic architecture among SCZ and SUDs suggested by the genetic correlation and polygenic overlap analyses was further supported by the GenomicSEM results. According to the Kaiser criterion, a one-factor model provided the best representation of the shared genetic structure. This common latent factor explained a moderate proportion of the genetic variance in SCZ (∼23%), while accounting for a substantially larger proportion of the genetic variance in the different SUD traits (>60% in all cases). Factor loadings were similar for CanUD, OUD, and PAU, but lower for TUD, suggesting that the genetic component shared between SCZ and SUDs is less strongly represented in TUD than in the other SUDs.

These findings are consistent with previous GenomicSEM studies supporting a moderate shared genetic architecture between SCZ and SUDs. A recent GenomicSEM analysis including SCZ, CanUD, OUD, PAU, nicotine use disorder, and nine additional psychiatric conditions identified a five-factor model in which the SUD factor was moderately correlated with a factor defined by SCZ and bipolar disorder (r=0.44) [19]. Similarly, another GenomicSEM analysis including CanUD, SCZ, and seven additional psychiatric conditions identified a three-factor model in which the factor including CanUD was mildly correlated with the factor including SCZ [18], and the cannabis use phenotype loaded to both SCZ and CanUD-including factors [18]. Together, these findings support the existence of a moderate but robust shared genetic architecture between SCZ and SUDs.

Consistent with these findings, PRSs derived from CanUD, and PAU were consistently associated with SCZ diagnosis across both SCZ samples, whereas the OUD-derived PRS was associated with SCZ diagnosis in only one of the samples. The TUD-derived PRS showed a suggestive association with SCZ diagnosis in one of the samples, however it did not survive the multiple testing correction. In the reverse direction, SCZ-derived PRS was associated with SUD diagnosis in the SUD sample, replicating previous findings based on a less powered SCZ GWAS [50]. The weaker and less consistent findings for TUD and OUD are in line with the LDSC, MiXeR and GenomicSEM results, all of which suggested a smaller shared genetic component between SCZ and TUD or OUD than between SCZ and the other SUDs. Differences in comorbidity patterns across samples may also have contributed to the variability observed in the SUD-informed PRS associations. In contrast, the AF-derived PRS was not associated with SCZ diagnosis in either sample despite the positive genome-wide genetic correlation observed between SCZ and AF. These results likely reflect the fact that genome-wide genetic correlation and PRS performance capture different aspects of genetic architecture. While genome-wide genetic correlation reflects the overall concordance of genetic effects across the genome, PRS performance depends on the predictive value of individual variants. Because AF captures genetic influences shared across multiple SUDs, these shared effects may be less informative for predicting SCZ than the disorder-specific genetic components represented by individual SUD PRSs.

Mendelian randomization analyses added an important dimension suggesting bidirectional causal effects between SCZ and CanUD, consistent with a recent study that applied a different Mendelian randomization framework to the same GWAS datasets [18]. These findings suggest that part of the genetic liability to SCZ may increase the risk of developing CanUD and, conversely, that genetic liability to CanUD may contribute to SCZ risk. This bidirectional causal effect is also plausible from a clinical and developmental perspective. Genetic liability to SCZ may increase vulnerability to CanUD [13], potentially through behavioural dysregulation or self-medication mechanisms. Conversely, cannabis use has been linked to an earlier onset of SCZ [51].

At the locus level, the PolarMorphism analyses identified numerous previously unreported pleiotropic loci, most of which were specific to individual SCZ-SUD pairs, while a subset was shared across multiple SCZ-SUD pairs. These findings further support the existence of pair-specific and broadly shared genetic mechanisms underlying the shared genetic architecture between SCZ and SUDs.

The enrichment of genes mapped to pleiotropic loci for individual SCZ-SUD pairs revealed distinct pathway profiles underlying each comorbidity. Genes uniquely annotated for SCZ-CanUD were enriched in pathways related to neuronal plasticity and chemical synaptic transmission, whereas those unique for SCZ-PAU were enriched in urate transport and the negative regulation of cellular catabolic processes. On the other hand, genes associated with SCZ-TUD were enriched in pathways involved in cell cycle regulation, neuronal plasticity, and proteostasis. The findings for SCZ-OUD were more limited, likely reflecting the lower statistical power of the OUD GWAS, with *TSNARE1* being the only gene annotated for this pair.

Nevertheless, enrichment analyses of shared mapped genes across different SCZ-SUD pairs identified convergent biological mechanisms. The greatest overlap was observed among genes shared by SCZ-CanUD, SCZ-PAU, and SCZ-TUD, which were enriched in pathways related to synaptic transmission and antigen receptor-mediated signalling. Additional convergence was observed for genes shared between SCZ-CanUD and SCZ-TUD, which were enriched in cholinergic synaptic transmission. Likewise, genes shared between SCZ-PAU and SCZ-TUD were enriched in fatty acid biosynthetic processes. Finally, *TSNARE1*, the only gene annotated for SCZ-OUD, was also identified in SCZ-PAU, suggesting that disruption of vesicle trafficking and membrane fusion processes may contibute to both comorbidities.

Together, these findings support the hypothesis that the genetic architecture underlying SCZ– SUD comorbidities is shaped by both shared and disorder-specific mechanisms. The identification of these convergent pathways may facilitate the development of more precise therapeutic strategies tailored to biologically defined patient subgroups.

Several limitations of this study should be considered. First, there is a substantial imbalance in statistical power across the SUD GWAS datasets included, largely driven by differences in sample size. Consequently, some non-significant findings may reflect limited power rather than a true absence of shared genetic architecture. This issue is particularly relevant for MR analyses, where the number of genetic instruments used to detect causality is limited. Second, phenotypic heterogeneity and measurement bias across SUD definitions should be considered. The SUD phenotypes included in this study derive from GWAS meta-analyses that differ in diagnostic criteria, assessment methods, and phenotype definitions (e.g., clinical diagnoses based on DSM/ICD criteria versus self-reported measures or questionnaire-based constructs such as AUDIT-derived problematic alcohol use). This heterogeneity may introduce variability in the underlying genetic signals. Third, differences in control definitions, particularly in cohorts where substance use was not systematically assessed, may result in misclassification and attenuation of associations. Fourth, our analyses were restricted to individuals of European ancestry. This reflects both the greater availability of large-scale GWAS data in European populations and the fact that European ancestry predominates in the samples analysed. While this approach reduces population stratification, it limits the generalizability of our findings to other ancestral groups.

In conclusion, this study provides comprehensive evidence for a shared genetic architecture between SCZ and SUDs, largely driven by pleiotropic variants exerting concordant effect across disorders. Our findings further demonstrate that this shared genetic component comprises both SCZ-SUD pair-specific and broadly shared genetic mechanisms and biological pathways. In addition, we found evidence supporting a bidirectional causal relationship between SCZ and CanUD. Together, these findings advance our understanding of the biological basis underlying the co-occurrence of SCZ and SUDs and provide a framework for future studies aimed at identifying biologically more homogeneous patient subgroups and developing more targeted prevention and personalised treatment strategies.

## Data Availability

All data produced in the present work are contained in the manuscript.

## Author contributions

SA: Data curation, Investigation, Visualization, Writing–original draft. DK: Data curation, Investigation, Visualization, Writing–review and editing. MSA: Investigation, Writing–review and editing. SP, AMP-G, JG-P, MB, PJ-F, CA, KA, EV, GM, LM, MH, MDM, OR, AN-F, JB, MOK, BC-F, DR-E, AG-P, ECS, MA, GF, FS, I-GA, VA, DED, AJF, CF, MJ, FUL, GJ, CK, JR, EZR, MS, AS, CS, JW, JZ, LF, UH, BA, PF, JC, and TGS: Resources, Writing–review and editing. MM, BC: Conceptualization, Investigation, Writing-review and editing.

