## Supplementary Methods for "Pleiotropic genetic architecture linking schizophrenia and substance use disorders"

*Quality control of the schizophrenia cohorts*

The CIBERSAM cohort was genotyped using the Illumina PsychArray (Illumina, San Diego, USA) as part of the third wave of the SCZ GWAS carried out by the Psychiatric Genomics Consortium (PGC), and quality control was performed according to the PGC pipeline. Briefly, individuals with a SNP missingness > 2% or extreme heterozygosity values were removed. Also, SNPs with a call rate < 95%, deviation from Hardy–Weinberg equilibrium ($P < 1\times10^{-6}$ in controls or $P < 1\times10^{-10}$ in SCZ patients), missingness > 2% or difference in missingness between SCZ patients and controls > 2% were excluded.

The PsyCourse cohort was genotyped using the Illumina Global Screening Arrays versions 1 and 3 (Illumina, San Diego, USA). Briefly, individuals with a SNP missingness > 2%, extreme heterozygosity values, or discrepancies between reported and genetically inferred sex were filtered out. SNPs with a call rate < 98%, deviation from Hardy–Weinberg equilibrium ($p < 1\times10^{-4}$), or minor allele frequency (MAF) < 0.5% were excluded. In addition, palindromic SNPs and variants displaying substantial MAF differences (>10%) compared with European reference populations were also removed.

In both cohorts, genotype imputation was conducted using the Michigan Imputation Server [1], and only SNPs with imputation quality scores ($R^2$) > 0.3 and MAF > 0.01 were retained.

*Quality control of the polysubstance use disorder cohort*

The SUD cohort from Galicia was genotyped using the Axiom Spain Biobank Array (Thermo Fisher Scientific) at the Spanish National Center for Genotyping (CEGEN, Santiago de Compostela, Spain). The array included ~750.000 SNPs, designed by CEGEN for better coverage of Spanish variants than generic global arrays. Briefly, quality controls included removal of individuals and SNPs with missingness > 2%, removal of SNPs with deviation from Hardy–Weinberg equilibrium ($p < 1\times10^{-}5$), removal of SNPs with MAF differences > 15% compared with the 1000 Genomes Project European Samples, and removal of individuals with discrepancies between reported and genetically inferred sex. Data were prepared for imputation using a pre-imputation data-preparation toolkit from the McCarthy Group (https://www.chg.ox.ac.uk/~wrayner/tools/). Imputation was performed using The TOPMed Imputation Panel [2] and Server [3], and only SNPs with imputation quality scores (R2) > 0.3 and MAF > 0.01 were retained.
