## Supplementaty Tables for "Pleiotropic genetic architecture linking schizophrenia and substance use disorders"

**Table S1. Genetic correlation between schizophrenia, substance use disorders and addiction factor.**

| Trait 1 | Trait 2 | rg | se | z | p-value | h2_obs | h2_obs_se | h2_int | h2_int_se | gcov_int | gcov_int_se |
|---|---|---|---|---|---|---|---|---|---|---|---|
| SCZ | PAU | 0,3172 | 0,0248 | 12,7660 | 2,52E-37 | 0,0344 | 0,0018 | 0,9955 | 0,0118 | 0,0241 | 0,0091 |
| SCZ | AUD | 0,3417 | 0,0213 | 16,0380 | 6,95E-58 | 0,2452 | 0,0118 | 10.204 | 0,0095 | 0,0261 | 0,0065 |
| SCZ | OUD | 0,3641 | 0,0489 | 74,4880 | 9,42E-14 | 0,0299 | 0,0034 | 10.041 | 0,0173 | 0,0153 | 0,014 |
| SCZ | CanUD | 0,3728 | 0,0227 | 16,4200 | 1,38E-60 | 0,0238 | 0,0013 | 0,9934 | 0,0075 | 0,006 | 0,0059 |
| SCZ | TUD | 0,1001 | 0,022 | 4,5510 | 5,34E-06 | 0,024 | 0,0012 | 0,9815 | 0,0074 | 0,0047 | 0,006 |
| SCZ | AF | 0,3537 | 0,0259 | 13,6690 | 1,55E-42 | 0,0214 | 0,0009 | 0,9922 | 0,0103 | 0,0183 | 0,0084 |
| PAU | AUD | 0,9986 | 0,0078 | 128,5740 | 0,00E+00 | 0,2549 | 0,0143 | 10.071 | 0,0114 | 0,7164 | 0,011 |
| PAU | OUD | 0,6872 | 0,0567 | 12,1210 | 8,10E-34 | 0,0321 | 0,0039 | 0,9869 | 0,0208 | 0,1288 | 0,0184 |
| PAU | CanUD | 0,7143 | 0,0278 | 25,6480 | 4,39E-145 | 0,0238 | 0,0016 | 0,9995 | 0,0097 | 0,0808 | 0,0078 |
| PAU | TUD | 0,5881 | 0,0234 | 25,1370 | 1,95E-139 | 0,0245 | 0,0014 | 0,977 | 0,0101 | 0,0381 | 0,0074 |
| PAU | AF | 0,9153 | 0,0142 | 64,6470 | 0,00E+00 | 0,0214 | 0,0012 | 0,985 | 0,0116 | 0,5 | 0,0115 |
| AUD | OUD | 0,7909 | 0,0454 | 17,4330 | 4,56E-68 | 0,03 | 0,0035 | 10.043 | 0,0172 | 0,1544 | 0,0146 |
| AUD | CanUD | 0,7746 | 0,0232 | 33,3420 | 9,54E-244 | 0,0239 | 0,0013 | 0,9934 | 0,0081 | 0,1153 | 0,0061 |
| AUD | TUD | 0,6429 | 0,0209 | 30,7900 | 3,61E-208 | 0,0239 | 0,0012 | 0,9827 | 0,008 | 0,0535 | 0,0059 |
| AUD | AF | 0,9115 | 0,017 | 53,5360 | 0,00E+00 | 0,0214 | 0,0009 | 0,9923 | 0,0105 | 0,3474 | 0,0084 |
| OUD | CanUD | 0,7304 | 0,0584 | 12,5160 | 6,12E-36 | 0,0305 | 0,0028 | 0,9861 | 0,0226 | 0,2011 | 0,0147 |
| OUD | TUD | 0,4747 | 0,0594 | 7,9880 | 1,37E-15 | 0,0459 | 0,0031 | 10.392 | 0,0267 | 0,0671 | 0,0153 |
| OUD | AF | 0,7493 | 0,0504 | 14,8680 | 5,29E-50 | 0,0211 | 0,0016 | 10.130 | 0,0208 | 0,3676 | 0,0154 |
| CanUD | TUD | 0,5951 | 0,0275 | 21,6660 | 4,28E-104 | 0,0241 | 0,0011 | 0,9805 | 0,0072 | 0,0362 | 0,0055 |
| CanUD | AF | 0,7439 | 0,0238 | 31,2090 | 8,03E-214 | 0,0214 | 0,001 | 0,9927 | 0,0106 | 0,1954 | 0,0083 |
| TUD | AF | 0,673 | 0,023 | 29,2390 | 6,14E-188 | 0,0214 | 0,0009 | 0,9919 | 0,0103 | 0,0668 | 0,0077 |

SCZ: Schizophrenia; AF: Addiction Factor; CanUD: Cannabis Use Disorder; OUD: Opioid Use Disorder; PAU: Problematic Alcohol Use; TUD: Tobacco Use Disorder

**Table S2. Genomic regions displaying local genetic correlations between schizophrenia, substance use disorders and addiction factor.**

| Locus | Chr | Start | Stop | N SNPs | N PCS | rho | rho (lower) | rho (upper) | r2 | r2 (lower) | r2 (upper) | p | FDR | Genes in region |
|---|---|---|---|---|---|---|---|---|---|---|---|---|---|---|
| **SCZ - CanUD** | | | | | | | | | | | | | | |
| 1719 | 11 | 112755447 | 113889019 | 2663 | 262 | 0,541094 | 0,37754 | 0,7103 | 0,292783 | 0,14254 | 0,50453 | 2,4724E-09 | 4,5987E-06 | *NCAM1, TTC12, ANKK1, DRD2, TMPRSS5, ZW10, CLDN25, USP28,* |
| 2244 | 18 | 5575813 | 6756496 | 3024 | 359 | 1 | 0,57888 | 1 | 1 | 0,3351 | 1 | 1,8625E-07 | 0,00017322 | *EPB41L3, TMEM200C, L3MBTL4, C18orf64, ARHGAP28* |
| 1273 | 8 | 27406512 | 28344176 | 2474 | 274 | 0,574613 | 0,37011 | 0,79468 | 0,33018 | 0,13698 | 0,63152 | 4,5653E-07 | 0,00028305 | *CLU, SCARA3, CCDC25, ESCO2, PBK, SCARA5, NUGGC, ELP3, PNOC,* |
| 1640 | 11 | 27018308 | 28577718 | 2539 | 243 | 0,679074 | 0,41427 | 0,98669 | 0,461141 | 0,17162 | 0,97356 | 3,618E-06 | 0,00168239 | *FIBIN, BBOX1, CCDC34, LGR4, LIN7C, BDNF, KIF18A, METTL15* |
| 611 | 4 | 11050120 | 12209028 | 3469 | 291 | 1 | 0,5152 | 1 | 1 | 0,26543 | 1 | 6,039E-06 | 0,00224652 | *HS3ST1* |
| 1668 | 11 | 57675987 | 58452708 | 1949 | 109 | 1 | 0,58036 | 1 | 1 | 0,33682 | 1 | 1,3779E-05 | 0,00427137 | *OR9Q1, OR6Q1, OR9I1, OR9Q2, OR1S2, OR1S1, OR10Q1, OR10W1,* |
| 1190 | 7 | 104109232 | 105312563 | 2297 | 252 | 0,531181 | 0,30533 | 0,78353 | 0,282154 | 0,09323 | 0,61391 | 1,866E-05 | 0,00489317 | *LHFPL3, KMT2E, SRPK2, PUS7, RINT1, EFCAB10, ATXN7L1* |
| 2484 | 22 | 40378784 | 41722787 | 1888 | 203 | 0,665854 | 0,38393 | 1 | 0,443361 | 0,1474 | 1 | 2,1046E-05 | 0,00489317 | *FAM83F, TNRC6B, ADSL, SGSM3, MKL1, MCHR1, SLC25A17, ST13,* |
| 278 | 2 | 72160303 | 73544290 | 2120 | 247 | 0,779219 | 0,43298 | 1 | 0,607182 | 0,18747 | 1 | 4,7864E-05 | 0,00973703 | *CYP26B1, EXOC6B, SPR, EMX1, SFXN5, RAB11FIP5, NOTO, SMYD5,* |
| 1272 | 8 | 26679136 | 27406511 | 2317 | 191 | 0,729562 | 0,41196 | 1 | 0,532261 | 0,16971 | 1 | 5,235E-05 | 0,00973703 | *ADRA1A, RP11-521M14.2, STMN4, TRIM35, PTK2B, CHRNA2,* [illegible] |
| 861 | 5 | 102903986 | 103788460 | 1987 | 256 | 0,518463 | 0,28606 | 0,78944 | 0,268804 | 0,08183 | 0,62322 | 6,2283E-05 | 0,00986414 | |
| 1754 | 12 | 13559528 | 14656849 | 2881 | 276 | 1 | 0,51237 | 1 | 1 | 0,26253 | 1 | 6,8856E-05 | 0,00986414 | *GRIN2B, ATF7IP, PLBD1* |
| 2233 | 17 | 76596288 | 77412786 | 2706 | 361 | 0,686007 | 0,37191 | 1 | 0,470606 | 0,13832 | 1 | 6,8943E-05 | 0,00986414 | *CYTH1, USP36, TIMP2, DDC8, LGALS3BP, CANT1, C1QTNF1-AS1,* [illegible] |
| 1127 | 7 | 28890887 | 30175113 | 3636 | 323 | 0,64126 | 0,33698 | 1 | 0,411215 | 0,11356 | 1 | 0,00010024 | 0,01331747 | *CPVL, CHN2, PRR15, WIPF3, SCRN1, FKBP14, PLEKHA8, MTURN* |
| 766 | 4 | 182946259 | 183603892 | 1990 | 298 | 0,48693 | 0,25012 | 0,75179 | 0,237101 | 0,06256 | 0,56518 | 0,00012716 | 0,01576772 | *TENM3* |
| 2139 | 16 | 58508979 | 59885499 | 3583 | 282 | 0,540317 | 0,28129 | 0,83303 | 0,291943 | 0,07913 | 0,69394 | 0,00014882 | 0,01730079 | *NDRG4, SETD6, CNOT1, SLC38A7, GOT2, RP11-105C20.2* |
| 2281 | 18 | 52512524 | 53762996 | 2220 | 240 | 0,561951 | 0,28014 | 0,94805 | 0,315789 | 0,07848 | 0,8988 | 0,00021197 | 0,02319212 | *RAB27B, CCDC68, TCF4* |
| 1175 | 7 | 85356716 | 86446538 | 2526 | 158 | 0,643748 | 0,33084 | 1 | 0,414412 | 0,10945 | 1 | 0,00026136 | 0,0270071 | *GRM3* |
| 2026 | 14 | 99474534 | 100786189 | 3123 | 355 | 0,482614 | 0,23385 | 0,77737 | 0,232917 | 0,05469 | 0,6043 | 0,00033306 | 0,03260453 | *BCL11B, SETD3, CCNK, CCDC85C, HHIPL1, CYP46A1, EML1, EVL,* [illegible] |
| 2164 | 16 | 83077870 | 83778371 | 3067 | 334 | 0,858406 | 0,42331 | 1 | 0,736861 | 0,17919 | 1 | 0,00039227 | 0,03648148 | *CDH13* |
| 649 | 4 | 49325139 | 53879571 | 1947 | 220 | -0,834183 | -1 | -0,39963 | 0,69586 | 0,15971 | 1 | 0,00041388 | 0,03665812 | *DCUN1D4, LRRC66, SGCB, SPATA18, USP46, ERVMER34-1,* [illegible] |
| 63 | 1 | 73992171 | 75132092 | 2023 | 222 | 0,422643 | 0,19855 | 0,65593 | 0,178627 | 0,03942 | 0,43025 | 0,00044123 | 0,03730399 | *LRRIQ3, FPGT, FPGT-TNNI3K, TNNI3K, LRRC53, C1orf173* |
| 1887 | 13 | 43931932 | 45476446 | 3239 | 303 | 0,648383 | 0,29966 | 1 | 0,420401 | 0,0898 | 1 | 0,00048875 | 0,03952468 | *ENOX1, CCDC122, LACC1, SMIM2, SERP2, TSC22D1* |
| **SCZ - OUD** | | | | | | | | | | | | | | |
| 1367 | 8 | 142611890 | 143611461 | 259 | 48 | 1 | 0,6958 | 1 | 1 | 0,48414 | 1 | 4,9354E-08 | 1,0167E-05 | *TSNARE1, BAI1* |
| 2090 | 15 | 90632719 | 91593623 | 148 | 27 | 0,900763 | 0,66918 | 1 | 0,811373 | 0,44781 | 1 | 9,689E-07 | 9,9797E-05 | *IDH2, SEMA4B, CIB1, GDPGP1, TTLL13, RP11-697E2.6, NGRN,* [illegible] |
| 950 | 6 | 25684630 | 26396200 | 201 | 31 | 0,813284 | 0,52758 | 1 | 0,661431 | 0,27834 | 1 | 2,4415E-05 | 0,0016765 | *SCGN, HIST1H2AA, HIST1H2BA, SLC17A4, SLC17A1, SLC17A3,* [illegible] |
| 951 | 6 | 26396201 | 27261035 | 201 | 25 | 0,961701 | 0,61918 | 1 | 0,924868 | 0,38338 | 1 | 0,0001565 | 0,0080599 | *BTN3A1, BTN3A3, BTN2A1, BTN1A1, HMGN4, ABT1, ZNF322,* [illegible] |
| 952 | 6 | 27261036 | 28666364 | 368 | 27 | 0,747844 | 0,41818 | 1 | 0,55927 | 0,17487 | 1 | 0,00026894 | 0,0110802 | *POM121L2, ZNF391, ZNF184, HIST1H2BL, HIST1H2AI, HIST1H3H,* [illegible] |
| 953 | 6 | 28666365 | 29529755 | 376 | 27 | 0,936907 | 0,54126 | 1 | 0,877794 | 0,29296 | 1 | 0,00041201 | 0,013803 | *TRIM27, C6orf100, ZNF311, OR2W1, OR2B3, OR2J1, OR2J3,* [illegible] |
| 1719 | 11 | 112755447 | 113889019 | 194 | 39 | 0,634206 | 0,32472 | 0,98518 | 0,402217 | 0,10544 | 0,97058 | 0,00050243 | 0,013803 | *NCAM1, TTC12, ANKK1, DRD2, TMPRSS5, ZW10, CLDN25, USP28,* |
| 2124 | 16 | 24014706 | 25235251 | 106 | 39 | 1 | 0,49273 | 1 | 1 | 0,24278 | 1 | 0,00053604 | 0,013803 | *PRKCB, CACNG3, RBBP6, TNRC6A, SLC5A11, ARHGAP17, LCMT1,* [illegible] |
| 266 | 2 | 57952946 | 59251996 | 70 | 19 | 0,956017 | 0,5749 | 1 | 0,913968 | 0,33068 | 1 | 0,00085568 | 0,01958556 | *VRK2, FANCL* |
| 852 | 5 | 91956906 | 93814604 | 126 | 27 | 1 | 0,35157 | 1 | 1 | 0,12706 | 1 | 0,00242714 | 0,04999908 | *NR2F1, FAM172A, POU5F2, KIAA0825* |
| **SCZ - PAU** | | | | | | | | | | | | | | |
| 1719 | 11 | 112755447 | 113889019 | 2646 | 260 | 0,680517 | 0,53768 | 0,82866 | 0,463104 | 0,2891 | 0,68668 | 1,0548E-15 | 2,2519E-12 | *NCAM1, TTC12, ANKK1, DRD2, TMPRSS5, ZW10, CLDN25, USP28,* |
| 1301 | 8 | 64215359 | 66018204 | 3008 | 255 | 0,543596 | 0,3636 | 0,73534 | 0,295496 | 0,1322 | 0,54073 | 6,4615E-08 | 6,8976E-05 | *BHLHE22, CYP7B1* |
| 2281 | 18 | 52512524 | 53762996 | 2212 | 239 | 0,539768 | 0,34741 | 0,74308 | 0,291349 | 0,12069 | 0,55217 | 4,1581E-07 | 0,00029592 | *RAB27B, CCDC68, TCF4* |
| 844 | 5 | 82720249 | 84094797 | 2545 | 311 | 0,666962 | 0,42807 | 0,94937 | 0,444838 | 0,18324 | 0,9013 | 6,3059E-07 | 0,00033658 | *VCAN, HAPLN1, EDIL3* |
| 1637 | 11 | 24892440 | 25662918 | 2522 | 164 | -0,831564 | -1 | -0,52154 | 0,691499 | 0,272 | 1 | 2,7734E-06 | 0,00118425 | *LUZP2* |
| 364 | 2 | 180336046 | 181309939 | 2565 | 232 | 0,737783 | 0,44768 | 1 | 0,544324 | 0,20042 | 1 | 5,3236E-06 | 0,0016237 | *ZNF385B, CWC22* |
| 2386 | 20 | 15962577 | 16661516 | 2317 | 236 | 0,782152 | 0,47237 | 1 | 0,611761 | 0,22313 | 1 | 4,8623E-06 | 0,0016237 | *MACROD2, KIF16B* |
| 1184 | 7 | 97420208 | 98173564 | 1792 | 188 | 0,706514 | 0,42023 | 1 | 0,499162 | 0,17659 | 1 | 1,1143E-05 | 0,00237905 | *ASNS, OCM2, LMTK2, BHLHA15, TECPR1, BRI3, BAIAP2L1* |
| 1445 | 9 | 100310518 | 101596750 | 3099 | 304 | 0,553602 | 0,32242 | 0,82089 | 0,306475 | 0,10396 | 0,67386 | 9,1528E-06 | 0,00237905 | *TMOD1, TSTD2, NCBP1, XPA, FOXE1, C9orf156, HEMGN, ANP32B,* |
| 2208 | 17 | 44865833 | 45883901 | 2050 | 166 | 0,656556 | 0,39352 | 0,98827 | 0,431065 | 0,15486 | 0,97667 | 1,0135E-05 | 0,00237905 | *WNT3, WNT9B, GOSR2, RP11-156P1.2, RPRML, CDC27, MYL4,* [illegible] |
| 1375 | 9 | 3079046 | 4025355 | 2304 | 321 | 0,529999 | 0,30634 | 0,78363 | 0,280899 | 0,09384 | 0,61408 | 1,2454E-05 | 0,00241723 | *RFX3, AL365202.1, GLIS3* |
| 1738 | 11 | 133521643 | 134351064 | 2003 | 312 | 0,351201 | 0,19618 | 0,51331 | 0,123342 | 0,03849 | 0,26349 | 1,8113E-05 | 0,00297474 | *SPATA19, IGSF9B, JAM3, NCAPD3, VPS26B, THYN1, ACAD8,* [illegible] |
| 2385 | 20 | 15238520 | 15962576 | 2017 | 280 | 0,673687 | 0,38372 | 1 | 0,453854 | 0,14724 | 1 | 1,7281E-05 | 0,00297474 | *MACROD2* |
| 1183 | 7 | 96564982 | 97420207 | 1824 | 234 | 0,847369 | 0,47723 | 1 | 0,718034 | 0,22775 | 1 | 2,1084E-05 | 0,003001 | *DLX6, DLX5, ACN9, TAC1* |

| | | | | | | | | | | | | | | |
|---|---|---|---|---|---|---|---|---|---|---|---|---|---|---|
| 1596 | 10 | 122803507 | 123856184 | 2764 | 301 | 0,72495 | 0,4119 | 1 | 0,525553 | 0,16966 | 1 | 2,0554E-05 | 0,003001 | *FGFR2, ATE1, NSMCE4A, TACC2* |
| 2489 | 22 | 45170650 | 46351048 | 3651 | 354 | 0,935899 | 0,52091 | 1 | 0,875907 | 0,27134 | 1 | 2,3051E-05 | 0,00307587 | *PRR5-ARHGAP8, ARHGAP8, PHF21B, NUP50, KIAA0930, UPK3A, FA* |
| 1676 | 11 | 68000950 | 68887344 | 1859 | 206 | 0,638727 | 0,36346 | 0,96599 | 0,407972 | 0,1321 | 0,93313 | 2,4748E-05 | 0,00310807 | *C11orf24, LRP5, PPP6R3, GAL, MTL5, CPT1A, MRPL21, IGHMBP2, N* |
| 580 | 3 | 181424324 | 183203155 | 3190 | 334 | 0,548676 | 0,30632 | 0,83387 | 0,301045 | 0,09383 | 0,69534 | 2,817E-05 | 0,00334132 | *SOX2, ATP11B, DCUN1D1, MCCC1, LAMP3, MCF2L2, B3GNT5* |
| 2207 | 17 | 43460501 | 44865832 | 2515 | 110 | 0,547082 | 0,30922 | 0,79552 | 0,299298 | 0,09562 | 0,63285 | 3,3771E-05 | 0,00379474 | *ARHGAP27, PLEKHM1, CRHR1, SPPL2C, MAPT, STH, KANSL1, ARL1* |
| 593 | 3 | 195710291 | 196383152 | 1768 | 251 | 0,588583 | 0,31839 | 0,93778 | 0,34643 | 0,10137 | 0,87943 | 5,0834E-05 | 0,0054265 | *TFRC, ZDHHC19, SLC51A, PCYT1A, RP11-447L10.1, TCTEX1D2, TM4* |
| 1640 | 11 | 27018308 | 28577718 | 2525 | 240 | 0,480114 | 0,26306 | 0,72236 | 0,23051 | 0,0692 | 0,52181 | 5,3625E-05 | 0,00545191 | *FIBIN, BBOX1, CCDC34, LGR4, LIN7C, BDNF, KIF18A, METTL15* |
| 1854 | 12 | 126871453 | 127545377 | 2017 | 239 | -0,671625 | -1 | -0,36004 | 0,45108 | 0,12963 | 1 | 8,2155E-05 | 0,00797272 | |
| 624 | 4 | 25832019 | 27385580 | 2953 | 346 | 0,510873 | 0,26055 | 0,78811 | 0,260991 | 0,06789 | 0,62112 | 0,00010566 | 0,009808 | *SEL1L3, SMIM20, RBPJ, CCKAR, TBC1D19, STIM2* |
| 1137 | 7 | 40261241 | 41415925 | 2138 | 263 | 0,693468 | 0,35027 | 1 | 0,480898 | 0,12269 | 1 | 0,00014932 | 0,01227724 | *SUGCT* |
| 2026 | 14 | 99474534 | 100786189 | 3058 | 348 | 0,426573 | 0,21952 | 0,65101 | 0,181965 | 0,04819 | 0,42381 | 0,00013928 | 0,01227724 | *BCL11B, SETD3, CCNK, CCDC85C, HHIPL1, CYP46A1, EML1, EVL, DE* |
| 2235 | 17 | 78438333 | 79333774 | 2884 | 277 | 0,486893 | 0,24878 | 0,75078 | 0,237065 | 0,06189 | 0,56367 | 0,00014951 | 0,01227724 | *NPTX1, RPTOR, CHMP6, AC127496.1, BAIAP2, AATK, AZI1, ENTHD2* |
| 1280 | 8 | 36641175 | 38803980 | 3449 | 368 | 0,535039 | 0,27273 | 0,85067 | 0,286266 | 0,07438 | 0,72364 | 0,00016314 | 0,01290022 | *KCNU1, ZNF703, RP11-863K10.7, ERLIN2, PROSC, GPR124, BRF2, R* |
| 724 | 4 | 135809543 | 137400031 | 3616 | 250 | 0,635019 | 0,31721 | 1 | 0,403249 | 0,10062 | 1 | 0,00017843 | 0,01360559 | |
| 1846 | 12 | 117091844 | 118256124 | 2874 | 315 | 0,539283 | 0,26634 | 0,92565 | 0,290826 | 0,07093 | 0,85683 | 0,00022078 | 0,01582334 | *C12orf49, RNFT2, HRK, FBXW8, TESC, FBXO21, NOS1, KSR2* |
| 2285 | 18 | 56832313 | 57635821 | 2356 | 296 | 0,638263 | 0,32557 | 1 | 0,407379 | 0,106 | 1 | 0,00022234 | 0,01582334 | *GRP, RAX, CPLX4, LMAN1, CCBE1, PMAIP1* |
| 1971 | 14 | 38722115 | 40343754 | 3531 | 236 | -0,985525 | -1 | -0,49881 | 0,97126 | 0,24881 | 1 | 0,00023667 | 0,01629969 | *CLEC14A, SEC23A, GEMIN2, TRAPPC6B, PNN, MIA2, RP11-407N17.* |
| 2495 | 22 | 50347997 | 51244236 | 2333 | 273 | 0,515439 | 0,2566 | 0,81853 | 0,265678 | 0,06585 | 0,66999 | 0,00025451 | 0,01698032 | *PIM3, IL17REL, TTLL8, MLC1, MOV10L1, PANX2, TRABD, SELO, TUE* |
| 2041 | 15 | 29141036 | 30604119 | 2692 | 297 | 0,484487 | 0,23351 | 0,76986 | 0,234728 | 0,05452 | 0,59269 | 0,00027962 | 0,01809063 | *APBA2, FAM189A1, NDNL2, TJP1, GOLGA8J, GOLGA8T* |
| 50 | 1 | 57529671 | 58387483 | 2145 | 240 | 0,655403 | 0,32467 | 1 | 0,429554 | 0,10541 | 1 | 0,00029758 | 0,01868634 | *DAB1* |
| 585 | 3 | 187939200 | 189451805 | 3586 | 355 | 0,558365 | 0,26739 | 0,91493 | 0,311772 | 0,0715 | 0,83709 | 0,00032391 | 0,01920943 | *LPP, TPRG1, TP63* |
| 903 | 5 | 153734495 | 154599750 | 1957 | 245 | 0,549576 | 0,26801 | 0,88189 | 0,302034 | 0,07183 | 0,77774 | 0,00031845 | 0,01920943 | *GALNT10, SAP30L, HAND1, LARP1, FAXDC2, CNOT8, GEMIN5, MRF* |
| 2025 | 14 | 98885323 | 99474533 | 2035 | 223 | 0,572247 | 0,27753 | 0,91634 | 0,327467 | 0,07702 | 0,83968 | 0,00034269 | 0,01977437 | *C14orf177* |
| 975 | 6 | 39785908 | 40778401 | 2846 | 265 | 0,63115 | 0,29626 | 1 | 0,39835 | 0,08777 | 1 | 0,00036409 | 0,02045617 | *DAAM2, MOCS1, LRFN2* |
| 896 | 5 | 145319097 | 147180171 | 3821 | 322 | 0,676109 | 0,32037 | 1 | 0,457123 | 0,10264 | 1 | 0,00038834 | 0,02072781 | *SH3RF2, PLAC8L1, LARS, RBM27, POU4F3, TCERG1, GPR151, PPP2I* |
| 1197 | 7 | 113339387 | 115321301 | 3112 | 278 | 0,501099 | 0,23898 | 0,80604 | 0,251101 | 0,05711 | 0,6497 | 0,00038286 | 0,02072781 | *PPP1R3A, FOXP2, MDFIC* |
| 2488 | 22 | 44409534 | 45170649 | 2607 | 362 | 0,545655 | 0,25429 | 0,89531 | 0,297739 | 0,06466 | 0,80158 | 0,00040351 | 0,02101194 | *PARVB, PARVG, KIAA1644, RP1-32I10.10, LDOC1L, PRR5, PRR5-ARI* |
| 280 | 2 | 75148104 | 75942397 | 2113 | 257 | 0,509901 | 0,241 | 0,87375 | 0,26 | 0,05808 | 0,76345 | 0,00042719 | 0,02166717 | *POLE4, TACR1, EVA1A, MRPL19, GCFC2* |
| 860 | 5 | 101575368 | 102903985 | 2970 | 170 | 0,620444 | 0,29653 | 1 | 0,38495 | 0,08793 | 1 | 0,00045669 | 0,02166717 | *SLCO4C1, SLCO6A1, PAM, GIN1, PPIP5K2, C5orf30, NUDT12* |
| 959 | 6 | 31250557 | 31320268 | 1223 | 34 | 0,605311 | 0,30716 | 0,92147 | 0,366401 | 0,09434 | 0,8491 | 0,00044866 | 0,02166717 | |
| 1988 | 14 | 56206431 | 57460781 | 3384 | 320 | 0,358751 | 0,16391 | 0,56095 | 0,128702 | 0,02687 | 0,31467 | 0,00044627 | 0,02166717 | *PELI2, TMEM260, RP11-1085N6.3, OTX2* |
| 474 | 3 | 61277588 | 62609818 | 3002 | 352 | 0,500574 | 0,2356 | 0,81306 | 0,250575 | 0,05551 | 0,66107 | 0,00048661 | 0,02258514 | *PTPRG, C3orf14, FEZF2, CADPS* |
| 967 | 6 | 32682214 | 32897998 | 1808 | 71 | 0,791301 | 0,40867 | 1 | 0,626157 | 0,16701 | 1 | 0,00053144 | 0,0241409 | *HLA-DQA2, HLA-DQB2, HLA-DOB, TAP2, PSMB8, PSMB9, TAP1* |
| 145 | 1 | 189904638 | 191419275 | 3398 | 198 | 0,581477 | 0,26839 | 0,98071 | 0,338116 | 0,07203 | 0,96179 | 0,00056515 | 0,0251374 | *BRINP3* |
| 2181 | 17 | 7264459 | 8554763 | 2751 | 302 | 0,361961 | 0,16304 | 0,57522 | 0,131016 | 0,02658 | 0,33087 | 0,0006041 | 0,0263215 | *TNK1, TMEM256-PLSCR3, C17orf61-PLSCR3, TMEM256, NLGN2, SI* |
| 577 | 3 | 176931164 | 178110322 | 2568 | 297 | 0,464407 | 0,21333 | 0,74866 | 0,215674 | 0,04551 | 0,56049 | 0,00063285 | 0,02692126 | *KCNMB2* |
| 692 | 4 | 102544804 | 104384534 | 3539 | 273 | -0,369591 | -0,58671 | -0,16329 | 0,136598 | 0,02666 | 0,34423 | 0,00064308 | 0,02692126 | *BANK1, SLC39A8, NFKB1, MANBA, UBE2D3, CISD2, SLC9B1, SLC9B* |
| 2378 | 20 | 7959829 | 9279550 | 3148 | 315 | 0,439269 | 0,19386 | 0,71901 | 0,192957 | 0,03758 | 0,51698 | 0,00067762 | 0,02782147 | *TMX4, PLCB1, PLCB4* |
| 901 | 5 | 151892618 | 152878627 | 2086 | 173 | 0,413409 | 0,1824 | 0,64319 | 0,170907 | 0,03327 | 0,41369 | 0,0007097 | 0,02858894 | *GRIA1* |
| 917 | 5 | 169506967 | 170898340 | 3284 | 334 | 0,409007 | 0,17676 | 0,66046 | 0,167287 | 0,03125 | 0,4362 | 0,00073625 | 0,02869363 | *DOCK2, FOXI1, C5orf58, LCP2, KCNIP1, KCNMB1, GABRP, RANBP1* |
| 950 | 6 | 25684630 | 26396200 | 1874 | 130 | 0,421898 | 0,18943 | 0,66594 | 0,177998 | 0,03588 | 0,44347 | 0,00075231 | 0,02869363 | *SCGN, HIST1H2AA, HIST1H2BA, SLC17A4, SLC17A1, SLC17A3, SLC1* |
| 1271 | 8 | 25859278 | 26679135 | 2713 | 251 | 0,521235 | 0,23441 | 0,85402 | 0,271686 | 0,05495 | 0,72935 | 0,00076516 | 0,02869363 | *EBF2, PPP2R2A, BNIP3L, PNMA2, DPYSL2, ADRA1A* |
| 1376 | 9 | 4025356 | 4596614 | 1872 | 298 | 0,650014 | 0,29234 | 1 | 0,422519 | 0,08546 | 1 | 0,00076657 | 0,02869363 | *GLIS3, SLC1A1, SPATA6L* |
| 1382 | 9 | 9170121 | 9920258 | 2411 | 277 | -0,718378 | -1 | -0,31058 | 0,516068 | 0,09646 | 1 | 0,0007795 | 0,02869363 | *PTPRD* |
| 450 | 3 | 29877023 | 31269697 | 3744 | 316 | -0,418125 | -0,68229 | -0,17864 | 0,174828 | 0,03191 | 0,46552 | 0,00079835 | 0,02888934 | *RBMS3, TGFBR2, GADL1* |
| 435 | 3 | 12859210 | 14312007 | 3800 | 349 | 0,495914 | 0,21497 | 0,84693 | 0,24593 | 0,04621 | 0,71729 | 0,00083253 | 0,02960269 | *CAND2, RPL32, IQSEC1, NUP210, HDAC11, FBLN2, WNT7A, TPRXL,* |
| 1053 | 6 | 123854858 | 125365054 | 3409 | 347 | 0,593736 | 0,25918 | 1 | 0,352523 | 0,06717 | 1 | 0,00084579 | 0,02960269 | *TRDN, NKAIN2, RNF217* |
| 73 | 1 | 84728348 | 86017267 | 3260 | 290 | -0,607525 | -1 | -0,26597 | 0,369087 | 0,07074 | 1 | 0,00089967 | 0,03098057 | *SAMD13, DNASE2B, RPF1, GNG5, CTBS, C1orf180, SSX2IP, LPAR3,* |
| 329 | 2 | 138073792 | 139505819 | 2970 | 228 | 0,504888 | 0,21435 | 0,82878 | 0,254912 | 0,04594 | 0,68688 | 0,0009231 | 0,03128297 | *THSD7B, HNMT, SPOPL, NXPH2* |
| 1847 | 12 | 118256125 | 119301061 | 2531 | 264 | 0,417373 | 0,17838 | 0,6775 | 0,1742 | 0,03182 | 0,45901 | 0,00095265 | 0,03177971 | *KSR2, RFC5, WSB2, VSIG10, PEBP1, TAOK3, SUDS3* |
| 78 | 1 | 89753965 | 90941549 | 2238 | 266 | 0,546955 | 0,22849 | 0,94827 | 0,29916 | 0,05221 | 0,89922 | 0,00106979 | 0,03507805 | *GBP6, LRRC8B, LRRC8C, RP11-302M6.4, LRRC8D, ZNF326* |
| 1400 | 9 | 23859375 | 24757729 | 1855 | 235 | 0,578223 | 0,23579 | 1 | 0,334342 | 0,05559 | 1 | 0,00108438 | 0,03507805 | *IZUMO3* |
| 1099 | 7 | 4139432 | 4938442 | 2682 | 319 | 0,442361 | 0,18652 | 0,73109 | 0,195683 | 0,03479 | 0,53449 | 0,00110167 | 0,03510545 | *SDK1, FOXK1, AP5Z1, RADIL, PAPOLB* |
| 848 | 5 | 86493764 | 87943482 | 2202 | 195 | 0,415036 | 0,16606 | 0,66963 | 0,172255 | 0,02758 | 0,4484 | 0,00120874 | 0,03792193 | *AC008394.1, RASA1, CCNH, TMEM161B* |
| 2280 | 18 | 51060987 | 52512523 | 3610 | 187 | 0,649694 | 0,27525 | 1 | 0,422102 | 0,07576 | 1 | 0,00122558 | 0,03792193 | *MBD2, POLI, STARD6, C18orf54, DYNAP, RAB27B* |
| 645 | 4 | 43962562 | 45186767 | 3531 | 210 | -0,469082 | -0,78273 | -0,20033 | 0,220038 | 0,04013 | 0,61266 | 0,00126171 | 0,03848216 | *KCTD8, YIPF7, GUF1, GNPDA2* |
| 2271 | 18 | 41425455 | 42974165 | 3034 | 299 | 0,560603 | 0,23243 | 0,97569 | 0,314276 | 0,05402 | 0,95197 | 0,00130432 | 0,03922145 | *SETBP1, SLC14A2* |
| 1277 | 8 | 32454963 | 33982537 | 2828 | 248 | 0,459394 | 0,18213 | 0,7636 | 0,211043 | 0,03317 | 0,58309 | 0,00133177 | 0,03949068 | *NRG1, FUT10, TTI2, MAK16, RNF122, DUSP26* |
| 518 | 3 | 112408514 | 113657665 | 3125 | 252 | 0,534341 | 0,21759 | 0,93726 | 0,28552 | 0,04735 | 0,87845 | 0,00137463 | 0,04020322 | *CD200R1L, CD200R1, GTPBP8, C3orf17, BOC, WDR52, SPICE1, SID* |

| | | | | | | | | | | | | | | |
|---|---|---|---|---|---|---|---|---|---|---|---|---|---|---|
| 466 | 3 | 51953969 | 54074844 | 3566 | 304 | -0,34015 | -0,55393 | -0,13122 | 0,115702 | 0,01722 | 0,30684 | 0,00145515 | 0,04198304 | *RRP9, PARP3, GPR62, PCBP4, RP11-155D18.14, ABHD14B, ABHD14* |
| 1992 | 14 | 60099483 | 61652816 | 2276 | 189 | 0,694188 | 0,2952 | 1 | 0,481897 | 0,08714 | 1 | 0,00149063 | 0,04243327 | *RTN1, LRRC9, PCNXL4, DHRS7, PPM1A, C14orf39, SIX6, SIX1, SIX4,* |
| 551 | 3 | 148638309 | 149998411 | 3408 | 315 | -0,865611 | -1 | -0,34609 | 0,749283 | 0,11978 | 1 | 0,00155585 | 0,0437071 | *GYG1, HLTF, HPS3, CP, TM4SF18, TM4SF1, TM4SF4, WWTR1, COM* |
| 244 | 2 | 40087419 | 40786176 | 1970 | 250 | 0,425577 | 0,17229 | 0,70356 | 0,181116 | 0,02969 | 0,49499 | 0,00160286 | 0,04427552 | *SLC8A1, AC007377.1* |
| 346 | 2 | 160200380 | 161432089 | 3029 | 208 | 0,423038 | 0,16887 | 0,69818 | 0,178962 | 0,02852 | 0,48745 | 0,00161756 | 0,04427552 | *BAZ2B, MARCH7, CD302, LY75, LY75-CD302, PLA2R1, ITGB6, RBMS* |
| 124 | 1 | 166458032 | 167163954 | 2039 | 185 | 0,42436 | 0,16685 | 0,72413 | 0,180081 | 0,02786 | 0,52437 | 0,00171229 | 0,04617354 | *POGK, TADA1, ILDR2, MAEL, GPA33, DUSP27* |
| 388 | 2 | 210842458 | 212520453 | 3511 | 287 | 0,513306 | 0,20841 | 0,93313 | 0,263483 | 0,04344 | 0,87072 | 0,00175145 | 0,04617354 | *UNC80, RPE, KANSL1L, ACADL, MYL1, LANCL1, CPS1, ERBB4* |
| 1023 | 6 | 91097025 | 92015279 | 2218 | 244 | 0,350467 | 0,13094 | 0,58168 | 0,122827 | 0,01715 | 0,33835 | 0,00176009 | 0,04617354 | *MAP3K7* |
| 1230 | 7 | 157199332 | 158031247 | 2678 | 315 | 0,541592 | 0,21936 | 1 | 0,293321 | 0,04823 | 1 | 0,00177341 | 0,04617354 | *DNAJB6, AC006372.1, PTPRN2, AC005481.5* |
| 1162 | 7 | 67474723 | 68569224 | 3770 | 292 | 0,425368 | 0,16321 | 0,71592 | 0,180938 | 0,02664 | 0,51255 | 0,00180794 | 0,04650544 | |
| 305 | 2 | 110097869 | 112644828 | 2709 | 297 | 0,618579 | 0,24344 | 1 | 0,382639 | 0,05926 | 1 | 0,00184789 | 0,0469672 | *SH3RF3, SEPT10, SOWAHC, RGPD5, LIMS3, MALL, NPHP1, LIMS3L,* |
| 1181 | 7 | 93691726 | 95174847 | 2434 | 258 | 0,541584 | 0,21653 | 0,94907 | 0,293313 | 0,04689 | 0,90074 | 0,00194022 | 0,04873376 | *COL1A2, CASD1, SGCE, PEG10, PPP1R9A, PON1, PON3, PON2, ASB* |
| 265 | 2 | 57433654 | 57952945 | 1647 | 135 | 0,577756 | 0,22141 | 1 | 0,333802 | 0,04916 | 1 | 0,00205963 | 0,04902126 | |
| 650 | 4 | 53879572 | 54659778 | 1802 | 176 | 0,559627 | 0,23084 | 0,98642 | 0,313183 | 0,05328 | 0,97303 | 0,00206647 | 0,04902126 | *SCFD2, FIP1L1, LNX1* |
| 852 | 5 | 91956906 | 93814604 | 2431 | 286 | 0,348362 | 0,13994 | 0,57722 | 0,121356 | 0,01958 | 0,33319 | 0,00201992 | 0,04902126 | *NR2F1, FAM172A, POU5F2, KIAA0825* |
| 1588 | 10 | 113117616 | 114255954 | 2660 | 251 | 0,44637 | 0,1796 | 0,74145 | 0,199246 | 0,03225 | 0,54975 | 0,00200887 | 0,04902126 | *GPAM, TECTB, ACSL5, ZDHHC6, VTI1A* |
| 2311 | 19 | 3893910 | 4741718 | 1959 | 314 | 0,51912 | 0,20217 | 1 | 0,269485 | 0,04087 | 1 | 0,00204948 | 0,04902126 | *ATCAY, NMRK2, DAPK3, EEF2, PIAS4, AC016586.1, ZBTB7A, MAP2K* |
| 212 | 2 | 7966710 | 8978798 | 2186 | 319 | 0,416471 | 0,1612 | 0,70136 | 0,173448 | 0,02598 | 0,49191 | 0,00211429 | 0,0496045 | *ID2, KIDINS220* |
| 153 | 1 | 199263390 | 200134005 | 2537 | 190 | 0,771825 | 0,32246 | 1 | 0,595714 | 0,10398 | 1 | 0,00214696 | 0,04982347 | *NR5A2* |
| **SCZ - TUD** | | | | | | | | | | | | | | |
| 63 | 1 | 73992171 | 75132092 | 2018 | 221 | 0,538366 | 0,35517 | 0,73337 | 0,289838 | 0,12615 | 0,53784 | 1,4071E-07 | 0,00029436 | *NEGR1* |
| 62 | 1 | 72513120 | 73992170 | 2829 | 164 | 0,55585 | 0,33893 | 0,81593 | 0,308969 | 0,11487 | 0,66574 | 6,9917E-06 | 0,00731328 | *LRRIQ3, FPGT, FPGT-TNNI3K, TNNI3K, LRRC53, C1orf173* |
| 134 | 1 | 177746473 | 178915113 | 2339 | 209 | 0,583029 | 0,32442 | 0,88377 | 0,339923 | 0,10525 | 0,78105 | 5,9319E-05 | 0,04136505 | *SEC16B, RASAL2, TEX35, C1orf220, C1ORF220, RALGPS2, ANGPTL1* |
| 796 | 5 | 22149043 | 23246745 | 2481 | 220 | 0,638386 | 0,34855 | 1 | 0,407536 | 0,12149 | 1 | 0,00010123 | 0,0423538 | *CDH12* |
| 849 | 5 | 87943483 | 89584466 | 2123 | 291 | 0,442776 | 0,22965 | 0,66789 | 0,19605 | 0,05274 | 0,44608 | 8,4894E-05 | 0,0423538 | *MEF2C* |
| **SCZ - AF** | | | | | | | | | | | | | | |
| 1719 | 11 | 112755447 | 113889019 | 1489 | 150 | 0,682012 | 0,50675 | 0,86192 | 0,465141 | 0,25679 | 0,74291 | 1,4537E-10 | 1,5903E-07 | *NCAM1, TTC12, ANKK1, DRD2, TMPRSS5, ZW10, CLDN25, USP28, H* |
| 692 | 4 | 102544804 | 104384534 | 1645 | 135 | -0,883931 | -1 | -0,62777 | 0,781335 | 0,39409 | 1 | 4,0102E-08 | 2,1936E-05 | *BANK1, SLC39A8, NFKB1, MANBA, UBE2D3, CISD2, SLC9B1, SLC9B2* |
| 266 | 2 | 57952946 | 59251996 | 1245 | 157 | 0,885333 | 0,6062 | 1 | 0,783815 | 0,36748 | 1 | 1,4923E-07 | 5,4418E-05 | *VRK2, FANCL* |
| 425 | 3 | 3128153 | 4109393 | 1918 | 216 | 1 | 0,62361 | 1 | 1 | 0,38889 | 1 | 1,722E-06 | 0,00047096 | *IL5RA, TRNT1, CRBN, SUMF1, LRRN1* |
| 1466 | 9 | 121008530 | 122677719 | 1971 | 174 | 0,739525 | 0,46753 | 1 | 0,546897 | 0,21859 | 1 | 2,6226E-06 | 0,00057382 | *BRINP1, RP11-295D22.1* |
| 1498 | 10 | 10969482 | 11856924 | 1223 | 189 | 1 | 0,59922 | 1 | 1 | 0,35906 | 1 | 5,8929E-06 | 0,00107448 | *CELF2, USP6NL, ECHDC3* |
| 1375 | 9 | 3079046 | 4025355 | 1096 | 178 | 0,932916 | 0,57281 | 1 | 0,870333 | 0,32811 | 1 | 1,0279E-05 | 0,00160646 | *RFX3, AL365202.1, GLIS3* |
| 40 | 1 | 43512670 | 45167235 | 1799 | 188 | 0,960438 | 0,56743 | 1 | 0,922441 | 0,32197 | 1 | 1,8887E-05 | 0,0019933 | *FAM183A, EBNA1BP2, WDR65, TMEM125, C1orf210, TIE1, MPL, CD* |
| 63 | 1 | 73992171 | 75132092 | 1077 | 94 | 0,829016 | 0,51021 | 1 | 0,687268 | 0,26032 | 1 | 1,9093E-05 | 0,0019933 | *LRRIQ3, FPGT, FPGT-TNNI3K, TNNI3K, LRRC53, C1orf173* |
| 79 | 1 | 90941550 | 91870059 | 1134 | 117 | 1 | 0,54155 | 1 | 1 | 0,29327 | 1 | 1,5194E-05 | 0,0019933 | *BARHL2, ZNF644, HFM1* |
| 1794 | 12 | 56987106 | 58748139 | 1657 | 145 | 0,603482 | 0,35656 | 0,88845 | 0,36419 | 0,12714 | 0,78934 | 2,0042E-05 | 0,0019933 | *BAZ2A, ATP5B, PTGES3, NACA, PRIM1, HSD17B6, SDR9C7, RDH16,* |
| 2141 | 16 | 61092845 | 62607090 | 1402 | 129 | 1 | 0,50867 | 1 | 1 | 0,25874 | 1 | 2,6565E-05 | 0,00242187 | *CDH8* |
| 513 | 3 | 107736520 | 108638900 | 1272 | 109 | 1 | 0,50789 | 1 | 1 | 0,25796 | 1 | 3,386E-05 | 0,00271133 | *CD47, IFT57, HHLA2, MYH15, KIAA1524, DZIP3, RETNLB, TRAT1, GU* |
| 1472 | 9 | 128785783 | 129617771 | 926 | 184 | 0,7122 | 0,40709 | 1 | 0,507228 | 0,16572 | 1 | 3,4697E-05 | 0,00271133 | *MVB12B, LMX1B, ZBTB43* |
| 2402 | 20 | 37253658 | 38427594 | 1764 | 151 | 1 | 0,52101 | 1 | 1 | 0,27145 | 1 | 3,9341E-05 | 0,00286928 | *ARHGAP40, SLC32A1, ACTR5, PPP1R16B, FAM83D, DHX35* |
| 62 | 1 | 72513120 | 73992170 | 1710 | 61 | 0,861765 | 0,53728 | 1 | 0,74264 | 0,28867 | 1 | 5,0927E-05 | 0,00341364 | *NEGR1* |
| 2090 | 15 | 90632719 | 91593623 | 1300 | 107 | 0,980645 | 0,5725 | 1 | 0,961666 | 0,32775 | 1 | 5,3046E-05 | 0,00341364 | *IDH2, SEMA4B, CIB1, GDPGP1, TTLL13, RP11-697E2.6, NGRN, GAB* |
| 2029 | 14 | 103617473 | 104439106 | 1234 | 84 | 0,940444 | 0,56098 | 1 | 0,884434 | 0,3147 | 1 | 6,1198E-05 | 0,00371945 | *EIF5, MARK3, CKB, TRMT61A, BAG5, KLC1, RP11-73M18.2, APOPT1* |
| 2281 | 18 | 52512524 | 53762996 | 1285 | 142 | 0,601104 | 0,32652 | 0,93325 | 0,361326 | 0,10662 | 0,87096 | 7,0089E-05 | 0,00403567 | *RAB27B, CCDC68, TCF4* |
| 1667 | 11 | 56623495 | 57675986 | 856 | 124 | 0,792617 | 0,442 | 1 | 0,628242 | 0,19537 | 1 | 8,1008E-05 | 0,00427067 | *OR5AK2, LRRC55, APLNR, TNKS1BP1, SSRP1, P2RX3, PRG3, PRG2, P* |
| 2242 | 18 | 3722829 | 4603961 | 1153 | 194 | 1 | 0,52807 | 1 | 1 | 0,27886 | 1 | 8,1978E-05 | 0,00427067 | *DLGAP1* |
| 2172 | 16 | 89106305 | 90292787 | 1601 | 170 | 1 | 0,47822 | 1 | 1 | 0,2287 | 1 | 0,00017463 | 0,00868387 | *ACSF3, CDH15, SLC22A31, ZNF778, ANKRD11, AC137932.1, SPG7, I* |
| 179 | 1 | 230227248 | 231287764 | 1863 | 219 | 0,983445 | 0,46824 | 1 | 0,967164 | 0,21924 | 1 | 0,00035739 | 0,0150812 | *GALNT2, PGBD5, COG2, AGT, CAPN9, C1orf198, TTC13, ARV1, FAM* |
| 912 | 5 | 162386028 | 163781268 | 1793 | 176 | 1 | 0,40422 | 1 | 1 | 0,1634 | 1 | 0,00033176 | 0,0150812 | *CCNG1, AC112205.1, NUDCD2, HMMR, MAT2B* |
| 1197 | 7 | 113339387 | 115321301 | 1738 | 157 | 0,880478 | 0,44543 | 1 | 0,775241 | 0,1984 | 1 | 0,00039978 | 0,0150812 | *PPP1R3A, FOXP2, MDFIC* |
| 1360 | 8 | 135313674 | 136220342 | 1145 | 140 | 1 | 0,45025 | 1 | 1 | 0,20273 | 1 | 0,00034411 | 0,0150812 | *ZFAT* |
| 1476 | 9 | 132999453 | 134141936 | 1250 | 209 | -0,807913 | -1 | -0,39151 | 0,652723 | 0,15328 | 1 | 0,00038231 | 0,0150812 | *NCS1, HMCN2, AL354898.1, ASS1, FUBP3, PRDM12, EXOSC2, ABL1* |
| 1884 | 13 | 41003944 | 42037054 | 1020 | 105 | 1 | 0,45682 | 1 | 1 | 0,20868 | 1 | 0,00037682 | 0,0150812 | *AL133318.1, FOXO1, MRPS31, SLC25A15, ELF1, WBP4, KBTBD6, KB* |
| 2305 | 18 | 77148138 | 78017072 | 1288 | 150 | 0,625976 | 0,30616 | 1 | 0,391846 | 0,09374 | 1 | 0,00039484 | 0,0150812 | *NFATC1, AC018445.1, CTDP1, KCNG2, PQLC1, HSBP1L1, TXNL4A, R* |
| 38 | 1 | 41093876 | 41977966 | 1318 | 149 | 0,686425 | 0,3224 | 1 | 0,471179 | 0,10394 | 1 | 0,00049556 | 0,0164287 | *RIMS3, NFYC, KCNQ4, CITED4, CTPS1, SLFNL1, SCMH1, FOXO6, EDI* |
| 969 | 6 | 33194976 | 33864262 | 1370 | 128 | 0,663366 | 0,31818 | 1 | 0,440055 | 0,10124 | 1 | 0,00045571 | 0,0164287 | *VPS52, RPS18, B3GALT4, WDR46, PFDN6, RGL2, TAPBP, ZBTB22, D* |
| 2096 | 15 | 96864279 | 98025684 | 1203 | 200 | 0,655086 | 0,30917 | 1 | 0,429138 | 0,09558 | 1 | 0,00048972 | 0,0164287 | *NR2F2, AC087477.1, SPATA8* |
| 2470 | 22 | 25282437 | 26166934 | 796 | 111 | 0,741511 | 0,37513 | 1 | 0,549838 | 0,14072 | 1 | 0,00048103 | 0,0164287 | *SGSM1, TMEM211, KIAA1671, CRYBB3, CRYBB2, LRP5L, ADRBK2, M* |

| | | | | | | | | | | | | | | |
|---|---|---|---|---|---|---|---|---|---|---|---|---|---|---|
| 570 | 3 | 168972663 | 170174578 | 1418 | 147 | 0,85189 | 0,41546 | 1 | 0,725716 | 0,17261 | 1 | 0,00058425 | 0,01756112 | *MECOM, ACTRT3, MYNN, LRRC34, LRRIQ4, LRRC31, SAMD7, SEC62* |
| 701 | 4 | 113916003 | 115306149 | 1706 | 163 | 0,805622 | 0,39863 | 1 | 0,649027 | 0,1589 | 1 | 0,00057424 | 0,01756112 | *ANK2, CAMK2D, ARSJ* |
| 1027 | 6 | 95176338 | 96480153 | 1721 | 91 | 1 | 0,47202 | 1 | 1 | 0,22307 | 1 | 0,00059393 | 0,01756112 | *MANEA, FUT9* |
| 1919 | 13 | 79489046 | 80778339 | 1218 | 150 | 0,891449 | 0,43034 | 1 | 0,794682 | 0,1852 | 1 | 0,00058434 | 0,01756112 | *RBM26, NDFIP2* |
| 1367 | 8 | 142611890 | 143611461 | 1340 | 146 | 0,749142 | 0,35768 | 1 | 0,561214 | 0,12794 | 1 | 0,00061719 | 0,0177686 | *TSNARE1, BAI1* |
| 2386 | 20 | 15962577 | 16661516 | 1339 | 141 | 0,818474 | 0,39469 | 1 | 0,6699 | 0,15578 | 1 | 0,00065352 | 0,01833213 | *MACROD2, KIF16B* |
| 2235 | 17 | 78438333 | 79333774 | 1498 | 160 | 0,834658 | 0,39248 | 1 | 0,696654 | 0,15404 | 1 | 0,00068173 | 0,01835716 | *NPTX1, RPTOR, CHMP6, AC127496.1, BAIAP2, AATK, AZI1, ENTHD2* |
| 2283 | 18 | 54957868 | 55809315 | 1512 | 157 | -1 | -1 | -0,36831 | 1 | 0,13606 | 1 | 0,00068797 | 0,01835716 | *ST8SIA3, ONECUT2, FECH, NARS, ATP8B1, NEDD4L* |
| 624 | 4 | 25832019 | 27385580 | 1397 | 218 | 0,696135 | 0,31399 | 1 | 0,484605 | 0,09859 | 1 | 0,00072612 | 0,01891372 | *SEL1L3, SMIM20, RBPJ, CCKAR, TBC1D19, STIM2* |
| 2026 | 14 | 99474534 | 100786189 | 1567 | 211 | 0,477152 | 0,21131 | 0,77455 | 0,227674 | 0,04465 | 0,59993 | 0,00075478 | 0,01920293 | *BCL11B, SETD3, CCNK, CCDC85C, HHIPL1, CYP46A1, EML1, EVL, DE* |
| 2079 | 15 | 78514102 | 79292536 | 459 | 91 | 0,880709 | 0,4252 | 1 | 0,775648 | 0,1808 | 1 | 0,00096103 | 0,02389478 | *ACSBG1, DNAJA4, WDR61, CRABP1, IREB2, HYKK, AC027228.1, PSM* |
| 347 | 2 | 161432090 | 163607635 | 2068 | 121 | 0,841713 | 0,39451 | 1 | 0,708481 | 0,15564 | 1 | 0,0010003 | 0,02430939 | *TANK, PSMD14, TBR1, SLC4A10, DPP4, GCG, FAP, IFIH1, GCA, KCN* |
| 580 | 3 | 181424324 | 183203155 | 1609 | 157 | 0,806026 | 0,36345 | 1 | 0,649678 | 0,13209 | 1 | 0,00102215 | 0,02430939 | *SOX2, ATP11B, DCUN1D1, MCCC1, LAMP3, MCF2L2, B3GNT5* |
| 449 | 3 | 28649811 | 29877022 | 1969 | 225 | 0,521045 | 0,21787 | 0,87967 | 0,271488 | 0,04747 | 0,77381 | 0,00104715 | 0,02437409 | *RBMS3* |
| 1988 | 14 | 56206431 | 57460781 | 1570 | 180 | 0,552078 | 0,2393 | 0,95774 | 0,304791 | 0,05727 | 0,91726 | 0,00113943 | 0,02596951 | *PELI2, TMEM260, RP11-1085N6.3, OTX2* |
| 56 | 1 | 65894185 | 66778015 | 1206 | 83 | 0,648081 | 0,30563 | 1 | 0,42001 | 0,09341 | 1 | 0,001278 | 0,0276797 | *LEPR, LEPROT, PDE4B* |
| 1951 | 13 | 112319065 | 113573118 | 1556 | 207 | 0,681211 | 0,29717 | 1 | 0,464048 | 0,08831 | 1 | 0,00125251 | 0,0276797 | *RP11-65D24.2, SOX1, SPACA7, TUBGCP3, C13orf35, ATP11A, MCF2* |
| 1963 | 14 | 29029225 | 30831154 | 1616 | 193 | 0,558191 | 0,24872 | 1 | 0,311577 | 0,06186 | 1 | 0,00129037 | 0,0276797 | *FOXG1, C14orf23, PRKD1* |
| 1473 | 9 | 129617772 | 130902727 | 1128 | 113 | 0,492861 | 0,20651 | 0,79675 | 0,242912 | 0,04265 | 0,63481 | 0,00137979 | 0,02902866 | *ZBTB34, RALGPS1, ANGPTL2, GARNL3, SLC2A8, ZNF79, RPL12, LRS* |
| 1809 | 12 | 74080748 | 75563968 | 2244 | 114 | 0,877273 | 0,38851 | 1 | 0,769607 | 0,15108 | 1 | 0,00149439 | 0,03084647 | *ATXN7L3B, KCNC2* |
| 96 | 1 | 109271450 | 110224230 | 1217 | 141 | 1 | 0,30608 | 1 | 1 | 0,0968 | 1 | 0,00176352 | 0,03370595 | *FNDC7, STXBP3, AKNAD1, GPSM2, CLCC1, WDR47, TAF13, TMEM1* |
| 638 | 4 | 37880861 | 38984838 | 1769 | 212 | 0,691008 | 0,28396 | 1 | 0,477492 | 0,08063 | 1 | 0,00181065 | 0,03370595 | *TBC1D1, PTTG2, AC021860.1, KLF3, TLR10, TLR1, TLR6, FAM114A1,* |
| 1026 | 6 | 93696826 | 95176337 | 2146 | 166 | 0,990389 | 0,42529 | 1 | 0,980871 | 0,18087 | 1 | 0,00167155 | 0,03370595 | *EPHA7* |
| 1723 | 11 | 117295642 | 118901213 | 2099 | 233 | 0,549809 | 0,21379 | 1 | 0,30229 | 0,04607 | 1 | 0,0017422 | 0,03370595 | *DSCAML1, FXYD2, FXYD6-FXYD2, FXYD6, TMPRSS13, IL10RA, TMP* |
| 1849 | 12 | 120567741 | 121817509 | 1684 | 143 | 0,830876 | 0,35825 | 1 | 0,690354 | 0,12834 | 1 | 0,00181778 | 0,03370595 | *GCN1L1, RPLP0, PXN, SIRT4, PLA2G1B, MSI1, COX6A1, AL021546.6* |
| 2489 | 22 | 45170650 | 46351048 | 1686 | 213 | 0,649589 | 0,26209 | 1 | 0,421966 | 0,06869 | 1 | 0,00178643 | 0,03370595 | *PRR5-ARHGAP8, ARHGAP8, PHF21B, NUP50, KIAA0930, UPK3A, FA* |
| 7 | 1 | 4773404 | 5362404 | 945 | 147 | 0,752298 | 0,31979 | 1 | 0,565952 | 0,10226 | 1 | 0,00195943 | 0,03572694 | *AJAP1* |
| 1474 | 9 | 130902728 | 132193798 | 869 | 98 | 0,772458 | 0,33404 | 1 | 0,596691 | 0,11158 | 1 | 0,00201888 | 0,03618846 | *LCN2, C9orf16, CIZ1, DNM1, GOLGA2, SWI5, AL359091.2, TRUB2,* |
| 2095 | 15 | 95662426 | 96864278 | 1459 | 217 | 0,455128 | 0,18148 | 0,76864 | 0,207141 | 0,03294 | 0,59081 | 0,0020509 | 0,03618846 | *AC016251.1* |
| 1264 | 8 | 19488889 | 20135628 | 1146 | 158 | -0,799446 | -1 | -0,32625 | 0,639114 | 0,10644 | 1 | 0,00233424 | 0,04036467 | *CSGALNACT1, INTS10, LPL, SLC18A1, ATP6V1B2, LZTS1* |
| 2233 | 17 | 76596288 | 77412786 | 1080 | 196 | 0,648931 | 0,26093 | 1 | 0,421112 | 0,06808 | 1 | 0,00236137 | 0,04036467 | *CYTH1, USP36, TIMP2, DDC8, LGALS3BP, CANT1, C1QTNF1-AS1, C1* |
| 434 | 3 | 11997659 | 12859209 | 1507 | 108 | 1 | 0,39944 | 1 | 1 | 0,16056 | 1 | 0,00245711 | 0,04135505 | *TIMP4, PPARG, TSEN2, C3orf83, MKRN2, RAF1, TMEM40, CAND2* |
| 620 | 4 | 21416381 | 22835185 | 2151 | 148 | 0,988628 | 0,39742 | 1 | 0,977386 | 0,15827 | 1 | 0,00267398 | 0,04432324 | *KCNIP4, GPR125* |
| 2072 | 15 | 69089816 | 70767983 | 1665 | 233 | 0,440227 | 0,15969 | 0,75905 | 0,1938 | 0,0255 | 0,57615 | 0,00275863 | 0,0450439 | *ANP32A, SPESP1, NOX5, GLCE, PAQR5, KIF23, RPLP1, TLE3* |
| 2083 | 15 | 82191078 | 84209760 | 1695 | 185 | 0,64461 | 0,24856 | 1 | 0,415522 | 0,06178 | 1 | 0,00299766 | 0,04785267 | *MEX3B, RP11-597K23.2, EFTUD1, FAM154B, GOLGA6L10, GOLGA6* |
| 2243 | 18 | 4603962 | 5575812 | 1258 | 166 | 0,524756 | 0,18845 | 0,92303 | 0,275369 | 0,03551 | 0,85199 | 0,00301813 | 0,04785267 | *C18orf42, ZBTB14, EPB41L3* |

SCZ: Schizophrenia; AF: Addiction Factor; CanUD: Cannabis Use Disorder; OUD: Opioid Use Disorder; TUD: Tobacco Use Disorder

**Table S3. Comparisons of the Akaike Information Criterion (AIC) against minimum- and maximum-overlap reference bivariate MiXeR models.**

| Model | best_vs_min_AIC | best_vs_max_AIC |
|---|---|---|
| SCZ-CanUD | 0.867 | 2.426 |
| SCZ-OUD | -0.406 | 0.385 |
| SCZ-PAU | 2.540 | 4.110 |
| SCZ-TUD | 7.727 | 0.0356 |

**Table S4. Genomic structural equation model among schizophrenia and substance use disorders.**

| Exploratory factor analyses (EFA) | | | | Confirmatory factor analyes (CFA) | | | | | | |
|---|---|---|---|---|---|---|---|---|---|---|
| **Trait** | **MR** | **h2** | **u2** | **Estimate** | **SE** | **Z** | **P-value** | **Std.all** | **$R^2$** | **Residual** |
| SCZ | 0,37 | 0,14 | 0,86 | 0,476 | 0,005 | 92,743 | < 0,004 | 0,476 | 22,66% | 0,844 |
| CanUD | 0,90 | 0,8 | 0,2 | 1,069 | 0,004 | 241,255 | < 0,001 | 0,884 | 78,14% | 0,217 |
| OUD | 0,81 | 0,66 | 0,34 | 0,99 | 0,004 | 222,091 | < 0,002 | 0,816 | 66,58% | 0,329 |
| PAU | 0,84 | 0,71 | 0,29 | 1 | 0 | NA | NA | 0,827 | 68,36% | 0,316 |
| TUD | 0,63 | 0,39 | 0,61 | 0,784 | 0,005 | 164,118 | < 0,003 | 0,784 | 61,47% | 0,578 |

SCZ: Schizophrenia; CanUD: Cannabis Use Disorder; OUD: Opioid Use Disorder; PAU: Problematic Alcohol Use; TUD: Tobacco Use

**Table S5. Association of substance use disorder- and addiction factor-PRSs with schizophrenia diagnosis.**

| Trait-informed PRS | PsyCourse cohort | | | | | | CIBERSAM cohort | | | | | |
|---|---|---|---|---|---|---|---|---|---|---|---|---|
| | Model variables | Beta ± SE | P-value | Nagelkerke R2* | OR (95% IC) | FDR** | Model variables | Beta ± SE | P-value | Nagelkerke R2* | OR (95% IC) | FDR** |
| AF | Intercept | 1,25 ± 0,25 | 8,37E-07 | | | | Intercept | 1,69 ± 0,16 | 1,20E-25 | | | |
| | AF-PRS | -0,01 ± 0,07 | 8,74E-01 | 3,66469E-05 | 0.99 (0.86-1.14) | 8,74E-01 | AF-PRS | 0,05 ± 0,04 | 2,10E-01 | 0,000725015 | 1.05 (0.97-1.14) | 2,62E-01 |
| | Sex | 0,95 ± 0,14 | 1,21E-11 | | | | Sex | -0,47 ± 0,08 | 9,62E-09 | | | |
| | Center | -0,09 ± 0,01 | 1,85E-09 | | | | Center | -0,12 ± 0,02 | 2,10E-09 | | | |
| | PC1 | -1,59 ± 2,42 | 5,10E-01 | | | | PC1 | -36,51 ± 3,2 | 4,37E-30 | | | |
| | PC2 | 0,71 ± 2,07 | 7,32E-01 | | | | PC2 | 31,37 ± 2,8 | 3,99E-29 | | | |
| | PC3 | -0,66 ± 2,09 | 7,53E-01 | | | | PC3 | 7,35 ± 2,38 | 2,02E-03 | | | |
| | PC4 | 2,31 ± 4,39 | 5,98E-01 | | | | PC4 | -4,78 ± 3,23 | 1,39E-01 | | | |
| | PC5 | -4,95 ± 2,99 | 9,76E-02 | | | | PC5 | 0,5 ± 2,39 | 8,34E-01 | | | |
| | PC6 | -8,39 ± 4,61 | 6,86E-02 | | | | PC6 | 1,67 ± 2,38 | 4,83E-01 | | | |
| | PC7 | -1,65 ± 2,82 | 5,58E-01 | | | | PC7 | 6,74 ± 2,36 | 4,27E-03 | | | |
| | PC8 | 5,13 ± 5 | 3,05E-01 | | | | PC8 | 4,42 ± 2,31 | 5,54E-02 | | | |
| | PC9 | 4,07 ± 4,21 | 3,34E-01 | | | | PC9 | 6,46 ± 2,34 | 5,75E-03 | | | |
| | PC10 | -0,78 ± 2,31 | 7,35E-01 | | | | PC10 | -6,32 ± 2,29 | 5,89E-03 | | | |
| CanUD | Intercept | 1,32 ± 0,25 | 2,24E-07 | | | | Intercept | 1,7 ± 0,16 | 6,46E-26 | | | |
| | CanUD-PRS | 0,27 ± 0,07 | 1,14E-04 | 0,0220662 | 1.32 (1.14-1.51) | 5,70E-04 | CanUD-PRS | 0,12 ± 0,04 | 2,03E-03 | | | |
| | Sex | 0,99 ± 0,14 | 4,56E-12 | | | | Sex | -0,46 ± 0,08 | 1,56E-08 | 0,0044032 | 1.13 (1.05-1.23) | 5,07E-03 |
| | Center | -0,09 ± 0,01 | 2,26E-10 | | | | Center | -0,12 ± 0,02 | 6,87E-10 | | | |
| | PC1 | -1,79 ± 2,5 | 4,75E-01 | | | | PC1 | -36,34 ± 3,2 | 7,15E-30 | | | |
| | PC2 | 1,22 ± 2,1 | 5,59E-01 | | | | PC2 | 30,56 ± 2,81 | 1,35E-27 | | | |
| | PC3 | -0,71 ± 2,09 | 7,36E-01 | | | | PC3 | 7,59 ± 2,39 | 1,48E-03 | | | |
| | PC4 | 2,62 ± 4,42 | 5,53E-01 | | | | PC4 | -4,77 ± 3,23 | 1,40E-01 | | | |
| | PC5 | -5,06 ± 3 | 9,16E-02 | | | | PC5 | 0,36 ± 2,39 | 8,81E-01 | | | |
| | PC6 | -9,04 ± 4,85 | 6,23E-02 | | | | PC6 | 1,77 ± 2,38 | 4,57E-01 | | | |
| | PC7 | -1,24 ± 2,9 | 6,69E-01 | | | | PC7 | 6,49 ± 2,36 | 6,01E-03 | | | |
| | PC8 | 5,3 ± 5,2 | 3,08E-01 | | | | PC8 | 4,35 ± 2,31 | 5,96E-02 | | | |
| | PC9 | 4,08 ± 4,39 | 3,52E-01 | | | | PC9 | 6,68 ± 2,34 | 4,34E-03 | | | |
| | PC10 | -1,52 ± 2,29 | 5,08E-01 | | | | PC10 | -6,4 ± 2,3 | 5,33E-03 | | | |
| OUD | Intercept | 1,3 ± 0,25 | 2,84E-07 | | | | Intercept | 1,71 ± 0,16 | 4,83E-26 | | | |
| | OUD-PRS | 0,15 ± 0,07 | 3,41E-02 | 0,00658116 | 1.16 (1.01-1.33) | 5,69E-02 | OUD-PRS | 0,1 ± 0,04 | 1,14E-02 | 0,00295694 | 1.11 (1.02-1.20) | 1,90E-02 |
| | Sex | 0,95 ± 0,14 | 1,56E-11 | | | | Sex | -0,47 ± 0,08 | 1,13E-08 | | | |
| | Center | -0,09 ± 0,01 | 4,42E-10 | | | | Center | -0,12 ± 0,02 | 7,07E-10 | | | |
| | PC1 | -1,92 ± 2,45 | 4,34E-01 | | | | PC1 | -36,26 ± 3,2 | 9,61E-30 | | | |
| | PC2 | 0,89 ± 2,08 | 6,68E-01 | | | | PC2 | 31,03 ± 2,8 | 1,67E-28 | | | |
| | PC3 | -0,74 ± 2,1 | 7,24E-01 | | | | PC3 | 7,41 ± 2,39 | 1,90E-03 | | | |
| | PC4 | 2,58 ± 4,62 | 5,77E-01 | | | | PC4 | -4,43 ± 3,22 | 1,70E-01 | | | |
| | PC5 | -5,03 ± 3,04 | 9,80E-02 | | | | PC5 | 0,38 ± 2,4 | 8,74E-01 | | | |
| | PC6 | -8,69 ± 4,85 | 7,30E-02 | | | | PC6 | 1,66 ± 2,39 | 4,88E-01 | | | |
| | PC7 | -1,36 ± 2,86 | 6,34E-01 | | | | PC7 | 6,54 ± 2,36 | 5,60E-03 | | | |
| | PC8 | 5,2 ± 5,19 | 3,16E-01 | | | | PC8 | 4,4 ± 2,31 | 5,63E-02 | | | |
| | PC9 | 4,07 ± 4,34 | 3,48E-01 | | | | PC9 | 6,58 ± 2,34 | 4,93E-03 | | | |
| | PC10 | -0,94 ± 2,31 | 6,84E-01 | | | | PC10 | -6,08 ± 2,3 | 8,08E-03 | | | |
| | Intercept | 1,3 ± 0,25 | 2,66E-07 | | | | Intercept | 1,7 ± 0,16 | 9,82E-26 | | | |
| | PAU-PRS | 0,21 ± 0,07 | 2,43E-03 | 0,0135763 | 1.24 (1.08-1.42) | 6,07E-03 | PAU-PRS | 0,2 ± 0,04 | 5,44E-07 | 0,0116823 | 1.23 (1.13-1.33) | 2,72E-06 |
| | Sex | 0,97 ± 0,14 | 9,19E-12 | | | | Sex | -0,46 ± 0,08 | 1,92E-08 | | | |

| | | | | | | | | | | | | |
|---|---|---|---|---|---|---|---|---|---|---|---|---|
| | Center | -0,09 ± 0,01 | 3,61E-10 | | | | Center | -0,12 ± 0,02 | 8,26E-10 | | | |
| | PC1 | -1,66 ± 2,44 | 4,95E-01 | | | | PC1 | -36,57 ± 3,2 | 3,47E-30 | | | |
| | PC2 | 0,72 ± 2,08 | 7,30E-01 | | | | PC2 | 30,44 ± 2,81 | 2,65E-27 | | | |
| PAU | PC3 | -0,63 ± 2,1 | 7,63E-01 | | | | PC3 | 7,25 ± 2,39 | 2,42E-03 | | | |
| | PC4 | 3,07 ± 4,47 | 4,91E-01 | | | | PC4 | -4,52 ± 3,24 | 1,63E-01 | | | |
| | PC5 | -4,76 ± 2,99 | 1,11E-01 | | | | PC5 | 0,31 ± 2,39 | 8,96E-01 | | | |
| | PC6 | -8,43 ± 4,65 | 6,97E-02 | | | | PC6 | 1,34 ± 2,38 | 5,73E-01 | | | |
| | PC7 | -1,8 ± 2,83 | 5,25E-01 | | | | PC7 | 6,17 ± 2,37 | 9,19E-03 | | | |
| | PC8 | 5,15 ± 5,03 | 3,06E-01 | | | | PC8 | 4,29 ± 2,31 | 6,35E-02 | | | |
| | PC9 | 3,96 ± 4,24 | 3,50E-01 | | | | PC9 | 6,34 ± 2,35 | 6,88E-03 | | | |
| | PC10 | -1,26 ± 2,3 | 5,84E-01 | | | | PC10 | -6,4 ± 2,3 | 5,48E-03 | | | |
| | Intercept | 1,24 ± 0,25 | 8,79E-07 | | | | Intercept | 1,69 ± 0,16 | 1,40E-25 | | | |
| | TUD-PRS | 0,14 ± 0,07 | 4,74E-02 | 0,00577297 | 1.15 (1.002-1.32) | 5,92E-02 | TUD-PRS | 0,01 ± 0,04 | 8,34E-01 | 2,01522E-05 | 1.01 (0.93-1.09) | 8,34E-01 |
| | Sex | 0,98 ± 0,14 | 4,99E-12 | | | | Sex | -0,47 ± 0,08 | 1,16E-08 | | | |
| | Center | -0,09 ± 0,01 | 1,24E-09 | | | | Center | -0,12 ± 0,02 | 2,00E-09 | | | |
| | PC1 | -1,58 ± 2,41 | 5,13E-01 | | | | PC1 | -36,57 ± 3,2 | 3,42E-30 | | | |
| | PC2 | 0,92 ± 2,08 | 6,59E-01 | | | | PC2 | 31,33 ± 2,8 | 5,18E-29 | | | |
| TUD | PC3 | -0,5 ± 2,1 | 8,12E-01 | | | | PC3 | 7,32 ± 2,38 | 2,11E-03 | | | |
| | PC4 | 2,49 ± 4,46 | 5,76E-01 | | | | PC4 | -4,78 ± 3,23 | 1,40E-01 | | | |
| | PC5 | -5,13 ± 2,99 | 8,65E-02 | | | | PC5 | 0,44 ± 2,39 | 8,52E-01 | | | |
| | PC6 | -8,29 ± 4,71 | 7,81E-02 | | | | PC6 | 1,68 ± 2,38 | 4,80E-01 | | | |
| | PC7 | -1,9 ± 2,82 | 5,01E-01 | | | | PC7 | 6,73 ± 2,36 | 4,31E-03 | | | |
| | PC8 | 4,85 ± 5,04 | 3,36E-01 | | | | PC8 | 4,35 ± 2,31 | 5,91E-02 | | | |
| | PC9 | 3,77 ± 4,23 | 3,73E-01 | | | | PC9 | 6,52 ± 2,34 | 5,27E-03 | | | |
| | PC10 | -0,83 ± 2,32 | 7,19E-01 | | | | PC10 | -6,33 ± 2,29 | 5,82E-03 | | | |

* The Nagelkerke R2 correspond to the variance explained by the AF-, CanUD-, OUD-, PAU-, and TUD-PRS.

** The FDR values correspond to the corrected PRS p-value after accounting for multiple testing.

SCZ: Schizophrenia; AF: Addiction Factor; CanUD: Cannabis Use Disorder; OR: Odds ratio; OUD: Opioid Use Disorder; PAU: Problematic Alcohol Use; PRS: Polygenic Risk Score; TUD: Tobacco Use Disorder

**Table S6. Association of schizophrenia-PRSs with SUD diagnosis.**

| Trait-informed PRS | Galician cohort | | | | |
|---|---|---|---|---|---|
| | **Model variables** | **Beta ± SE** | **P-value** | **Nagelkerke R2*** | **OR (95% IC)** |
| SCZ | Intercept | 2,18 ± 0,14 | 1,90E-55 | | |
| | SCZ-PRS | 0,18 ± 0,05 | 1,42E-04 | 0,009 | 1.19 (1.09-1.30) |
| | Sex | -1,55 ± 0,1 | 2,74E-52 | | |
| | PC1 | 0,02 ± 0,15 | 9,15E-01 | | |
| | PC2 | -0,19 ± 0,18 | 2,84E-01 | | |
| | PC3 | -0,26 ± 0,18 | 1,60E-01 | | |
| | PC4 | 0,94 ± 0,46 | 4,31E-02 | | |
| | PC5 | 0,04 ± 0,12 | 7,27E-01 | | |
| | PC6 | 0 ± 0,12 | 9,85E-01 | | |
| | PC7 | 0,06 ± 0,3 | 8,39E-01 | | |
| | PC8 | 0,1 ± 0,14 | 4,67E-01 | | |
| | PC9 | -0,09 ± 0,14 | 4,93E-01 | | |
| | PC10 | -0,25 ± 0,17 | 1,43E-01 | | |

* The Nagelkerke R2 correspond to the variance explained by the SCZ-PRS.

SCZ: Schizophrenia; SUD: Substance use disorder; OR: Odds ratio; PRS: Polygenic Risk Score.

**Table S7. Causal inference analyses between schizophrenia and substance use disorders.**

| Method | $N_{SNPs}$ | Beta | SE | 95% CI | P-value* | $N_{SNPs}$ | Beta | SE | 95% CI | P-value* |
|---|---|---|---|---|---|---|---|---|---|---|
| | **SCZ → CanUD** | | | | | **CanUD → SCZ** | | | | |
| Inverse variance weighted | 130 | 0,182562986 | 0,022992636 | (0.137 - 0.227) | 2,02078E-15 | 19 | 0,406414538 | 0,080684464 | (0.248 - 0.564) | 4,72674E-07 |
| Weighted median | 130 | 0,156224609 | 0,023636857 | (0.109 - 0.202) | 3,85973E-11 | 19 | 0,355445424 | 0,059665296 | (0.238 - 0.472) | 2,56404E-09 |
| Weighted mode | 130 | 0,150002943 | 0,064004306 | (0.024 - 0.275) | 0,020625203 | 19 | 0,372596885 | 0,103002107 | (0.170 - 0.574) | 0,001969589 |
| MR-PRESSO | 126 | 0,178688091 | 0,020078755 | (0.139 - 0.218) | 5,61691E-19 | 15 | 0,340857701 | 0,054741159 | (0.233 - 0.448) | 4,7631E-10 |
| Steiger | 130 | 0,182562986 | 0,022992636 | (0.137 - 0.227) | 2,02078E-15 | 16 | 0,301356875 | 0,056609074 | (0.190 - 0.412) | 1,01805E-07 |
| | **SCZ → OUD** | | | | | **OUD → SCZ** | | | | |
| Inverse variance weighted | 39 | 0,222754859 | 0,053261673 | (0.118 - 0.327) | 2,89E-05 | 3 | 0,793896709 | 0,344326724 | (0.119 - 1.468) | 2,11E-02 |
| Weighted median | 39 | 0,135189226 | 0,054248449 | (0.028 - 0.241) | 1,27E-02 | 3 | 0,596270046 | 0,135307609 | (0.331 - 0.861) | 1,05E-05 |
| Weighted mode | 39 | 0,109093098 | 0,095073438 | (-0.077 - 0.295) | 2,58E-01 | 3 | -0,03051353 | 0,21113575 | (-0.444 - 0.383) | 8,98E-01 |
| MR-PRESSO | NA | NA | NA | NA | NA | NA | NA | NA | NA | NA |
| Steiger | NA | NA | NA | NA | NA | NA | NA | NA | NA | NA |
| | **SCZ → PAU** | | | | | **PAU → SCZ** | | | | |
| Inverse variance weighted | 130 | 0,04699676 | 0,007517652 | (0.032 - 0.061) | 4,06473E-10 | 65 | 0,677014725 | 0,199231673 | (0.286 - 1.067) | 0,000678487 |
| Weighted median | 130 | 0,041909 | 0,005730417 | (0.030 - 0.053) | 2,60409E-13 | 65 | 0,031982108 | 0,113909944 | (-0.191 - 0.255) | 0,778889422 |
| Weighted mode | 130 | 0,036505984 | 0,018960722 | (0.000 - 0.073) | 0,056386729 | NA | NA | NA | NA | NA |
| MR-PRESSO | NA | NA | NA | NA | NA | NA | NA | NA | NA | NA |
| Steiger | NA | NA | NA | NA | NA | NA | NA | NA | NA | NA |
| | **SCZ → TUD** | | | | | **TUD → SCZ** | | | | |
| Inverse variance weighted | 129 | 0,031670395 | 0,008494067 | (0.015 - 0.048) | 0,000192599 | 43 | 0,243805323 | 0,203657809 | (-0.155 - 0.642) | 0,231255026 |
| Weighted median | 129 | 0,014658579 | 0,008503143 | (-0.002 - 0.031) | 0,084725685 | NA | NA | NA | NA | NA |
| Weighted mode | NA | NA | NA | NA | NA | NA | NA | NA | NA | NA |
| MR-PRESSO | NA | NA | NA | NA | NA | NA | NA | NA | NA | NA |
| Steiger | NA | NA | NA | NA | NA | NA | NA | NA | NA | NA |

SCZ: Schizophrenia; CanUD: Cannabis Use Disorder; OUD: Opioid Use Disorder; PAU: Problematic Alcohol Use; TUD: Tobacco Use Disorder
*Adjusted p-value in the case of the inverse variance weighted method.

**Table S8. LD-independent SNPs targetting horizontal pleiotropic loci between schizophrenia and addiction factor.**

| SNP | CHR | BP | r | angle | z.scz | z.af | pval.scz | pval.af | r.pval | r.qval | theta.pval | theta.qval |
|---|---|---|---|---|---|---|---|---|---|---|---|---|
| rs4702* | 15 | 91426560 | 9,785 | 3,141 | 6,920 | 6,919 | 2,794E-21 | 1,92E-06 | 0,000 | 0,000 | 0,000 | 0,000 |
| rs1813006* | 4 | 103001649 | 11,486 | -1,949 | -5,377 | 10,149 | 2,451E-13 | 3,178E-12 | 0,000 | 0,000 | 0,000 | 0,005 |
| rs7121986* | 11 | 113355444 | 11,153 | -2,050 | -5,468 | 9,720 | 8,478E-14 | 2,478E-11 | 0,000 | 0,000 | 0,000 | 0,005 |
| rs8180995* | 8 | 143326237 | 8,437 | 3,116 | 6,004 | 5,928 | 1,767E-16 | 4,515E-05 | 0,000 | 0,000 | 0,000 | 0,005 |
| rs11210201 | 1 | 73766431 | 9,897 | -2,736 | -6,254 | 7,670 | 1,545E-17 | 1,401E-07 | 0,000 | 0,000 | 0,000 | 0,005 |
| rs114245623 | 19 | 54488872 | 4,462 | -3,142 | -3,155 | 3,155 | 1,742E-05 | 0,0303611 | 0,000 | 0,003 | 0,000 | 0,005 |
| rs35797074 | 3 | 161783701 | 4,545 | -3,142 | 3,214 | 3,214 | 1,062E-05 | 0,0269786 | 0,000 | 0,002 | 0,000 | 0,005 |
| rs71367544 | 18 | 77574374 | 7,339 | -3,141 | -5,189 | 5,190 | 1,428E-12 | 0,0003688 | 0,000 | 0,000 | 0,000 | 0,005 |
| rs7580750 | 2 | 73788126 | 5,703 | -3,141 | -4,033 | 4,033 | 3,515E-08 | 0,0056475 | 0,000 | 0,000 | 0,000 | 0,005 |
| rs7829812 | 8 | 89722274 | 4,801 | -3,142 | -3,395 | 3,395 | 3,463E-06 | 0,0198114 | 0,000 | 0,001 | 0,000 | 0,005 |
| rs2287184 | 12 | 109970251 | 3,911 | 3,141 | 2,765 | 2,765 | 0,0001622 | 0,0570359 | 0,000 | 0,013 | 0,000 | 0,007 |
| rs4821990 | 22 | 41438710 | 6,405 | -3,139 | -4,527 | 4,531 | 6,561E-10 | 0,0018729 | 0,000 | 0,000 | 0,000 | 0,008 |
| rs61882758 | 11 | 46611805 | 7,737 | -3,121 | -5,443 | 5,498 | 1,062E-13 | 0,0001612 | 0,000 | 0,000 | 0,000 | 0,008 |
| rs9842022 | 3 | 669852 | 3,368 | 3,142 | -2,382 | 2,382 | 0,001131 | 0,1021534 | 0,003 | 0,049 | 0,000 | 0,008 |
| rs11682175* | 2 | 57987593 | 9,539 | -2,679 | -5,923 | 7,478 | 8,699E-16 | 2,847E-07 | 0,000 | 0,000 | 0,000 | 0,009 |
| rs7730945 | 5 | 153679702 | 4,728 | 3,141 | -3,343 | 3,343 | 4,693E-06 | 0,0217946 | 0,000 | 0,001 | 0,000 | 0,009 |
| rs9565485* | 13 | 79881563 | 5,912 | 3,140 | -4,182 | 4,179 | 1,089E-08 | 0,0041326 | 0,000 | 0,000 | 0,000 | 0,010 |
| rs10220824 | 15 | 89839487 | 4,283 | 3,141 | 3,029 | 3,028 | 3,051E-05 | 0,037148 | 0,000 | 0,004 | 0,000 | 0,013 |
| rs2494633 | 1 | 2398763 | 4,739 | 3,141 | -3,351 | 3,351 | 4,852E-06 | 0,0214876 | 0,000 | 0,001 | 0,000 | 0,013 |
| rs12892189* | 14 | 104319989 | 9,683 | -2,332 | -5,331 | 8,083 | 3,477E-13 | 2,866E-08 | 0,000 | 0,000 | 0,000 | 0,013 |
| rs1108074 | 12 | 2333484 | 8,622 | 2,986 | -6,329 | 5,854 | 7,285E-18 | 5,898E-05 | 0,000 | 0,000 | 0,000 | 0,014 |
| rs17160838 | 7 | 138952700 | 4,040 | 3,141 | 2,857 | 2,856 | 9,091E-05 | 0,0493382 | 0,000 | 0,009 | 0,000 | 0,014 |
| rs55814644 | 7 | 110011981 | 4,415 | 3,141 | 3,122 | 3,122 | 0,0000176 | 0,0316853 | 0,000 | 0,003 | 0,000 | 0,014 |
| rs7560829 | 2 | 241366623 | 3,779 | -3,141 | -2,672 | 2,673 | 0,000245 | 0,0666561 | 0,001 | 0,019 | 0,000 | 0,014 |
| rs12022771 | 1 | 97870315 | 6,398 | -3,137 | -4,519 | 4,529 | 5,727E-10 | 0,0018824 | 0,000 | 0,000 | 0,000 | 0,014 |
| rs159961 | 1 | 8484228 | 5,550 | 3,140 | -3,925 | 3,923 | 8,844E-08 | 0,0071022 | 0,000 | 0,000 | 0,000 | 0,014 |
| rs74950305 | 19 | 19639448 | 5,269 | 3,140 | -3,727 | 3,725 | 3,344E-07 | 0,010586 | 0,000 | 0,000 | 0,000 | 0,014 |
| rs10469137 | 18 | 59621444 | 4,200 | -3,141 | 2,970 | 2,970 | 0,0000444 | 0,0409363 | 0,000 | 0,006 | 0,000 | 0,015 |
| rs6428028 | 1 | 190666694 | 3,902 | 3,141 | -2,759 | 2,759 | 0,0001699 | 0,0583381 | 0,000 | 0,014 | 0,000 | 0,015 |
| rs7291996 | 22 | 42492104 | 6,780 | 3,129 | 4,808 | 4,779 | 5,524E-11 | 0,0010048 | 0,000 | 0,000 | 0,000 | 0,015 |
| rs28529862 | 14 | 60181498 | 4,028 | -3,141 | -2,848 | 2,849 | 0,0001035 | 0,050598 | 0,000 | 0,010 | 0,000 | 0,016 |
| rs159542 | 5 | 60485778 | 7,087 | -3,128 | 4,994 | 5,029 | 7,947E-12 | 0,0005388 | 0,000 | 0,000 | 0,000 | 0,016 |
| rs4664191 | 2 | 156852749 | 5,656 | -3,140 | 3,998 | 4,001 | 4,905E-08 | 0,0058932 | 0,000 | 0,000 | 0,000 | 0,016 |

| | | | | | | | | | | | | |
|---|---|---|---|---|---|---|---|---|---|---|---|---|
| rs2721781 | 7 | 24725834 | 4,984 | 3,141 | -3,525 | 3,523 | 1,373E-06 | 0,0156221 | 0,000 | 0,000 | 0,000 | 0,017 |
| rs1913499 | 5 | 101983210 | 4,112 | -3,141 | -2,907 | 2,908 | 6,598E-05 | 0,0459806 | 0,000 | 0,008 | 0,000 | 0,017 |
| rs4619651 | 2 | 97416153 | 4,033 | -3,141 | 2,852 | 2,852 | 9,348E-05 | 0,0496653 | 0,000 | 0,009 | 0,000 | 0,017 |
| rs112016302 | 2 | 58243567 | 5,461 | 3,140 | -3,864 | 3,860 | 1,282E-07 | 0,0080815 | 0,000 | 0,000 | 0,000 | 0,017 |
| rs12141694 | 1 | 166572901 | 4,152 | 3,141 | -2,936 | 2,936 | 6,109E-05 | 0,0439572 | 0,000 | 0,007 | 0,000 | 0,017 |
| rs7565005 | 2 | 201013277 | 5,731 | 3,139 | -4,054 | 4,050 | 3,105E-08 | 0,0054495 | 0,000 | 0,000 | 0,000 | 0,018 |
| rs11791983* | 9 | 122182701 | 3,530 | 3,141 | -2,497 | 2,496 | 0,0006863 | 0,0867196 | 0,002 | 0,034 | 0,000 | 0,019 |
| rs7933981* | 11 | 113438068 | 11,135 | -1,731 | 4,669 | 10,108 | 1,896E-10 | 3,617E-12 | 0,000 | 0,000 | 0,000 | 0,019 |
| rs1835719 | 2 | 201146000 | 3,457 | 3,141 | 2,444 | 2,444 | 0,0008221 | 0,0925774 | 0,003 | 0,040 | 0,000 | 0,019 |
| rs331934 | 5 | 98139720 | 3,468 | 3,141 | -2,452 | 2,452 | 0,0008294 | 0,0924797 | 0,002 | 0,039 | 0,000 | 0,019 |
| rs422149 | 13 | 114931086 | 4,972 | 3,140 | 3,517 | 3,514 | 1,483E-06 | 0,015583 | 0,000 | 0,000 | 0,000 | 0,019 |
| rs425405 | 6 | 32887469 | 3,652 | 3,141 | -2,582 | 2,582 | 0,0004065 | 0,0764245 | 0,001 | 0,026 | 0,000 | 0,019 |
| rs442439 | 6 | 28573006 | 8,450 | 2,899 | 6,326 | 5,601 | 6,451E-18 | 0,0001154 | 0,000 | 0,000 | 0,000 | 0,019 |
| rs4665177 | 2 | 22537264 | 4,169 | -3,141 | -2,948 | 2,949 | 5,952E-05 | 0,0430247 | 0,000 | 0,006 | 0,000 | 0,019 |
| rs6730085 | 2 | 172864845 | 3,591 | 3,141 | 2,540 | 2,539 | 0,0005272 | 0,0805614 | 0,002 | 0,030 | 0,000 | 0,019 |
| rs4300612* | 15 | 83919472 | 3,625 | -3,141 | 2,563 | 2,564 | 0,000451 | 0,0776709 | 0,001 | 0,027 | 0,000 | 0,019 |
| rs12431331 | 13 | 50142595 | 3,656 | -3,141 | -2,585 | 2,586 | 0,0004265 | 0,0760182 | 0,001 | 0,025 | 0,000 | 0,019 |
| rs9818755* | 3 | 107898593 | 4,282 | 3,141 | 3,029 | 3,027 | 3,345E-05 | 0,0372231 | 0,000 | 0,005 | 0,000 | 0,020 |
| rs9939774 | 16 | 30068354 | 6,760 | -3,112 | 4,744 | 4,816 | 8,134E-11 | 0,0009197 | 0,000 | 0,000 | 0,000 | 0,020 |
| rs4576401 | 8 | 102811265 | 3,423 | 3,141 | -2,421 | 2,420 | 0,0009685 | 0,0967564 | 0,003 | 0,043 | 0,000 | 0,021 |
| rs12898090 | 14 | 89406377 | 4,432 | 3,140 | 3,135 | 3,133 | 0,0000161 | 0,0310781 | 0,000 | 0,003 | 0,000 | 0,021 |
| rs57364768* | 16 | 89743000 | 4,437 | -3,140 | 3,137 | 3,139 | 1,733E-05 | 0,030761 | 0,000 | 0,003 | 0,000 | 0,022 |
| rs11678993 | 2 | 213512215 | 3,722 | -3,141 | -2,631 | 2,632 | 0,0003131 | 0,070873 | 0,001 | 0,022 | 0,000 | 0,023 |
| rs2824021 | 21 | 18119490 | 3,806 | 3,141 | 2,692 | 2,690 | 0,0002476 | 0,06408 | 0,001 | 0,017 | 0,000 | 0,023 |
| rs13393198 | 2 | 155965460 | 3,488 | 3,141 | 2,467 | 2,466 | 0,000698 | 0,0897064 | 0,002 | 0,038 | 0,000 | 0,024 |
| rs4076439 | 8 | 139621368 | 4,000 | 3,141 | 2,829 | 2,827 | 0,0001075 | 0,0516814 | 0,000 | 0,010 | 0,000 | 0,024 |
| rs1987744 | 18 | 5412426 | 3,581 | 3,141 | -2,532 | 2,532 | 0,0005477 | 0,0823497 | 0,002 | 0,030 | 0,000 | 0,025 |
| rs11792283 | 9 | 35590106 | 3,466 | -3,141 | 2,451 | 2,451 | 0,000762 | 0,0915963 | 0,002 | 0,040 | 0,000 | 0,025 |
| rs2848499 | 11 | 98118537 | 3,795 | 3,141 | -2,684 | 2,682 | 0,0002486 | 0,0656474 | 0,001 | 0,018 | 0,000 | 0,025 |
| rs6759693 | 2 | 5399202 | 3,379 | -3,141 | -2,389 | 2,390 | 0,001171 | 0,1010033 | 0,003 | 0,048 | 0,000 | 0,025 |
| rs4384732 | 2 | 24033707 | 4,001 | -3,140 | 2,828 | 2,830 | 0,000113 | 0,0514448 | 0,000 | 0,010 | 0,000 | 0,025 |
| rs4332635* | 13 | 41901855 | 4,819 | 3,139 | -3,410 | 3,405 | 3,386E-06 | 0,0194503 | 0,000 | 0,001 | 0,000 | 0,026 |
| rs35560064 | 20 | 48016283 | 3,611 | 3,141 | -2,554 | 2,553 | 0,0004865 | 0,0797669 | 0,001 | 0,028 | 0,000 | 0,026 |
| rs57901004 | 17 | 47011897 | 3,528 | 3,141 | -2,495 | 2,494 | 0,0006462 | 0,0869355 | 0,002 | 0,034 | 0,000 | 0,026 |
| rs13034670 | 2 | 146122551 | 3,398 | -3,141 | -2,403 | 2,403 | 0,001094 | 0,0990749 | 0,003 | 0,046 | 0,000 | 0,027 |

| | | | | | | | | | | | | |
|---|---|---|---|---|---|---|---|---|---|---|---|---|
| rs2238281 | 14 | 72424912 | 6,646 | 3,116 | -4,730 | 4,669 | 1,067E-10 | 0,0013546 | 0,000 | 0,000 | 0,000 | 0,027 |
| rs8138039 | 22 | 41442653 | 3,883 | -3,140 | 2,745 | 2,746 | 0,0001632 | 0,058737 | 0,001 | 0,014 | 0,000 | 0,027 |
| rs10438359 | 15 | 70172513 | 3,716 | 3,141 | 2,628 | 2,627 | 0,0003108 | 0,0706346 | 0,001 | 0,022 | 0,000 | 0,027 |
| rs9379832 | 6 | 26186200 | 6,034 | -3,128 | -4,252 | 4,282 | 7,019E-09 | 0,0033007 | 0,000 | 0,000 | 0,000 | 0,028 |
| rs2726759 | 4 | 150511435 | 3,523 | -3,141 | -2,490 | 2,492 | 0,0006863 | 0,0872975 | 0,002 | 0,035 | 0,000 | 0,028 |
| rs7413681 | 1 | 97177371 | 4,799 | 3,138 | -3,396 | 3,391 | 3,335E-06 | 0,0199735 | 0,000 | 0,001 | 0,000 | 0,028 |
| rs4671378 | 2 | 60494312 | 4,496 | -3,139 | 3,177 | 3,181 | 1,449E-05 | 0,0285847 | 0,000 | 0,002 | 0,000 | 0,029 |
| rs79081799* | 1 | 91433264 | 4,187 | -3,140 | -2,959 | 2,962 | 5,281E-05 | 0,0420928 | 0,000 | 0,006 | 0,000 | 0,029 |
| rs34493921 | 1 | 175122759 | 3,801 | -3,140 | -2,686 | 2,688 | 0,0002377 | 0,0650662 | 0,001 | 0,018 | 0,000 | 0,030 |
| rs62442944 | 7 | 2015047 | 5,469 | -3,132 | -3,858 | 3,876 | 1,591E-07 | 0,0078155 | 0,000 | 0,000 | 0,000 | 0,030 |
| rs7198943 | 16 | 9970419 | 4,441 | -3,139 | -3,138 | 3,143 | 1,731E-05 | 0,0310335 | 0,000 | 0,003 | 0,000 | 0,030 |
| rs75365900 | 5 | 152810929 | 4,201 | 3,139 | 2,972 | 2,969 | 4,709E-05 | 0,0410427 | 0,000 | 0,006 | 0,000 | 0,030 |
| rs7602823 | 2 | 27309406 | 4,488 | -3,139 | 3,171 | 3,176 | 1,606E-05 | 0,0288465 | 0,000 | 0,002 | 0,000 | 0,030 |
| rs3130099* | 6 | 33282181 | 4,598 | 3,138 | -3,254 | 3,249 | 9,11E-06 | 0,0257895 | 0,000 | 0,002 | 0,000 | 0,031 |
| rs113755698 | 4 | 85501322 | 3,731 | 3,140 | -2,639 | 2,637 | 0,00032 | 0,0703119 | 0,001 | 0,021 | 0,000 | 0,031 |
| rs884763 | 18 | 5052648 | 4,850 | -3,137 | 3,425 | 3,433 | 2,778E-06 | 0,0181389 | 0,000 | 0,001 | 0,000 | 0,031 |
| rs2861344* | 18 | 53446374 | 7,453 | -2,974 | 5,045 | 5,486 | 5,864E-12 | 0,00016 | 0,000 | 0,000 | 0,000 | 0,033 |
| rs10456874 | 6 | 111608300 | 4,887 | -3,137 | -3,451 | 3,460 | 2,446E-06 | 0,0175794 | 0,000 | 0,001 | 0,000 | 0,033 |
| rs9932414 | 16 | 68219969 | 4,090 | -3,139 | -2,890 | 2,893 | 7,853E-05 | 0,0470843 | 0,000 | 0,008 | 0,000 | 0,033 |
| rs8093631 | 18 | 42134697 | 3,532 | 3,140 | -2,499 | 2,497 | 0,0006491 | 0,0866193 | 0,002 | 0,034 | 0,000 | 0,034 |
| rs111321345 | 6 | 111013145 | 3,493 | -3,140 | -2,469 | 2,471 | 0,0007636 | 0,0899584 | 0,002 | 0,037 | 0,000 | 0,034 |
| rs12621041 | 2 | 144725082 | 3,923 | 3,139 | -2,775 | 2,772 | 0,0001544 | 0,0571356 | 0,000 | 0,013 | 0,000 | 0,034 |
| rs1264306 | 6 | 30882203 | 4,141 | 3,139 | -2,930 | 2,926 | 6,056E-05 | 0,0446236 | 0,000 | 0,007 | 0,000 | 0,034 |
| rs2163729 | 3 | 71266371 | 4,748 | 3,137 | 3,362 | 3,353 | 3,767E-06 | 0,0210086 | 0,000 | 0,001 | 0,000 | 0,034 |
| rs4781521 | 16 | 13741889 | 4,773 | -3,136 | -3,370 | 3,380 | 3,85E-06 | 0,0203671 | 0,000 | 0,001 | 0,000 | 0,034 |
| rs4810237 | 20 | 37487150 | 5,147 | 3,134 | -3,647 | 3,633 | 6,073E-07 | 0,0126707 | 0,000 | 0,000 | 0,000 | 0,034 |
| rs60504229 | 3 | 35084608 | 3,691 | 3,140 | -2,611 | 2,609 | 0,0003514 | 0,0734351 | 0,001 | 0,023 | 0,000 | 0,034 |
| rs6482355 | 10 | 18651860 | 5,249 | -3,135 | -3,706 | 3,718 | 4,47E-07 | 0,0107259 | 0,000 | 0,000 | 0,000 | 0,034 |
| rs6536 | 6 | 131160876 | 3,711 | -3,140 | -2,623 | 2,626 | 0,0003583 | 0,0715845 | 0,001 | 0,022 | 0,000 | 0,034 |
| rs6757752 | 2 | 225272366 | 3,994 | 3,139 | -2,826 | 2,823 | 0,0001157 | 0,0527476 | 0,000 | 0,010 | 0,000 | 0,034 |
| rs7144423 | 14 | 35572547 | 4,735 | -3,137 | 3,344 | 3,352 | 4,475E-06 | 0,0210437 | 0,000 | 0,001 | 0,000 | 0,034 |
| rs994082 | 8 | 115557887 | 3,918 | -3,139 | -2,769 | 2,772 | 0,0001467 | 0,0571306 | 0,000 | 0,013 | 0,000 | 0,034 |
| rs55727458 | 13 | 102031359 | 4,957 | -3,134 | -3,498 | 3,512 | 1,769E-06 | 0,0159559 | 0,000 | 0,000 | 0,000 | 0,034 |
| rs59017643 | 2 | 196411559 | 4,321 | -3,138 | -3,052 | 3,058 | 3,031E-05 | 0,0358526 | 0,000 | 0,004 | 0,000 | 0,034 |
| rs2534671 | 6 | 31465661 | 3,404 | 3,140 | -2,408 | 2,406 | 0,0009767 | 0,0987276 | 0,003 | 0,045 | 0,000 | 0,035 |

| | | | | | | | | | | | | |
|---|---|---|---|---|---|---|---|---|---|---|---|---|
| rs6493261 | 15 | 47427443 | 3,682 | -3,139 | -2,602 | 2,605 | 0,0003897 | 0,0738364 | 0,001 | 0,024 | 0,000 | 0,035 |
| rs10899438 | 11 | 77803997 | 3,516 | 3,140 | -2,488 | 2,485 | 0,0006955 | 0,0880977 | 0,002 | 0,035 | 0,000 | 0,035 |
| rs1644007 | 12 | 44689702 | 3,431 | -3,140 | -2,425 | 2,427 | 0,0009155 | 0,0957518 | 0,003 | 0,043 | 0,000 | 0,035 |
| rs2172733 | 3 | 107470258 | 3,535 | 3,140 | -2,501 | 2,499 | 0,0006715 | 0,08639 | 0,002 | 0,034 | 0,000 | 0,035 |
| rs12814239* | 12 | 57569478 | 7,007 | 3,024 | 5,098 | 4,807 | 2,872E-12 | 0,0009375 | 0,000 | 0,000 | 0,000 | 0,035 |
| rs698905 | 2 | 206253276 | 3,612 | 3,140 | -2,555 | 2,553 | 0,0004656 | 0,0798019 | 0,001 | 0,028 | 0,000 | 0,035 |
| rs12576181* | 11 | 113367370 | 3,906 | 3,139 | -2,764 | 2,760 | 0,000159 | 0,0582494 | 0,000 | 0,013 | 0,000 | 0,036 |
| rs143319035 | 7 | 131598900 | 3,923 | 3,139 | 2,776 | 2,772 | 0,0001472 | 0,0564479 | 0,000 | 0,013 | 0,000 | 0,036 |
| rs17765013 | 6 | 90641851 | 3,690 | 3,139 | 2,610 | 2,607 | 0,0003494 | 0,0727485 | 0,001 | 0,023 | 0,000 | 0,036 |
| rs1865024 | 10 | 78720204 | 4,816 | -3,134 | 3,399 | 3,412 | 3,582E-06 | 0,0188677 | 0,000 | 0,001 | 0,000 | 0,036 |
| rs1906145 | 10 | 3737199 | 4,316 | -3,137 | -3,049 | 3,055 | 2,932E-05 | 0,0360355 | 0,000 | 0,004 | 0,000 | 0,036 |
| rs907482 | 12 | 122747023 | 3,713 | -3,139 | -2,624 | 2,627 | 0,0003314 | 0,0714028 | 0,001 | 0,022 | 0,000 | 0,036 |
| rs13042129* | 20 | 38301402 | 3,411 | 3,140 | -2,413 | 2,411 | 0,0009276 | 0,0980334 | 0,003 | 0,045 | 0,000 | 0,036 |
| rs75907840 | 3 | 181420180 | 4,727 | -3,134 | 3,337 | 3,349 | 4,88E-06 | 0,0211851 | 0,000 | 0,001 | 0,000 | 0,036 |
| rs549381 | 18 | 26424631 | 3,507 | -3,140 | -2,479 | 2,481 | 0,000759 | 0,0886038 | 0,002 | 0,036 | 0,000 | 0,037 |
| rs17773723 | 5 | 57794834 | 4,390 | -3,137 | 3,100 | 3,108 | 2,061E-05 | 0,032441 | 0,000 | 0,003 | 0,000 | 0,037 |
| rs17862085* | 4 | 118657117 | 4,953 | 3,132 | -3,510 | 3,494 | 1,544E-06 | 0,016495 | 0,000 | 0,000 | 0,000 | 0,037 |
| rs2492971 | 1 | 72887188 | 4,036 | 3,138 | 2,857 | 2,851 | 9,352E-05 | 0,0497167 | 0,000 | 0,009 | 0,000 | 0,037 |
| rs79777688 | 3 | 69417442 | 4,093 | -3,138 | 2,892 | 2,897 | 8,104E-05 | 0,0461634 | 0,000 | 0,008 | 0,000 | 0,037 |
| rs11706295 | 3 | 24258084 | 3,658 | -3,139 | -2,585 | 2,588 | 0,0004147 | 0,075723 | 0,001 | 0,025 | 0,000 | 0,038 |
| rs2842188 | 1 | 44014280 | 4,197 | 3,137 | -2,971 | 2,964 | 4,853E-05 | 0,0419336 | 0,000 | 0,006 | 0,000 | 0,038 |
| rs62297928 | 4 | 19171165 | 3,687 | -3,139 | 2,606 | 2,609 | 0,0003521 | 0,0725526 | 0,001 | 0,023 | 0,000 | 0,038 |
| rs77025955 | 11 | 134489259 | 3,570 | -3,139 | -2,523 | 2,526 | 0,0005615 | 0,0830386 | 0,002 | 0,031 | 0,000 | 0,038 |
| rs7832419 | 8 | 61176417 | 4,092 | 3,137 | 2,897 | 2,891 | 7,778E-05 | 0,0466574 | 0,000 | 0,008 | 0,000 | 0,038 |
| rs12658941 | 5 | 152005102 | 4,428 | -3,136 | -3,126 | 3,136 | 1,942E-05 | 0,0314055 | 0,000 | 0,003 | 0,000 | 0,039 |
| rs1796518 | 6 | 26388672 | 6,926 | -2,993 | 4,712 | 5,076 | 9,228E-11 | 0,0004771 | 0,000 | 0,000 | 0,000 | 0,039 |
| rs4377607* | 4 | 115570238 | 3,420 | 3,139 | -2,420 | 2,417 | 0,0009477 | 0,0971451 | 0,003 | 0,044 | 0,000 | 0,039 |
| rs75688627 | 21 | 38736954 | 3,802 | -3,138 | -2,686 | 2,690 | 0,0002519 | 0,0648699 | 0,001 | 0,018 | 0,000 | 0,039 |
| rs11814460 | 10 | 103261513 | 3,923 | 3,138 | 2,776 | 2,771 | 0,0001469 | 0,0564967 | 0,000 | 0,013 | 0,000 | 0,039 |
| rs4287009* | 9 | 122574967 | 4,009 | 3,138 | -2,838 | 2,832 | 9,827E-05 | 0,0519727 | 0,000 | 0,010 | 0,000 | 0,040 |
| rs142367172 | 11 | 29925504 | 3,552 | 3,139 | -2,513 | 2,510 | 0,0005993 | 0,0850163 | 0,002 | 0,032 | 0,000 | 0,040 |
| rs122730 | 5 | 127220271 | 3,697 | -3,139 | 2,613 | 2,616 | 0,0003399 | 0,0717537 | 0,001 | 0,023 | 0,000 | 0,040 |
| rs2385537 | 7 | 104755514 | 4,743 | -3,133 | -3,346 | 3,361 | 0,0000051 | 0,0210762 | 0,000 | 0,001 | 0,000 | 0,040 |
| rs4709845 | 6 | 164900598 | 4,497 | 3,135 | 3,185 | 3,174 | 1,239E-05 | 0,0289143 | 0,000 | 0,002 | 0,000 | 0,041 |
| rs12068631 | 1 | 151005763 | 3,592 | 3,139 | 2,541 | 2,538 | 0,0004996 | 0,080711 | 0,002 | 0,029 | 0,000 | 0,041 |

| | | | | | | | | | | | | |
|---|---|---|---|---|---|---|---|---|---|---|---|---|
| rs35399732 | 8 | 56589524 | 3,389 | 3,139 | -2,398 | 2,395 | 0,001006 | 0,1002941 | 0,003 | 0,047 | 0,000 | 0,041 |
| rs1859960 | 19 | 32659730 | 4,237 | -3,136 | 2,992 | 3,000 | 4,184E-05 | 0,0389618 | 0,000 | 0,005 | 0,000 | 0,041 |
| rs2901616 | 1 | 72122959 | 5,080 | 3,129 | -3,603 | 3,581 | 8,786E-07 | 0,0139994 | 0,000 | 0,000 | 0,000 | 0,042 |
| rs10240767 | 7 | 44632244 | 3,530 | -3,139 | 2,494 | 2,498 | 0,0006318 | 0,085633 | 0,002 | 0,034 | 0,000 | 0,042 |
| rs74800502 | 20 | 42068341 | 4,145 | 3,136 | 2,935 | 2,927 | 5,849E-05 | 0,0439895 | 0,000 | 0,007 | 0,000 | 0,043 |
| rs10071595 | 5 | 107622552 | 3,431 | 3,139 | -2,428 | 2,425 | 0,0009441 | 0,0961348 | 0,003 | 0,043 | 0,000 | 0,043 |
| rs10839224 | 11 | 49146066 | 3,486 | -3,139 | -2,464 | 2,467 | 0,0007454 | 0,0904579 | 0,002 | 0,038 | 0,000 | 0,043 |
| rs2073779 | 22 | 20077863 | 3,358 | 3,139 | 2,376 | 2,373 | 0,001147 | 0,1024101 | 0,004 | 0,050 | 0,000 | 0,043 |
| rs5996255 | 22 | 43407957 | 3,938 | 3,137 | -2,787 | 2,781 | 0,0001511 | 0,0563239 | 0,000 | 0,012 | 0,000 | 0,043 |
| rs4799358 | 18 | 31586132 | 3,462 | -3,139 | 2,446 | 2,450 | 0,0007896 | 0,0917814 | 0,002 | 0,040 | 0,000 | 0,043 |
| rs2043773 | 2 | 201330331 | 4,339 | 3,134 | 3,073 | 3,062 | 2,561E-05 | 0,0350746 | 0,000 | 0,004 | 0,000 | 0,043 |
| rs7205519* | 16 | 89819816 | 4,116 | 3,136 | 2,914 | 2,906 | 6,489E-05 | 0,0454931 | 0,000 | 0,007 | 0,000 | 0,043 |
| rs149910930 | 7 | 71609523 | 4,483 | 3,133 | -3,177 | 3,163 | 1,387E-05 | 0,0299476 | 0,000 | 0,002 | 0,000 | 0,043 |
| rs4544142 | 13 | 81799583 | 3,562 | -3,138 | -2,517 | 2,521 | 0,0005544 | 0,0836605 | 0,002 | 0,032 | 0,000 | 0,043 |
| rs2674023 | 2 | 59863721 | 3,539 | -3,138 | -2,500 | 2,504 | 0,0006636 | 0,0856913 | 0,002 | 0,033 | 0,000 | 0,044 |
| rs11918777 | 3 | 16856216 | 8,453 | -2,131 | -4,293 | 7,282 | 4,493E-09 | 5,731E-07 | 0,000 | 0,000 | 0,000 | 0,044 |
| rs7287806 | 22 | 29213011 | 4,274 | -3,135 | -3,017 | 3,028 | 3,634E-05 | 0,0377378 | 0,000 | 0,005 | 0,000 | 0,044 |
| rs9912766 | 17 | 30712780 | 3,926 | -3,136 | 2,772 | 2,779 | 0,0001571 | 0,0557813 | 0,000 | 0,013 | 0,000 | 0,044 |
| rs7646822 | 3 | 161346817 | 4,207 | 3,135 | 2,980 | 2,970 | 4,086E-05 | 0,0409768 | 0,000 | 0,006 | 0,000 | 0,045 |
| rs1211661 | 1 | 98532848 | 6,456 | 3,035 | -4,685 | 4,441 | 1,376E-10 | 0,0023068 | 0,000 | 0,000 | 0,000 | 0,045 |
| rs17101012 | 14 | 33869858 | 3,453 | 3,138 | 2,443 | 2,439 | 0,0008488 | 0,0932089 | 0,003 | 0,041 | 0,000 | 0,046 |
| rs16822842 | 2 | 144403290 | 3,373 | 3,138 | 2,387 | 2,383 | 0,001052 | 0,1009653 | 0,003 | 0,048 | 0,000 | 0,046 |
| rs6987670 | 8 | 9883177 | 4,821 | -3,128 | -3,398 | 3,421 | 3,946E-06 | 0,0189044 | 0,000 | 0,001 | 0,000 | 0,046 |
| rs6886 | 2 | 85622059 | 3,509 | 3,138 | 2,483 | 2,479 | 0,0006817 | 0,087992 | 0,002 | 0,036 | 0,000 | 0,046 |
| rs37040 | 16 | 58578262 | 3,528 | 3,138 | -2,497 | 2,492 | 0,0006489 | 0,087225 | 0,002 | 0,034 | 0,000 | 0,046 |
| rs1016501 | 6 | 163908942 | 4,586 | 3,130 | -3,252 | 3,233 | 9,792E-06 | 0,0265115 | 0,000 | 0,002 | 0,000 | 0,046 |
| rs11579023 | 1 | 35574197 | 3,401 | 3,138 | -2,407 | 2,403 | 0,001053 | 0,099127 | 0,003 | 0,046 | 0,000 | 0,046 |
| rs11681451 | 2 | 225334104 | 5,781 | -3,081 | 4,026 | 4,150 | 3,312E-08 | 0,0042954 | 0,000 | 0,000 | 0,000 | 0,046 |
| rs28625158 | 7 | 71811894 | 3,844 | 3,136 | -2,722 | 2,715 | 0,0002091 | 0,0624783 | 0,001 | 0,016 | 0,000 | 0,046 |
| rs302967 | 6 | 25126041 | 3,649 | -3,137 | 2,578 | 2,583 | 0,0003994 | 0,0754085 | 0,001 | 0,026 | 0,000 | 0,046 |
| rs6065579 | 20 | 41765490 | 4,027 | -3,135 | 2,843 | 2,853 | 9,576E-05 | 0,0496322 | 0,000 | 0,010 | 0,000 | 0,046 |
| rs62369905 | 5 | 45095000 | 4,508 | 3,131 | -3,196 | 3,180 | 1,419E-05 | 0,0291145 | 0,000 | 0,002 | 0,000 | 0,046 |
| rs7156664 | 14 | 63976756 | 4,135 | -3,134 | -2,919 | 2,929 | 6,926E-05 | 0,0444095 | 0,000 | 0,007 | 0,000 | 0,046 |
| rs7207400 | 17 | 43824360 | 4,689 | -3,129 | 3,305 | 3,326 | 5,789E-06 | 0,0220738 | 0,000 | 0,001 | 0,000 | 0,046 |
| rs9981253 | 21 | 39591296 | 3,439 | 3,138 | -2,434 | 2,429 | 0,0009299 | 0,0954728 | 0,003 | 0,042 | 0,000 | 0,046 |

| | | | | | | | | | | | | |
|---|---|---|---|---|---|---|---|---|---|---|---|---|
| rs17011040 | 2 | 124944108 | 3,791 | -3,136 | -2,677 | 2,684 | 0,0002499 | 0,0654967 | 0,001 | 0,018 | 0,000 | 0,046 |
| rs880451* | 12 | 75212368 | 3,783 | 3,136 | 2,678 | 2,671 | 0,0002554 | 0,0660169 | 0,001 | 0,019 | 0,000 | 0,046 |
| rs2300990 | 5 | 109033393 | 4,936 | -3,124 | -3,475 | 3,506 | 2,055E-06 | 0,0161291 | 0,000 | 0,000 | 0,000 | 0,047 |
| rs8014517 | 14 | 84697718 | 4,801 | 3,127 | -3,407 | 3,382 | 0,0000034 | 0,0203058 | 0,000 | 0,001 | 0,000 | 0,047 |
| rs116919748 | 12 | 110851422 | 5,196 | -3,123 | 3,657 | 3,691 | 6,018E-07 | 0,0110754 | 0,000 | 0,000 | 0,000 | 0,047 |
| rs7110722 | 11 | 466140 | 3,365 | -3,138 | 2,378 | 2,382 | 0,001133 | 0,1011933 | 0,003 | 0,049 | 0,000 | 0,047 |
| rs17190727 | 15 | 59073790 | 4,952 | -3,123 | 3,486 | 3,518 | 2,053E-06 | 0,0154856 | 0,000 | 0,000 | 0,000 | 0,047 |
| rs11925334 | 3 | 24153725 | 3,794 | -3,136 | -2,679 | 2,686 | 0,0002436 | 0,0652531 | 0,001 | 0,018 | 0,000 | 0,047 |
| rs4518538 | 6 | 123056035 | 3,810 | 3,136 | 2,698 | 2,690 | 0,0002125 | 0,0641158 | 0,001 | 0,017 | 0,000 | 0,047 |
| rs929579 | 14 | 75330697 | 4,114 | 3,134 | 2,914 | 2,903 | 7,184E-05 | 0,0456971 | 0,000 | 0,007 | 0,000 | 0,047 |
| rs9420* | 11 | 57510294 | 8,434 | -2,108 | 4,242 | 7,290 | 6,647E-09 | 5,338E-07 | 0,000 | 0,000 | 0,000 | 0,048 |
| rs72685748 | 1 | 111371369 | 3,561 | 3,137 | -2,521 | 2,516 | 0,0005883 | 0,0843011 | 0,002 | 0,032 | 0,000 | 0,048 |
| rs7942967* | 11 | 117486730 | 3,758 | 3,136 | 2,661 | 2,653 | 0,0002835 | 0,0678345 | 0,001 | 0,020 | 0,000 | 0,048 |
| rs657717 | 7 | 4180453 | 3,986 | 3,134 | -2,823 | 2,813 | 0,0001129 | 0,0535382 | 0,000 | 0,011 | 0,000 | 0,048 |
| rs11649759 | 17 | 2019048 | 4,030 | -3,134 | 2,844 | 2,855 | 0,0001012 | 0,0494426 | 0,000 | 0,010 | 0,000 | 0,049 |
| rs160272 | 5 | 93668741 | 3,498 | 3,137 | -2,477 | 2,471 | 0,0007202 | 0,0899625 | 0,002 | 0,037 | 0,000 | 0,049 |
| rs11715723 | 3 | 180276265 | 4,707 | 3,126 | 3,341 | 3,315 | 4,353E-06 | 0,0225149 | 0,000 | 0,001 | 0,000 | 0,049 |
| rs1608 | 22 | 50320943 | 3,717 | -3,136 | -2,624 | 2,632 | 0,0003262 | 0,0709072 | 0,001 | 0,022 | 0,000 | 0,049 |
| rs17194371 | 3 | 2464724 | 3,808 | -3,135 | -2,688 | 2,697 | 0,0002471 | 0,0642243 | 0,001 | 0,017 | 0,000 | 0,049 |
| rs12030332 | 1 | 97779535 | 4,539 | 3,128 | 3,220 | 3,198 | 9,758E-06 | 0,0277269 | 0,000 | 0,002 | 0,000 | 0,049 |
| rs7765920 | 6 | 26514771 | 3,771 | -3,136 | 2,662 | 2,670 | 0,0002809 | 0,0660981 | 0,001 | 0,019 | 0,000 | 0,049 |
| rs4512969 | 13 | 38150669 | 3,906 | 3,134 | 2,767 | 2,757 | 0,0001573 | 0,0577619 | 0,000 | 0,013 | 0,000 | 0,049 |
| rs869096 | 6 | 157959413 | 3,447 | -3,137 | 2,435 | 2,440 | 0,0008195 | 0,0930783 | 0,003 | 0,041 | 0,000 | 0,049 |
| rs55729347 | 7 | 67201924 | 3,545 | -3,137 | 2,503 | 2,510 | 0,000645 | 0,084127 | 0,002 | 0,033 | 0,000 | 0,049 |
| rs78278528 | 10 | 18976861 | 3,623 | 3,136 | 2,566 | 2,559 | 0,00045 | 0,078241 | 0,001 | 0,027 | 0,000 | 0,049 |
| rs1602967* | 7 | 113596013 | 3,359 | 3,137 | 2,378 | 2,373 | 0,001191 | 0,1024794 | 0,004 | 0,050 | 0,000 | 0,050 |

*Loci within statistically significant correlated regions between SCZ and AF using LAVA

**Table S9. LD-independent SNPs targetting horizontal pleiotropic loci between schizophrenia and cannabis use disorder.**

| SNP | CHR | BP | r | angle | z.scz | z.canud | pval.scz | pval.canud | r.pval | r.qval | theta.pval | theta.qval | Previously reported at Johnson et al., 2024 |
|---|---|---|---|---|---|---|---|---|---|---|---|---|---|
| rs10753623 | 1 | 163714218 | 4,865 | -3,036 | 3,347 | -3,530 | 2,028E-05 | 0,000197 | 0,000 | 0,004 | 0,002 | 0,040 | No |
| rs10927025 | 1 | 243639218 | 4,514 | 3,124 | -3,207 | -3,178 | 4,846E-06 | 0,0002143 | 0,000 | 0,016 | 0,000 | 0,025 | No |
| rs11243587 | 9 | 134861101 | 4,212 | -3,117 | -2,960 | -2,997 | 6,265E-06 | 0,0002159 | 0,000 | 0,040 | 0,001 | 0,028 | Yes |
| rs113623975* | 8 | 27412605 | 5,000 | -3,123 | 3,519 | 3,551 | 1,063E-11 | 9,485E-10 | 0,000 | 0,003 | 0,000 | 0,025 | No |
| rs11665242 | 18 | 50907127 | 4,148 | 3,127 | 2,943 | 2,922 | 6,096E-06 | 0,000123 | 0,000 | 0,049 | 0,001 | 0,025 | No |
| rs11667321 | 19 | 49250538 | 4,737 | 3,016 | 3,452 | 3,243 | 1,752E-06 | 0,00005348 | 0,000 | 0,007 | 0,003 | 0,047 | No |
| rs11669242 | 19 | 31087575 | 4,373 | -3,073 | -3,039 | 3,144 | 9,305E-05 | 0,000961 | 0,000 | 0,024 | 0,002 | 0,042 | No |
| rs11756267 | 6 | 33431516 | 4,622 | -3,130 | 3,259 | 3,278 | 2,214E-05 | 0,0004853 | 0,000 | 0,011 | 0,000 | 0,025 | No |
| rs12046000 | 1 | 91192396 | 4,485 | -3,135 | -3,167 | -3,176 | 2,939E-05 | 0,0004486 | 0,000 | 0,017 | 0,000 | 0,025 | No |
| rs12419325 | 11 | 28580338 | 4,743 | 3,078 | 3,407 | 3,300 | 4,258E-06 | 0,0001847 | 0,000 | 0,007 | 0,001 | 0,032 | Yes |
| rs12575544* | 11 | 112918985 | 4,272 | 3,120 | -3,037 | 3,005 | 4,678E-07 | 0,00002824 | 0,000 | 0,033 | 0,001 | 0,027 | No |
| rs1265047 | 6 | 31081464 | 5,806 | 3,021 | 4,228 | 3,980 | 1,465E-05 | 0,0005218 | 0,000 | 0,000 | 0,001 | 0,025 | No |
| rs13025111 | 2 | 185797159 | 4,194 | -3,077 | 2,917 | 3,013 | 3,58E-07 | 0,00003565 | 0,000 | 0,042 | 0,002 | 0,044 | No |
| rs1372072 | 3 | 16955259 | 4,141 | -3,068 | -2,874 | 2,982 | 3,481E-09 | 1,597E-06 | 0,000 | 0,050 | 0,003 | 0,050 | No |
| rs1474011 | 2 | 146118069 | 4,559 | 3,125 | 3,237 | 3,210 | 0,0001115 | 0,002353 | 0,000 | 0,014 | 0,000 | 0,025 | No |
| rs1611627 | 6 | 29797782 | 5,385 | -3,090 | -3,758 | -3,857 | 1,597E-05 | 0,0004283 | 0,000 | 0,001 | 0,000 | 0,025 | No |
| rs16892543 | 8 | 89340162 | 4,504 | -3,127 | -3,174 | 3,196 | 3,901E-08 | 5,808E-06 | 0,000 | 0,016 | 0,000 | 0,025 | No |
| rs17402063 | 2 | 60527297 | 5,035 | -3,116 | -3,537 | -3,583 | 0,0001021 | 0,0009132 | 0,000 | 0,002 | 0,000 | 0,025 | No |
| rs1759854 | 9 | 111675333 | 4,638 | 3,026 | 3,373 | 3,183 | 1,807E-05 | 0,0004166 | 0,000 | 0,011 | 0,003 | 0,048 | No |
| rs17651507 | 17 | 44059010 | 4,240 | -3,141 | -2,998 | 2,998 | 9,412E-05 | 0,001548 | 0,000 | 0,037 | 0,000 | 0,025 | No |
| rs1873304 | 10 | 106535696 | 4,149 | 3,124 | 2,947 | -2,921 | 4,047E-06 | 0,0001194 | 0,000 | 0,049 | 0,001 | 0,026 | No |
| rs1887402 | 1 | 44036085 | 7,103 | -2,925 | 4,743 | 5,287 | 3,608E-06 | 0,0001323 | 0,000 | 0,000 | 0,000 | 0,025 | No |
| rs1917881 | 1 | 73313762 | 4,572 | -3,083 | -3,185 | -3,280 | 2,31E-09 | 2,952E-06 | 0,000 | 0,013 | 0,001 | 0,033 | No |
| rs200451 | 1 | 6721234 | 4,831 | -3,079 | 3,362 | 3,469 | 7,05E-08 | 7,196E-06 | 0,000 | 0,005 | 0,001 | 0,031 | Yes |
| rs2129292 | 4 | 102991139 | 4,198 | 3,069 | -3,022 | 2,914 | 9,029E-05 | 0,0009496 | 0,000 | 0,042 | 0,003 | 0,048 | No |
| rs2396937 | 5 | 3405541 | 4,286 | 3,120 | -3,047 | 3,014 | 2,802E-05 | 0,0003941 | 0,000 | 0,032 | 0,001 | 0,026 | No |
| rs256373 | 5 | 60501595 | 4,630 | -3,102 | 3,241 | 3,306 | 3,171E-08 | 6,659E-06 | 0,000 | 0,011 | 0,001 | 0,029 | No |
| rs28791500 | 4 | 118771886 | 4,266 | 3,139 | 3,019 | 3,014 | 3,228E-06 | 0,0002308 | 0,000 | 0,034 | 0,000 | 0,025 | No |
| rs2906458 | 1 | 44336389 | 4,397 | -3,077 | -3,058 | 3,159 | 2,721E-05 | 0,0005178 | 0,000 | 0,022 | 0,002 | 0,040 | No |
| rs34453863 | 6 | 26628005 | 4,394 | -3,083 | 3,061 | -3,152 | 2,754E-06 | 0,00005633 | 0,000 | 0,022 | 0,002 | 0,037 | No |
| rs35225200 | 4 | 103146888 | 5,476 | -3,136 | -3,867 | -3,878 | 8,752E-19 | 2,504E-06 | 0,000 | 0,000 | 0,000 | 0,025 | Yes |
| rs35545554 | 19 | 19715613 | 5,763 | 3,132 | 4,085 | 4,065 | 8,967E-05 | 0,001666 | 0,000 | 0,000 | 0,000 | 0,025 | Yes |
| rs4073950 | 2 | 22463414 | 4,266 | 3,081 | 3,062 | 2,970 | 2,427E-06 | 0,0001175 | 0,000 | 0,034 | 0,002 | 0,042 | No |
| rs4128930 | 6 | 88583388 | 4,233 | -3,062 | 2,933 | 3,052 | 3,019E-05 | 0,0005268 | 0,000 | 0,038 | 0,003 | 0,049 | No |
| rs4702 | 15 | 91426560 | 4,570 | -3,129 | 3,221 | 3,242 | 2,794E-21 | 1,673E-07 | 0,000 | 0,014 | 0,000 | 0,025 | No |
| rs4713630 | 6 | 33472941 | 5,481 | -3,141 | 3,875 | 3,876 | 5,039E-05 | 0,0008022 | 0,000 | 0,000 | 0,000 | 0,017 | No |
| rs4743195 | 9 | 101034018 | 4,714 | -3,105 | 3,303 | -3,363 | 2,276E-05 | 0,0004791 | 0,000 | 0,008 | 0,001 | 0,027 | No |
| rs4807003 | 19 | 4957133 | 4,624 | -3,109 | -3,243 | -3,296 | 1,734E-06 | 0,0001988 | 0,000 | 0,011 | 0,001 | 0,027 | Yes |
| rs55952530 | 15 | 78917316 | 4,555 | 3,061 | -3,285 | 3,155 | 2,517E-05 | 0,0004088 | 0,000 | 0,014 | 0,002 | 0,041 | No |
| rs56114371 | 6 | 27274834 | 4,542 | 3,068 | -3,270 | -3,152 | 5,946E-37 | 6,035E-07 | 0,000 | 0,015 | 0,002 | 0,039 | No |
| rs563722 | 11 | 74128941 | 4,492 | 3,136 | 3,181 | 3,172 | 0,0001535 | 0,002137 | 0,000 | 0,017 | 0,000 | 0,025 | No |
| rs600011 | 3 | 107295392 | 4,773 | -3,102 | 3,342 | 3,408 | 1,468E-06 | 0,0001115 | 0,000 | 0,006 | 0,001 | 0,027 | No |
| rs6088520 | 20 | 33132364 | 5,707 | -3,093 | 3,986 | -4,084 | 0,0001021 | 0,001664 | 0,000 | 0,000 | 0,000 | 0,025 | No |
| rs62012056 | 15 | 82539619 | 4,666 | 3,103 | 3,331 | 3,268 | 2,622E-05 | 0,0006818 | 0,000 | 0,010 | 0,001 | 0,028 | No |

| | | | | | | | | | | | | | |
|---|---|---|---|---|---|---|---|---|---|---|---|---|---|
| rs62055046 | 16 | 71396960 | 4,158 | 3,135 | -2,946 | -2,935 | 6,166E-06 | 0,0002113 | 0,000 | 0,047 | 0,000 | 0,025 | No |
| rs6476547 | 9 | 36311270 | 10,032 | 1,650 | 9,190 | 4,022 | 6,667E-06 | 0,0002643 | 0,000 | 0,000 | 0,000 | 0,025 | No |
| rs6589386* | 11 | 113443753 | 4,446 | 3,134 | 3,150 | 3,138 | 1,352E-10 | 1,096E-06 | 0,000 | 0,019 | 0,000 | 0,025 | No |
| rs67959840 | 6 | 94048610 | 4,182 | -3,126 | 2,945 | 2,969 | 0,0001434 | 0,00158 | 0,000 | 0,044 | 0,001 | 0,025 | No |
| rs681846 | 13 | 56219314 | 4,231 | -3,131 | -2,984 | 3,000 | 4,058E-05 | 0,0005171 | 0,000 | 0,038 | 0,000 | 0,025 | No |
| rs6852412 | 4 | 143795347 | 4,393 | -3,083 | 3,061 | -3,152 | 4,556E-06 | 0,0001647 | 0,000 | 0,022 | 0,002 | 0,037 | No |
| rs6975268* | 7 | 104915608 | 4,176 | -3,099 | 2,921 | 2,985 | 2,355E-06 | 0,0001314 | 0,000 | 0,045 | 0,002 | 0,035 | No |
| rs71558326 | 6 | 108971060 | 4,758 | -3,043 | -3,281 | -3,447 | 2,716E-05 | 0,0004978 | 0,000 | 0,007 | 0,002 | 0,041 | No |
| rs7196177* | 16 | 83166796 | 4,284 | 3,058 | 3,092 | 2,965 | 3,017E-05 | 0,0006152 | 0,000 | 0,032 | 0,003 | 0,049 | No |
| rs72926963* | 18 | 53076790 | 4,490 | 3,136 | 3,179 | 3,171 | 0,0000118 | 0,0005081 | 0,000 | 0,017 | 0,000 | 0,025 | No |
| rs76582536 | 7 | 24548340 | 6,578 | 2,806 | -5,024 | -4,245 | 0,0001298 | 0,001716 | 0,000 | 0,000 | 0,001 | 0,028 | No |
| rs78791936 | 13 | 66427641 | 4,142 | -3,090 | 2,891 | 2,966 | 3,037E-05 | 0,0006519 | 0,000 | 0,050 | 0,002 | 0,040 | No |
| rs79367267 | 1 | 243689543 | 4,217 | -3,142 | -2,982 | -2,982 | 5,985E-06 | 0,00022 | 0,000 | 0,040 | 0,000 | 0,013 | No |
| rs9272550 | 6 | 32606970 | 8,212 | 2,384 | -6,795 | 4,610 | 9,993E-07 | 0,0001469 | 0,000 | 0,000 | 0,000 | 0,025 | No |
| rs9380326 | 6 | 32798283 | 4,302 | 3,109 | 3,067 | -3,018 | 0,0001022 | 0,0009633 | 0,000 | 0,030 | 0,001 | 0,030 | No |
| rs9420 | 11 | 57510294 | 6,127 | 2,970 | 4,514 | 4,142 | 6,647E-09 | 2,323E-06 | 0,000 | 0,000 | 0,001 | 0,027 | Yes |
| rs9784441 | 4 | 34146884 | 4,275 | 3,133 | -3,029 | -3,016 | 3,594E-06 | 0,000119 | 0,000 | 0,033 | 0,000 | 0,025 | No |
| rs9788721 | 15 | 78802869 | 4,667 | -3,119 | 3,281 | -3,318 | 3,487E-07 | 0,00001883 | 0,000 | 0,010 | 0,001 | 0,025 | No |
| rs35277073 | 11 | 113350620 | 6,347 | 2,342 | 5,290 | 3,507 | 1,468E-13 | 0,00002824 | 0,000 | 0,000 | 0,003 | 0,048 | No |
| rs6738845 | 2 | 22748926 | 4,835 | -3,122 | 3,401 | 3,436 | 1,108E-06 | 0,0000628 | 0,000 | 0,005 | 0,000 | 0,025 | No |

*Loci within statistically significant correlated regions between SCZ and CanUD using LAVA

**Table S10. LD-independent SNPs targetting horizontal pleiotropic loci between schizophrenia and opioid use disorder.**

| SNP | CHR | BP | r | angle | z.scz | z.oud | pval.scz | pval.oud | r.pval | r.qval | theta.pval | theta.qval | Previously reported at Holen et al., 2023 |
|---|---|---|---|---|---|---|---|---|---|---|---|---|---|
| rs4129585* | 8 | 143312933 | 8,478 | 3,137 | 6,003 | 5,987 | 5,109E-18 | 3,728E-10 | 0,000 | 0,000 | 0,000 | 0,007 | No |
| rs2946004 | 11 | 45997805 | 4,212 | -3,142 | 2,978 | 2,978 | 2,024E-05 | 0,0012567 | 0,000 | 0,041 | 0,000 | 0,013 | No |

*Loci within statistically significant correlated regions between SCZ and OUD using LAVA

**Table S11. LD-independent SNPs targetting horizontal pleiotropic loci between schizophrenia and problematic alcohol use.**

| SNP | CHR | BP | r | angle | z.scz | z.pau | pval.scz | pval.pau | r.pval | r.qval | theta.pval | theta.qval | Previously reported at Wiström et al., 2022 |
|---|---|---|---|---|---|---|---|---|---|---|---|---|---|
| rs10128780 | 12 | 110323951 | 4,190 | -3,126 | 2,951 | 2,974 | 2,49E-05 | 0,000173 | 0,000 | 0,035 | 0,001 | 0,015 | No |
| rs10163350 | 16 | 7349994 | 4,368 | -3,020 | -2,994 | -3,181 | 1,86E-05 | 6,17E-05 | 0,000 | 0,020 | 0,004 | 0,032 | No |
| rs1056668 | 6 | 26510605 | 4,430 | -3,123 | -3,118 | -3,147 | 8,87E-06 | 7,06E-05 | 0,000 | 0,016 | 0,001 | 0,015 | No |
| rs10835372 | 11 | 28643913 | 5,317 | -2,491 | 3,101 | 4,319 | 6E-06 | 7,95E-08 | 0,000 | 0,000 | 0,007 | 0,044 | No |
| rs10838601 | 11 | 46392825 | 6,114 | 2,901 | -4,576 | -4,055 | 9,34E-11 | 2,45E-07 | 0,000 | 0,000 | 0,001 | 0,016 | Yes |
| rs10883553 | 10 | 102635475 | 4,427 | -2,879 | -2,918 | 3,329 | 0,000257 | 0,000178 | 0,000 | 0,017 | 0,007 | 0,047 | No |
| rs11210201 | 1 | 73766431 | 7,303 | 2,348 | -6,081 | -4,045 | 1,55E-17 | 1,44E-07 | 0,000 | 0,000 | 0,001 | 0,015 | No |
| rs11214489 | 11 | 112975934 | 4,241 | -3,059 | -2,936 | -3,060 | 2,61E-05 | 0,000115 | 0,000 | 0,030 | 0,003 | 0,028 | No |
| rs1125000* | 6 | 26287256 | 5,977 | 3,067 | -4,304 | 4,147 | 4,17E-08 | 3,81E-06 | 0,000 | 0,000 | 0,000 | 0,015 | No |
| rs114993052 | 1 | 91190855 | 4,111 | 3,063 | 2,963 | 2,849 | 2,46E-05 | 0,000309 | 0,000 | 0,045 | 0,003 | 0,030 | No |
| rs11600358 | 11 | 130861244 | 4,232 | 2,958 | -3,127 | 2,852 | 6,96E-05 | 0,001548 | 0,000 | 0,031 | 0,006 | 0,043 | No |
| rs11649759 | 17 | 2019048 | 4,323 | 3,119 | 3,074 | -3,040 | 0,000101 | 0,000689 | 0,000 | 0,023 | 0,001 | 0,015 | No |
| rs11654265* | 17 | 78602697 | 4,498 | -3,141 | 3,180 | 3,181 | 5,97E-06 | 5,85E-05 | 0,000 | 0,013 | 0,000 | 0,015 | No |
| rs117064001 | 22 | 42009666 | 4,697 | 3,042 | 3,403 | 3,238 | 1,18E-06 | 4,08E-05 | 0,000 | 0,006 | 0,002 | 0,024 | No |
| rs11755421 | 6 | 33715323 | 4,778 | -3,105 | -3,347 | -3,410 | 1,68E-06 | 1,68E-05 | 0,000 | 0,005 | 0,001 | 0,015 | No |
| rs11807834 | 1 | 230272624 | 5,062 | 2,755 | -3,908 | -3,217 | 2,98E-08 | 3,87E-05 | 0,000 | 0,001 | 0,006 | 0,043 | No |
| rs11859355 | 16 | 13580639 | 4,091 | 3,041 | 2,964 | 2,819 | 2,16E-05 | 0,000355 | 0,000 | 0,048 | 0,004 | 0,033 | No |
| rs1198594 | 1 | 98540744 | 6,781 | 2,041 | -5,917 | 3,311 | 1,4E-14 | 0,00045 | 0,000 | 0,000 | 0,003 | 0,029 | No |
| rs12331633 | 4 | 30832319 | 4,201 | 3,137 | 2,974 | 2,968 | 2,09E-05 | 0,000177 | 0,000 | 0,034 | 0,000 | 0,015 | No |
| rs12575367 | 11 | 17360590 | 4,309 | 3,083 | 3,091 | 3,002 | 9,34E-06 | 0,000145 | 0,000 | 0,024 | 0,002 | 0,022 | No |
| rs12892186 | 14 | 104318787 | 6,426 | -3,135 | -4,537 | -4,551 | 7,29E-11 | 8,99E-09 | 0,000 | 0,000 | 0,000 | 0,013 | Yes |
| rs12918180 | 16 | 6564548 | 4,861 | -3,052 | 3,360 | 3,513 | 1,48E-06 | 9,51E-06 | 0,000 | 0,003 | 0,002 | 0,020 | Yes |
| rs13018418 | 2 | 175023368 | 4,513 | -3,129 | -3,181 | 3,201 | 6,22E-05 | 0,000346 | 0,000 | 0,012 | 0,000 | 0,015 | No |
| rs13107325* | 4 | 103188709 | 14,327 | -2,285 | -7,746 | 12,052 | 2,9E-21 | 3,43E-43 | 0,000 | 0,000 | 0,000 | 0,000 | No |
| rs132548 | 22 | 29317559 | 4,319 | -3,081 | 3,007 | 3,100 | 1,57E-05 | 9,22E-05 | 0,000 | 0,024 | 0,002 | 0,023 | No |
| rs13409316 | 2 | 155937322 | 4,509 | -3,021 | -3,091 | 3,283 | 9,45E-05 | 0,000231 | 0,000 | 0,012 | 0,003 | 0,030 | No |
| rs1366840 | 2 | 185767854 | 5,108 | -3,115 | 3,588 | 3,636 | 2,79E-07 | 4,43E-06 | 0,000 | 0,001 | 0,000 | 0,015 | No |
| rs1406491* | 8 | 64633425 | 4,536 | -3,048 | 3,132 | 3,282 | 7,53E-06 | 3,54E-05 | 0,000 | 0,011 | 0,002 | 0,025 | No |
| rs144689944* | 11 | 25122748 | 4,152 | -2,947 | -2,789 | 3,075 | 0,000459 | 0,000549 | 0,000 | 0,040 | 0,007 | 0,047 | No |
| rs147487154 | 8 | 70990615 | 4,246 | -3,076 | -2,953 | 3,051 | 0,000192 | 0,000636 | 0,000 | 0,030 | 0,002 | 0,025 | No |
| rs1561296 | 2 | 201308117 | 4,835 | -3,092 | 3,376 | 3,461 | 1,16E-06 | 1,26E-05 | 0,000 | 0,004 | 0,001 | 0,016 | No |
| rs1630095* | 17 | 43750172 | 5,529 | 3,141 | -3,910 | -3,909 | 2,42E-08 | 7,86E-07 | 0,000 | 0,000 | 0,000 | 0,007 | No |
| rs1632065 | 5 | 153525007 | 4,501 | 3,104 | -3,213 | -3,153 | 4,77E-06 | 6,67E-05 | 0,000 | 0,013 | 0,001 | 0,016 | No |
| rs1647394 | 11 | 57418588 | 4,320 | 3,131 | -3,063 | -3,046 | 1,04E-05 | 0,000118 | 0,000 | 0,024 | 0,000 | 0,015 | Yes |
| rs1658810 | 2 | 200816382 | 6,258 | 2,183 | 5,349 | 3,248 | 6,51E-14 | 2,07E-05 | 0,000 | 0,000 | 0,004 | 0,032 | No |
| rs167238 | 8 | 130938211 | 4,744 | -3,124 | 3,339 | 3,369 | 1,87E-06 | 2,09E-05 | 0,000 | 0,005 | 0,000 | 0,015 | No |
| rs16884402 | 4 | 31141947 | 4,752 | -3,058 | 3,289 | 3,430 | 2,36E-06 | 1,54E-05 | 0,000 | 0,005 | 0,002 | 0,020 | No |
| rs16947293 | 18 | 27472712 | 4,281 | 3,052 | -3,094 | -2,959 | 9,4E-06 | 0,000178 | 0,000 | 0,027 | 0,003 | 0,029 | No |
| rs17119214 | 10 | 107371475 | 4,243 | -2,934 | 2,841 | 3,152 | 4,03E-05 | 7,5E-05 | 0,000 | 0,030 | 0,007 | 0,047 | No |
| rs17479770 | 2 | 225335814 | 4,474 | -3,114 | 3,142 | 3,185 | 6,47E-06 | 5,79E-05 | 0,000 | 0,014 | 0,001 | 0,015 | No |
| rs17651504* | 11 | 28460189 | 4,806 | -2,878 | -3,167 | -3,615 | 4,75E-06 | 5,75E-06 | 0,000 | 0,004 | 0,005 | 0,038 | No |
| rs1766377 | 1 | 49917084 | 4,120 | -2,972 | 2,787 | 3,035 | 6,65E-05 | 0,000136 | 0,000 | 0,044 | 0,007 | 0,045 | No |
| rs17759659 | 18 | 60958644 | 4,547 | 3,135 | -3,221 | 3,210 | 4,62E-05 | 0,000338 | 0,000 | 0,011 | 0,000 | 0,015 | No |
| rs1873309 | 10 | 106579771 | 4,116 | 3,082 | 2,954 | 2,866 | 2,59E-05 | 0,000285 | 0,000 | 0,045 | 0,002 | 0,026 | No |

| | | | | | | | | | | | | | |
|---|---|---|---|---|---|---|---|---|---|---|---|---|---|
| rs1875883 | 15 | 76638231 | 4,254 | -3,123 | -2,994 | -3,022 | 1,7E-05 | 0,000136 | 0,000 | 0,029 | 0,001 | 0,015 | No |
| rs1914391 | 7 | 71690601 | 4,645 | -3,109 | 3,258 | 3,311 | 3,24E-06 | 2,92E-05 | 0,000 | 0,007 | 0,001 | 0,015 | No |
| rs2023497 | 6 | 29551333 | 4,251 | 2,946 | 3,149 | 2,855 | 7,38E-06 | 0,000285 | 0,000 | 0,029 | 0,007 | 0,045 | No |
| rs211456 | 6 | 33389381 | 4,903 | 3,097 | 3,506 | 3,428 | 4,81E-07 | 1,44E-05 | 0,000 | 0,003 | 0,001 | 0,015 | No |
| rs2121637 | 8 | 94175169 | 4,307 | -2,979 | -2,919 | 3,167 | 0,000249 | 0,000377 | 0,000 | 0,025 | 0,005 | 0,039 | No |
| rs2159102 | 12 | 2288945 | 6,643 | 3,021 | -4,837 | -4,553 | 4,46E-12 | 7,76E-09 | 0,000 | 0,000 | 0,000 | 0,015 | No |
| rs2283431 | 15 | 89838045 | 4,150 | 3,074 | 2,984 | -2,885 | 0,00016 | 0,0013 | 0,000 | 0,040 | 0,003 | 0,026 | No |
| rs2304682 | 2 | 27307769 | 4,087 | 3,106 | -2,915 | -2,865 | 3,04E-05 | 0,000291 | 0,000 | 0,049 | 0,001 | 0,018 | No |
| rs2371263* | 2 | 212236088 | 4,160 | 3,084 | 2,984 | 2,899 | 2,01E-05 | 0,000243 | 0,000 | 0,039 | 0,002 | 0,024 | No |
| rs2485653 | 1 | 44594966 | 4,525 | -3,069 | 3,141 | 3,257 | 6,91E-06 | 4,02E-05 | 0,000 | 0,012 | 0,002 | 0,021 | No |
| rs2514218* | 11 | 113392994 | 9,214 | -2,407 | 5,216 | 7,596 | 1,35E-14 | 4,39E-21 | 0,000 | 0,000 | 0,000 | 0,015 | No |
| rs2524227* | 6 | 31302563 | 4,296 | -3,124 | 3,024 | 3,051 | 1,48E-05 | 0,000117 | 0,000 | 0,025 | 0,001 | 0,015 | No |
| rs2578377 | 5 | 153413390 | 4,212 | -3,137 | 2,975 | 2,982 | 2,01E-05 | 0,000165 | 0,000 | 0,033 | 0,000 | 0,015 | No |
| rs2583636 | 4 | 83153179 | 4,240 | 2,900 | 3,173 | 2,812 | 6,77E-06 | 0,000345 | 0,000 | 0,030 | 0,008 | 0,049 | No |
| rs2678890 | 2 | 58164435 | 7,700 | 3,140 | -5,447 | -5,443 | 7,53E-15 | 6,12E-12 | 0,000 | 0,000 | 0,000 | 0,003 | No |
| rs2693672* | 14 | 99741074 | 4,585 | -3,034 | 3,153 | 3,328 | 6,2E-06 | 2,77E-05 | 0,000 | 0,009 | 0,003 | 0,027 | No |
| rs28545550 | 16 | 13741891 | 4,568 | -3,108 | -3,203 | -3,257 | 4,06E-06 | 3,93E-05 | 0,000 | 0,010 | 0,001 | 0,015 | Yes |
| rs2909449 | 2 | 162866338 | 4,273 | 3,120 | 3,037 | 3,005 | 1,34E-05 | 0,000145 | 0,000 | 0,027 | 0,001 | 0,015 | No |
| rs3129081 | 6 | 29604683 | 4,152 | 3,129 | 2,945 | 2,927 | 2,6E-05 | 0,000217 | 0,000 | 0,040 | 0,000 | 0,015 | No |
| rs34169640 | 12 | 53838325 | 4,233 | -3,092 | -2,956 | -3,030 | 2,32E-05 | 0,000132 | 0,000 | 0,031 | 0,002 | 0,020 | No |
| rs35622757 | 7 | 109993384 | 4,149 | -2,972 | 2,807 | 3,056 | 5,75E-05 | 0,000122 | 0,000 | 0,040 | 0,006 | 0,044 | No |
| rs41396445* | 18 | 53242137 | 5,552 | 3,112 | -3,955 | -3,897 | 1,42E-08 | 8,32E-07 | 0,000 | 0,000 | 0,000 | 0,015 | No |
| rs442972 | 21 | 40534757 | 4,943 | -3,088 | -3,448 | 3,542 | 1,47E-05 | 7,37E-05 | 0,000 | 0,002 | 0,001 | 0,015 | No |
| rs4438849 | 5 | 103895102 | 4,170 | 2,935 | -3,097 | -2,792 | 1,11E-05 | 0,000385 | 0,000 | 0,038 | 0,008 | 0,048 | No |
| rs456917 | 6 | 66172505 | 4,123 | -3,113 | 2,895 | -2,936 | 0,000248 | 0,001026 | 0,000 | 0,044 | 0,001 | 0,016 | No |
| rs4702 | 15 | 91426560 | 7,512 | 1,713 | 6,833 | 3,120 | 2,79E-21 | 2,51E-05 | 0,000 | 0,000 | 0,004 | 0,035 | No |
| rs4743195* | 9 | 101034018 | 4,144 | 3,118 | -2,948 | -2,913 | 2,28E-05 | 0,000231 | 0,000 | 0,041 | 0,001 | 0,016 | No |
| rs4787488 | 16 | 29995803 | 6,502 | -2,902 | 4,315 | 4,864 | 5,89E-10 | 1,02E-09 | 0,000 | 0,000 | 0,001 | 0,015 | Yes |
| rs4936277* | 11 | 113431960 | 8,437 | -2,057 | 4,150 | 7,346 | 5,68E-10 | 1,45E-19 | 0,000 | 0,000 | 0,000 | 0,015 | Yes |
| rs4980502 | 11 | 63805335 | 4,214 | 2,902 | -3,153 | -2,796 | 7,45E-06 | 0,000372 | 0,000 | 0,033 | 0,009 | 0,050 | No |
| rs4987094* | 11 | 113215923 | 5,380 | 3,098 | -3,845 | -3,763 | 3,65E-08 | 1,94E-06 | 0,000 | 0,000 | 0,000 | 0,015 | No |
| rs57929572 | 3 | 71563561 | 5,274 | 2,387 | 4,362 | -2,964 | 1,57E-08 | 0,001333 | 0,000 | 0,001 | 0,008 | 0,048 | No |
| rs600011 | 3 | 107295392 | 4,604 | 2,960 | 3,400 | 3,104 | 1,47E-06 | 8,03E-05 | 0,000 | 0,009 | 0,004 | 0,035 | No |
| rs612823 | 11 | 133834104 | 6,227 | 2,283 | 5,240 | 3,364 | 2,49E-13 | 1,15E-05 | 0,000 | 0,000 | 0,006 | 0,040 | No |
| rs61733949 | 20 | 42088708 | 4,181 | 2,903 | 3,127 | 2,774 | 8,59E-06 | 0,000414 | 0,000 | 0,037 | 0,009 | 0,050 | No |
| rs62398834* | 5 | 152529901 | 4,314 | 3,035 | -3,131 | -2,968 | 7,2E-06 | 0,000168 | 0,000 | 0,024 | 0,004 | 0,031 | No |
| rs62406850 | 6 | 69946240 | 4,590 | 2,805 | -3,507 | 2,961 | 6,92E-06 | 0,001083 | 0,000 | 0,009 | 0,008 | 0,049 | No |
| rs643831* | 7 | 4190185 | 4,350 | 2,972 | -3,203 | 2,943 | 4,58E-05 | 0,001078 | 0,000 | 0,021 | 0,005 | 0,039 | No |
| rs6545798 | 2 | 60521311 | 4,286 | 3,139 | 3,033 | -3,028 | 0,000129 | 0,000718 | 0,000 | 0,026 | 0,000 | 0,015 | No |
| rs6546859 | 2 | 73842055 | 5,520 | -3,140 | -3,901 | -3,905 | 2,61E-08 | 8,04E-07 | 0,000 | 0,000 | 0,000 | 0,014 | No |
| rs6577497 | 1 | 8605667 | 4,283 | 2,903 | -3,204 | -2,843 | 5E-06 | 0,000297 | 0,000 | 0,026 | 0,008 | 0,048 | No |
| rs660745 | 19 | 49219459 | 4,572 | -3,129 | -3,223 | -3,243 | 4,08E-06 | 4,2E-05 | 0,000 | 0,010 | 0,000 | 0,015 | No |
| rs6698162 | 1 | 66319371 | 5,479 | -3,055 | -3,789 | -3,958 | 6,18E-08 | 6,1E-07 | 0,000 | 0,000 | 0,001 | 0,015 | Yes |
| rs67040074 | 2 | 229017414 | 4,515 | -3,090 | 3,151 | 3,233 | 6,73E-06 | 4,52E-05 | 0,000 | 0,012 | 0,001 | 0,018 | No |
| rs6771755* | 3 | 196025846 | 4,181 | 3,130 | -2,964 | -2,948 | 2,11E-05 | 0,000194 | 0,000 | 0,036 | 0,000 | 0,015 | No |
| rs6783882 | 3 | 16730790 | 4,314 | 3,092 | 3,088 | 3,012 | 1,12E-05 | 0,000139 | 0,000 | 0,024 | 0,002 | 0,020 | No |
| rs6893642 | 5 | 60117723 | 5,037 | -2,947 | 3,384 | 3,731 | 1,14E-06 | 2,74E-06 | 0,000 | 0,002 | 0,003 | 0,029 | No |

| | | | | | | | | | | | | | |
|---|---|---|---|---|---|---|---|---|---|---|---|---|---|
| rs7195496 | 16 | 58670346 | 4,094 | 3,132 | -2,901 | -2,888 | 3,54E-05 | 0,000262 | 0,000 | 0,048 | 0,000 | 0,015 | No |
| rs72687340 | 8 | 143343260 | 5,038 | 3,099 | 3,600 | 3,525 | 2,52E-07 | 8,25E-06 | 0,000 | 0,002 | 0,001 | 0,015 | No |
| rs72768621 | 16 | 24687305 | 4,152 | -2,923 | 2,771 | 3,092 | 6,37E-05 | 0,000103 | 0,000 | 0,040 | 0,008 | 0,049 | No |
| rs727690* | 14 | 60157360 | 4,256 | -3,137 | 3,006 | 3,013 | 1,89E-05 | 0,000141 | 0,000 | 0,029 | 0,000 | 0,015 | No |
| rs72843784 | 6 | 26498758 | 9,777 | 1,564 | 9,039 | 3,727 | 6,28E-36 | 3,45E-07 | 0,000 | 0,000 | 0,001 | 0,015 | No |
| rs72928914* | 18 | 53108857 | 4,208 | 3,017 | 3,067 | 2,881 | 1,26E-05 | 0,000258 | 0,000 | 0,033 | 0,005 | 0,036 | No |
| rs73169135 | 22 | 42656681 | 4,337 | 3,055 | 3,132 | 3,000 | 7,2E-06 | 0,000145 | 0,000 | 0,022 | 0,003 | 0,027 | No |
| rs7320347 | 13 | 89030964 | 4,421 | -2,971 | -2,990 | -3,256 | 1,81E-05 | 4,21E-05 | 0,000 | 0,017 | 0,005 | 0,038 | No |
| rs7323748 | 13 | 115037013 | 4,125 | 2,974 | 3,037 | 2,792 | 1,59E-05 | 0,000393 | 0,000 | 0,043 | 0,007 | 0,045 | No |
| rs7329530 | 13 | 97192839 | 4,232 | 3,125 | 3,005 | 2,980 | 1,82E-05 | 0,000165 | 0,000 | 0,031 | 0,001 | 0,015 | No |
| rs748832 | 3 | 16851202 | 5,541 | 3,078 | -3,979 | -3,855 | 1,32E-08 | 1,06E-06 | 0,000 | 0,000 | 0,001 | 0,015 | Yes |
| rs75070454 | 4 | 55281935 | 4,268 | 2,922 | 3,179 | 2,847 | 6,66E-06 | 0,000292 | 0,000 | 0,028 | 0,008 | 0,048 | No |
| rs7550959 | 1 | 97926839 | 5,491 | -2,397 | 3,097 | 4,534 | 5,42E-06 | 1,86E-08 | 0,000 | 0,000 | 0,008 | 0,048 | No |
| rs7591789 | 2 | 208501738 | 4,342 | -3,002 | 2,961 | 3,176 | 2,2E-05 | 6,42E-05 | 0,000 | 0,022 | 0,004 | 0,035 | No |
| rs76537915 | 1 | 115636496 | 4,693 | 3,125 | 3,332 | -3,305 | 2,61E-05 | 0,000224 | 0,000 | 0,006 | 0,000 | 0,015 | No |
| rs7666854 | 4 | 143891865 | 5,074 | -3,066 | 3,520 | 3,655 | 4,9E-07 | 4,07E-06 | 0,000 | 0,001 | 0,001 | 0,016 | No |
| rs76900682 | 1 | 174064560 | 4,130 | -3,127 | -2,909 | 2,931 | 0,000235 | 0,001051 | 0,000 | 0,043 | 0,001 | 0,015 | No |
| rs77955054 | 4 | 102020946 | 4,606 | 3,127 | -3,268 | 3,245 | 3,52E-05 | 0,00029 | 0,000 | 0,009 | 0,000 | 0,015 | No |
| rs78125189 | 8 | 143333888 | 4,386 | -3,010 | -2,998 | -3,201 | 1,71E-05 | 5,58E-05 | 0,000 | 0,019 | 0,004 | 0,032 | No |
| rs80238644* | 14 | 99690466 | 4,393 | -3,130 | -3,097 | -3,115 | 9,09E-06 | 8,31E-05 | 0,000 | 0,019 | 0,000 | 0,015 | No |
| rs839772 | 1 | 43873483 | 4,563 | 3,024 | 3,320 | 3,130 | 2,5E-06 | 7,23E-05 | 0,000 | 0,010 | 0,003 | 0,029 | Yes |
| rs879048* | 11 | 27638934 | 5,018 | -2,775 | -3,208 | -3,858 | 3,03E-06 | 1,38E-06 | 0,000 | 0,002 | 0,006 | 0,041 | No |
| rs9277976 | 6 | 33292745 | 4,416 | -3,057 | -3,056 | -3,188 | 1,12E-05 | 5,86E-05 | 0,000 | 0,017 | 0,002 | 0,026 | No |
| rs929444 | 14 | 72440976 | 5,324 | 2,444 | -4,361 | -3,055 | 9,87E-10 | 7,65E-05 | 0,000 | 0,000 | 0,007 | 0,047 | No |
| rs9347755 | 6 | 163932583 | 4,356 | -3,141 | -3,080 | -3,080 | 1,09E-05 | 9,94E-05 | 0,000 | 0,021 | 0,000 | 0,008 | No |
| rs9449621 | 6 | 84178342 | 4,742 | 3,078 | -3,406 | 3,300 | 1,59E-05 | 0,000235 | 0,000 | 0,005 | 0,001 | 0,017 | No |
| rs9545073 | 13 | 79909007 | 6,352 | 2,897 | 4,758 | 4,208 | 1,02E-11 | 8,46E-08 | 0,000 | 0,000 | 0,001 | 0,015 | Yes |
| rs9562018 | 13 | 96713182 | 4,316 | 3,141 | -3,052 | -3,052 | 1,25E-05 | 0,000115 | 0,000 | 0,024 | 0,000 | 0,007 | No |
| rs959141 | 2 | 48505401 | 4,149 | 3,032 | 3,013 | -2,852 | 0,000131 | 0,001501 | 0,000 | 0,040 | 0,004 | 0,034 | No |
| rs9607805 | 22 | 41854446 | 4,293 | 3,109 | 3,060 | -3,011 | 0,000107 | 0,000779 | 0,000 | 0,026 | 0,001 | 0,016 | No |
| rs9649275 | 7 | 104935801 | 6,031 | 2,111 | 5,211 | 3,037 | 4,08E-13 | 6,45E-05 | 0,000 | 0,000 | 0,008 | 0,048 | No |
| rs9661233 | 1 | 173844051 | 4,102 | -3,083 | 2,858 | 2,943 | 3,96E-05 | 0,000206 | 0,000 | 0,046 | 0,002 | 0,025 | No |
| rs9817020 | 3 | 178969711 | 4,098 | 3,042 | 2,969 | -2,825 | 0,000159 | 0,001658 | 0,000 | 0,047 | 0,004 | 0,033 | No |
| rs9902772 | 17 | 7823486 | 4,087 | 3,018 | 2,977 | 2,799 | 2,25E-05 | 0,000386 | 0,000 | 0,049 | 0,005 | 0,037 | No |
| rs1611236 | 6 | 29748690 | 5,867 | 2,798 | 4,489 | -3,777 | 8,47E-09 | 3,08E-05 | 0,000 | 0,000 | 0,002 | 0,020 | No |
| rs3733197 | 4 | 102839287 | 6,058 | -2,633 | -3,706 | 4,792 | 3,76E-06 | 5,55E-08 | 0,000 | 0,000 | 0,003 | 0,027 | No |
| rs2514225 | 11 | 113396081 | 5,321 | -2,744 | 3,370 | 4,118 | 1,14E-06 | 2,64E-07 | 0,000 | 0,000 | 0,004 | 0,032 | No |

*Loci within statistically significant correlated regions between SCZ and PAU using LAVA

**Table S12. LD-independent SNPs targetting horizontal pleiotropic loci between schizophrenia and tobacco use disorder.**

| SNP | CHR | BP | r | angle | z.scz | z.tud | pval.scz | pval.tud | r.pval | r.qval | theta.pval | theta.qval | Previously reported at Johnson et al., 2024 |
|---|---|---|---|---|---|---|---|---|---|---|---|---|---|
| rs10159117 | 1 | 101284293 | 4,513 | -3,069 | 3,133 | 3,249 | 0,0000152 | 0,0001718 | 0,000 | 0,013 | 0,002 | 0,036 | No |
| rs10159191 | 1 | 191321186 | 4,122 | 3,138 | -2,917 | -2,912 | 5,792E-05 | 0,0007529 | 0,000 | 0,044 | 0,000 | 0,008 | No |
| rs10198333 | 2 | 60490773 | 4,745 | 3,119 | 3,374 | -3,336 | 7,008E-06 | 0,0002042 | 0,000 | 0,006 | 0,000 | 0,017 | No |
| rs10234862 | 7 | 110120191 | 4,304 | -3,132 | 3,036 | 3,050 | 3,128E-05 | 0,0004164 | 0,000 | 0,026 | 0,000 | 0,014 | No |
| rs10438243 | 14 | 104345822 | 5,421 | 3,124 | 3,850 | 3,816 | 1,032E-07 | 1,002E-05 | 0,000 | 0,000 | 0,000 | 0,008 | No |
| rs10823790 | 10 | 73338253 | 4,642 | -3,052 | -3,208 | -3,355 | 9,808E-06 | 0,0001049 | 0,000 | 0,009 | 0,002 | 0,037 | No |
| rs10922037 | 1 | 189390041 | 4,675 | -2,969 | -3,160 | 3,446 | 3,054E-05 | 0,0001214 | 0,000 | 0,008 | 0,004 | 0,049 | No |
| rs10970783 | 9 | 32112528 | 4,199 | -3,098 | 2,937 | 3,002 | 4,877E-05 | 0,0005167 | 0,000 | 0,035 | 0,002 | 0,035 | No |
| rs1109771 | 6 | 32187605 | 4,698 | -3,140 | 3,320 | -3,324 | 1,027E-05 | 0,0002152 | 0,000 | 0,007 | 0,000 | 0,004 | No |
| rs111334890 | 4 | 85607376 | 4,228 | -3,140 | -2,988 | 2,991 | 7,601E-05 | 0,0008663 | 0,000 | 0,032 | 0,000 | 0,007 | No |
| rs11192193 | 10 | 106569253 | 5,393 | 3,010 | 3,937 | 3,686 | 5,094E-08 | 0,0000195 | 0,000 | 0,000 | 0,001 | 0,031 | No |
| rs113706174 | 22 | 51145702 | 5,221 | -3,097 | 3,650 | 3,733 | 4,853E-07 | 1,572E-05 | 0,000 | 0,001 | 0,001 | 0,023 | No |
| rs11557154 | 9 | 34107505 | 4,384 | -3,073 | 3,046 | -3,153 | 5,208E-05 | 0,0004434 | 0,000 | 0,020 | 0,002 | 0,037 | No |
| rs11606722 | 11 | 1476662 | 4,384 | 3,048 | 3,172 | -3,027 | 2,643E-05 | 0,0007598 | 0,000 | 0,020 | 0,003 | 0,042 | No |
| rs11651922 | 17 | 30672417 | 4,611 | 3,045 | 3,338 | 3,181 | 4,355E-06 | 0,00023 | 0,000 | 0,010 | 0,002 | 0,039 | No |
| rs117079136 | 22 | 41757770 | 4,629 | -3,002 | -3,156 | 3,386 | 2,958E-05 | 0,0001603 | 0,000 | 0,009 | 0,003 | 0,045 | No |
| rs11783093 | 8 | 27425349 | 9,344 | -2,239 | 4,962 | 7,918 | 4,681E-12 | 9,219E-20 | 0,000 | 0,000 | 0,000 | 0,004 | No |
| rs12035927 | 1 | 97254014 | 4,690 | 3,141 | -3,317 | 3,316 | 1,063E-05 | 0,0002213 | 0,000 | 0,007 | 0,000 | 0,001 | No |
| rs12420205 | 11 | 113394035 | 7,485 | -3,124 | -5,270 | -5,316 | 3,604E-13 | 7,772E-10 | 0,000 | 0,000 | 0,000 | 0,002 | No |
| rs12522815 | 5 | 108718286 | 4,100 | 3,126 | 2,911 | -2,888 | 0,0001182 | 0,001306 | 0,000 | 0,047 | 0,001 | 0,022 | No |
| rs12523398 | 5 | 45119647 | 4,220 | -3,139 | -2,982 | 2,986 | 7,296E-05 | 0,0008834 | 0,000 | 0,032 | 0,000 | 0,007 | No |
| rs12745861* | 1 | 73415014 | 6,451 | 3,140 | -4,563 | -4,560 | 3,685E-10 | 1,314E-07 | 0,000 | 0,000 | 0,000 | 0,001 | No |
| rs13107325 | 4 | 103188709 | 7,829 | 1,801 | -7,049 | 3,407 | 2,9E-21 | 0,0002041 | 0,000 | 0,000 | 0,002 | 0,037 | No |
| rs132528 | 22 | 29295898 | 4,598 | 3,019 | 3,350 | 3,150 | 3,978E-06 | 0,0002636 | 0,000 | 0,010 | 0,003 | 0,043 | No |
| rs13265006 | 8 | 52790523 | 4,288 | -3,099 | -3,000 | -3,065 | 3,676E-05 | 0,000392 | 0,000 | 0,027 | 0,001 | 0,034 | No |
| rs1353911 | 3 | 117777904 | 5,212 | 3,141 | -3,686 | -3,685 | 3,756E-07 | 2,009E-05 | 0,000 | 0,001 | 0,000 | 0,001 | No |
| rs1394092 | 3 | 136196071 | 4,307 | -3,123 | 3,031 | -3,060 | 5,279E-05 | 0,0006536 | 0,000 | 0,026 | 0,001 | 0,022 | No |
| rs1396724 | 2 | 22668907 | 4,799 | 3,129 | 3,404 | 3,383 | 2,514E-06 | 0,0000903 | 0,000 | 0,005 | 0,000 | 0,012 | No |
| rs1405238 | 14 | 99733954 | 5,021 | 2,903 | -3,756 | -3,332 | 2,328E-07 | 0,000111 | 0,000 | 0,002 | 0,004 | 0,048 | No |
| rs1426347 | 12 | 108583598 | 4,460 | 3,110 | 3,178 | -3,129 | 2,481E-05 | 0,0004963 | 0,000 | 0,016 | 0,001 | 0,027 | No |
| rs150299 | 15 | 89943337 | 4,832 | -3,109 | 3,388 | -3,445 | 7,458E-06 | 0,0001244 | 0,000 | 0,004 | 0,001 | 0,021 | No |
| rs152596 | 5 | 106781232 | 4,903 | 3,020 | -3,571 | 3,360 | 2,347E-06 | 0,0001864 | 0,000 | 0,003 | 0,002 | 0,037 | No |
| rs1526800 | 12 | 75313389 | 4,353 | 3,074 | -3,129 | -3,025 | 1,596E-05 | 0,0004603 | 0,000 | 0,022 | 0,002 | 0,037 | No |
| rs1558957 | 2 | 162890175 | 5,103 | -3,116 | 3,585 | 3,631 | 7,644E-07 | 2,663E-05 | 0,000 | 0,002 | 0,000 | 0,016 | No |
| rs161346 | 7 | 137048159 | 6,499 | 2,573 | -5,200 | 3,899 | 4,917E-12 | 1,613E-05 | 0,000 | 0,000 | 0,002 | 0,035 | No |
| rs16960778 | 18 | 26266275 | 4,308 | -3,026 | 2,957 | 3,133 | 4,803E-05 | 0,000292 | 0,000 | 0,026 | 0,004 | 0,048 | No |
| rs17479770 | 2 | 225335814 | 4,563 | 3,092 | 3,266 | 3,186 | 6,466E-06 | 0,0002251 | 0,000 | 0,011 | 0,001 | 0,032 | No |
| rs17503400 | 7 | 71706809 | 4,275 | 3,043 | 3,096 | 2,948 | 1,793E-05 | 0,0006398 | 0,000 | 0,028 | 0,003 | 0,046 | No |
| rs1806555 | 3 | 17016794 | 4,135 | 3,126 | -2,935 | -2,913 | 5,147E-05 | 0,0007477 | 0,000 | 0,042 | 0,001 | 0,021 | No |
| rs1881502 | 11 | 1507512 | 4,304 | -3,037 | 2,963 | -3,122 | 0,0000812 | 0,0005017 | 0,000 | 0,026 | 0,003 | 0,046 | No |
| rs1881835 | 15 | 93490189 | 4,286 | -3,126 | 3,019 | 3,042 | 3,314E-05 | 0,0004308 | 0,000 | 0,027 | 0,001 | 0,019 | No |
| rs1933873 | 6 | 120989799 | 4,101 | 3,057 | 2,961 | -2,837 | 8,077E-05 | 0,001593 | 0,000 | 0,047 | 0,003 | 0,046 | No |
| rs2033804 | 2 | 201222403 | 5,922 | 2,966 | -4,368 | -3,999 | 2,155E-09 | 3,549E-06 | 0,000 | 0,000 | 0,001 | 0,026 | No |
| rs2141233 | 7 | 115065555 | 4,464 | -3,127 | 3,145 | 3,168 | 1,528E-05 | 0,0002478 | 0,000 | 0,016 | 0,000 | 0,016 | No |

| | | | | | | | | | | | | | |
|---|---|---|---|---|---|---|---|---|---|---|---|---|---|
| rs2155286 | 11 | 112847142 | 4,182 | -3,112 | -2,935 | -2,979 | 5,439E-05 | 0,0005697 | 0,000 | 0,036 | 0,001 | 0,030 | No |
| rs2165212 | 2 | 175017480 | 4,237 | 3,140 | -2,997 | 2,995 | 7,623E-05 | 0,0008527 | 0,000 | 0,031 | 0,000 | 0,005 | Yes |
| rs2183573 | 21 | 40574305 | 4,327 | -3,128 | -3,049 | 3,070 | 4,755E-05 | 0,0006283 | 0,000 | 0,024 | 0,000 | 0,016 | No |
| rs2271920 | 8 | 27316117 | 4,104 | 3,131 | -2,910 | -2,894 | 5,724E-05 | 0,0008115 | 0,000 | 0,046 | 0,000 | 0,017 | No |
| rs2350976 | 17 | 17708529 | 4,943 | 3,138 | 3,498 | 3,492 | 1,234E-06 | 5,329E-05 | 0,000 | 0,003 | 0,000 | 0,006 | No |
| rs2639220 | 3 | 81034568 | 5,450 | -2,703 | 3,409 | -4,253 | 0,0000076 | 1,995E-06 | 0,000 | 0,000 | 0,004 | 0,050 | No |
| rs2648523 | 3 | 9412338 | 4,135 | 3,135 | -2,929 | 2,919 | 0,0001043 | 0,001154 | 0,000 | 0,042 | 0,000 | 0,013 | No |
| rs2710331 | 3 | 52837855 | 6,341 | 3,123 | 4,505 | -4,462 | 2,63E-09 | 6,78E-07 | 0,000 | 0,000 | 0,000 | 0,004 | No |
| rs2782638 | 1 | 43880124 | 4,730 | -3,139 | 3,343 | 3,347 | 4,423E-06 | 0,0001073 | 0,000 | 0,006 | 0,000 | 0,005 | No |
| rs2844508 | 6 | 31436499 | 4,118 | 3,135 | 2,916 | -2,907 | 0,000112 | 0,00121 | 0,000 | 0,044 | 0,000 | 0,013 | No |
| rs28483843 | 14 | 104193439 | 4,949 | 3,094 | -3,541 | -3,458 | 1,113E-06 | 6,246E-05 | 0,000 | 0,003 | 0,001 | 0,025 | No |
| rs28661259 | 15 | 77915301 | 4,141 | -3,119 | -2,912 | -2,945 | 6,046E-05 | 0,0006567 | 0,000 | 0,041 | 0,001 | 0,027 | No |
| rs28676999 | 15 | 40569884 | 5,855 | 2,503 | -4,745 | 3,429 | 2,658E-10 | 0,0001505 | 0,000 | 0,000 | 0,004 | 0,047 | No |
| rs2945997 | 11 | 45977339 | 4,880 | -2,985 | 3,313 | -3,583 | 0,0000122 | 6,429E-05 | 0,000 | 0,004 | 0,003 | 0,041 | No |
| rs2954558 | 7 | 24736636 | 5,280 | -3,118 | -3,711 | 3,756 | 8,413E-07 | 2,873E-05 | 0,000 | 0,001 | 0,000 | 0,010 | No |
| rs3094672 | 6 | 30993377 | 4,356 | -3,035 | 2,997 | 3,161 | 3,806E-05 | 0,0002583 | 0,000 | 0,022 | 0,003 | 0,046 | No |
| rs34430 | 5 | 107466006 | 4,093 | -3,052 | -2,829 | 2,958 | 0,0001711 | 0,0009758 | 0,000 | 0,048 | 0,004 | 0,047 | No |
| rs35103010 | 2 | 76405127 | 4,436 | 3,038 | -3,217 | 3,055 | 0,0000181 | 0,0006785 | 0,000 | 0,017 | 0,003 | 0,043 | No |
| rs35427970 | 17 | 7825945 | 4,200 | -3,067 | 2,914 | 3,025 | 0,0000605 | 0,0004684 | 0,000 | 0,035 | 0,003 | 0,041 | Yes |
| rs35431433 | 8 | 9660621 | 4,098 | 3,124 | 2,910 | 2,885 | 5,646E-05 | 0,0008399 | 0,000 | 0,047 | 0,001 | 0,023 | No |
| rs3897855 | 1 | 97918137 | 4,884 | -2,992 | 3,322 | 3,580 | 4,474E-06 | 3,512E-05 | 0,000 | 0,004 | 0,003 | 0,041 | No |
| rs4341564 | 11 | 83231605 | 4,584 | 3,126 | 3,254 | -3,229 | 1,482E-05 | 0,0003245 | 0,000 | 0,011 | 0,000 | 0,015 | No |
| rs4374510 | 3 | 16790756 | 4,566 | -3,139 | 3,227 | 3,230 | 9,392E-06 | 0,0001857 | 0,000 | 0,011 | 0,000 | 0,006 | No |
| rs4422110 | 2 | 146114898 | 4,515 | -3,023 | -3,097 | 3,286 | 4,288E-05 | 0,0002497 | 0,000 | 0,013 | 0,003 | 0,045 | No |
| rs4478795 | 1 | 243844576 | 4,261 | -3,135 | 3,008 | 3,018 | 3,298E-05 | 0,0004798 | 0,000 | 0,029 | 0,000 | 0,011 | No |
| rs4790351 | 17 | 1296092 | 4,167 | 3,122 | 2,961 | -2,932 | 8,962E-05 | 0,0011 | 0,000 | 0,038 | 0,001 | 0,025 | No |
| rs4822074 | 22 | 42362457 | 5,496 | -2,815 | -3,557 | 4,190 | 2,449E-06 | 2,868E-06 | 0,000 | 0,000 | 0,003 | 0,043 | No |
| rs4908761 | 1 | 8552219 | 4,571 | -3,084 | -3,185 | -3,278 | 1,146E-05 | 0,0001499 | 0,000 | 0,011 | 0,001 | 0,034 | Yes |
| rs4994764 | 7 | 1928662 | 5,298 | 3,139 | 3,749 | 3,744 | 2,326E-07 | 0,0000148 | 0,000 | 0,001 | 0,000 | 0,003 | No |
| rs548692 | 6 | 84235749 | 5,108 | 3,077 | 3,670 | -3,553 | 1,239E-06 | 7,694E-05 | 0,000 | 0,001 | 0,001 | 0,029 | No |
| rs56218837 | 5 | 101799787 | 4,353 | -3,139 | 3,077 | 3,080 | 2,113E-05 | 0,0003655 | 0,000 | 0,022 | 0,000 | 0,006 | No |
| rs576982 | 15 | 78870803 | 7,969 | 3,107 | 5,684 | 5,586 | 5,43E-15 | 1,005E-10 | 0,000 | 0,000 | 0,000 | 0,001 | No |
| rs596379 | 15 | 59052072 | 4,405 | 3,122 | -3,130 | -3,099 | 1,454E-05 | 0,0003347 | 0,000 | 0,019 | 0,001 | 0,021 | No |
| rs603542 | 1 | 44079411 | 6,976 | 3,086 | 5,001 | 4,865 | 5,116E-12 | 1,779E-08 | 0,000 | 0,000 | 0,000 | 0,006 | No |
| rs615315 | 1 | 35852521 | 4,304 | -3,073 | 2,991 | -3,095 | 7,052E-05 | 0,0005618 | 0,000 | 0,026 | 0,002 | 0,039 | No |
| rs62141882 | 2 | 58495006 | 4,742 | 2,990 | 3,477 | 3,224 | 1,618E-06 | 0,000187 | 0,000 | 0,006 | 0,003 | 0,044 | No |
| rs62519821 | 8 | 65464495 | 4,293 | -3,066 | 2,978 | -3,092 | 8,371E-05 | 0,0005693 | 0,000 | 0,027 | 0,002 | 0,040 | No |
| rs6457541 | 6 | 32285348 | 5,398 | -3,062 | 3,740 | -3,892 | 6,608E-07 | 1,443E-05 | 0,000 | 0,000 | 0,001 | 0,023 | No |
| rs6485721 | 11 | 47040854 | 4,305 | -3,141 | 3,043 | 3,045 | 2,892E-05 | 0,0004269 | 0,000 | 0,026 | 0,000 | 0,004 | No |
| rs6602338 | 10 | 8837965 | 4,562 | 3,140 | 3,227 | 3,224 | 8,126E-06 | 0,0001907 | 0,000 | 0,011 | 0,000 | 0,005 | No |
| rs6716047 | 2 | 60579769 | 4,097 | -3,047 | 2,828 | 2,964 | 9,026E-05 | 0,0006108 | 0,000 | 0,047 | 0,004 | 0,048 | No |
| rs6818974 | 4 | 31154580 | 4,438 | -3,047 | 3,063 | 3,212 | 2,176E-05 | 0,0002051 | 0,000 | 0,017 | 0,003 | 0,041 | No |
| rs6893642 | 5 | 60117723 | 5,513 | -2,771 | 3,520 | 4,242 | 1,135E-06 | 9,95E-07 | 0,000 | 0,000 | 0,003 | 0,046 | No |
| rs6897407 | 5 | 137798795 | 4,501 | 3,067 | -3,242 | -3,123 | 7,331E-06 | 0,0002991 | 0,000 | 0,014 | 0,002 | 0,037 | No |
| rs702915 | 2 | 65770596 | 4,206 | 3,111 | -2,997 | 2,952 | 7,191E-05 | 0,001016 | 0,000 | 0,034 | 0,001 | 0,031 | No |
| rs7042967 | 9 | 134856295 | 4,996 | -2,946 | 3,356 | 3,701 | 3,35E-06 | 0,0000191 | 0,000 | 0,002 | 0,003 | 0,044 | No |

| | | | | | | | | | | | | | |
|---|---|---|---|---|---|---|---|---|---|---|---|---|---|
| rs7119079 | 11 | 24365908 | 4,147 | 3,122 | -2,947 | 2,917 | 8,658E-05 | 0,001162 | 0,000 | 0,041 | 0,001 | 0,025 | No |
| rs7238071 | 18 | 77579812 | 6,085 | 2,801 | -4,653 | -3,922 | 1,744E-10 | 5,262E-06 | 0,000 | 0,000 | 0,002 | 0,036 | No |
| rs7263949 | 20 | 43690561 | 4,948 | -3,117 | 3,478 | -3,520 | 4,454E-06 | 8,809E-05 | 0,000 | 0,003 | 0,000 | 0,016 | No |
| rs73165497 | 7 | 140684421 | 4,187 | 3,088 | -3,000 | -2,920 | 3,767E-05 | 0,0007225 | 0,000 | 0,036 | 0,002 | 0,037 | No |
| rs73169123 | 22 | 42611834 | 5,444 | 3,124 | 3,866 | 3,833 | 1,068E-07 | 9,168E-06 | 0,000 | 0,000 | 0,000 | 0,008 | No |
| rs7540637 | 1 | 163730212 | 4,925 | -2,903 | 3,269 | -3,685 | 1,712E-05 | 3,924E-05 | 0,000 | 0,003 | 0,004 | 0,049 | No |
| rs7612158 | 3 | 24109112 | 4,121 | -3,093 | -2,879 | -2,949 | 7,651E-05 | 0,0006484 | 0,000 | 0,044 | 0,002 | 0,037 | No |
| rs7683507 | 4 | 33921627 | 4,200 | 3,134 | -2,976 | 2,964 | 7,923E-05 | 0,000967 | 0,000 | 0,035 | 0,000 | 0,014 | No |
| rs7692210 | 4 | 30833204 | 4,261 | 3,038 | 3,090 | 2,934 | 2,091E-05 | 0,0006752 | 0,000 | 0,029 | 0,004 | 0,047 | No |
| rs7745675 | 6 | 83784072 | 4,200 | 3,063 | 3,028 | -2,911 | 5,353E-05 | 0,0012 | 0,000 | 0,035 | 0,003 | 0,042 | No |
| rs77770606 | 2 | 162861575 | 4,159 | 3,069 | 2,994 | 2,887 | 3,821E-05 | 0,0008294 | 0,000 | 0,039 | 0,003 | 0,042 | No |
| rs78406870 | 1 | 44011061 | 4,305 | 3,141 | -3,045 | -3,044 | 2,631E-05 | 0,0004282 | 0,000 | 0,026 | 0,000 | 0,003 | Yes |
| rs7931440 | 11 | 28623873 | 4,997 | 3,099 | -3,571 | -3,495 | 9,885E-07 | 5,202E-05 | 0,000 | 0,002 | 0,001 | 0,022 | No |
| rs811041 | 6 | 26226032 | 6,763 | 3,102 | -4,829 | 4,735 | 1,813E-10 | 1,356E-07 | 0,000 | 0,000 | 0,000 | 0,004 | No |
| rs814143 | 6 | 163854303 | 4,963 | -3,128 | 3,497 | 3,522 | 1,479E-06 | 0,0000461 | 0,000 | 0,003 | 0,000 | 0,011 | No |
| rs880446 | 20 | 62133177 | 4,483 | -3,083 | -3,123 | 3,216 | 3,488E-05 | 0,0003397 | 0,000 | 0,015 | 0,002 | 0,035 | No |
| rs916896 | 17 | 44098797 | 5,105 | -2,970 | -3,452 | 3,761 | 4,916E-06 | 2,731E-05 | 0,000 | 0,001 | 0,003 | 0,041 | Yes |
| rs9257566 | 6 | 29144532 | 10,014 | 1,569 | 9,254 | 3,827 | 1,269E-36 | 5,449E-06 | 0,000 | 0,000 | 0,000 | 0,015 | No |
| rs9277769 | 6 | 33098596 | 4,131 | 3,066 | -2,976 | -2,865 | 4,017E-05 | 0,0009068 | 0,000 | 0,043 | 0,003 | 0,043 | No |
| rs9467804 | 6 | 26583129 | 4,919 | 3,133 | -3,485 | -3,471 | 1,52E-06 | 5,892E-05 | 0,000 | 0,003 | 0,000 | 0,008 | No |
| rs9861216 | 3 | 50059339 | 4,477 | -3,140 | -3,164 | 3,166 | 2,779E-05 | 0,0004223 | 0,000 | 0,015 | 0,000 | 0,004 | No |
| rs647478 | 1 | 73920040 | 8,784 | -2,291 | 4,760 | 7,382 | 3,884E-11 | 2,158E-17 | 0,000 | 0,000 | 0,000 | 0,007 | No |
| rs4887077 | 15 | 78978364 | 4,445 | -3,128 | -3,132 | -3,154 | 1,678E-05 | 0,0002629 | 0,000 | 0,017 | 0,000 | 0,016 | No |
| rs7195 | 6 | 32412539 | 4,378 | 3,074 | -3,147 | -3,043 | 1,587E-05 | 0,0004254 | 0,000 | 0,021 | 0,002 | 0,037 | No |
| rs11210320 | 1 | 74090402 | 5,571 | -2,831 | 3,621 | 4,233 | 5,38E-07 | 1,032E-06 | 0,000 | 0,000 | 0,003 | 0,042 | No |

*Loci within statistically significant correlated regions between SCZ and TUD using LAVA

**Table S13. List of coding genes mapped by the risk pleiotropic loci for schizophrenia-substance use disorders and schizophrenia-addiction factor pairs.**

| SCZ-AF | | SCZ-CanUD | | SCZ-OUD | | SCZ-PAU | | SCZ-TUD | |
|---|---|---|---|---|---|---|---|---|---|
| Concordant pleiotropic loci | Discordant pleiotropic loci | Concordant pleiotropic loci | Discordant pleiotropic loci | Concordant pleiotropic loci | Discordant pleiotropic loci | Concordant pleiotropic loci | Discordant pleiotropic loci | Concordant pleiotropic loci | Discordant pleiotropic loci |
| PLCH2 | DLGAP3 | ZBTB48 | PBX1 | TSNARE1 | NA | RERE | DPYD | RERE | ZMYM4 |
| SLC45A1 | RP11-244H3.4 | KLHL21 | BANK1 | | | C1orf210 | DENND2C | CCDC23 | KIAA0319L |
| RERE | ZMYM6 | PHF13 | SLC39A8 | | | TIE1 | NRAS | C1orf210 | NCDN |
| PTPRF | ZMYM1 | THAP3 | EPHA7 | | | MPL | CSDE1 | TIE1 | TFAP2E |
| KDM4A | SFPQ | DNAJC11 | FOXO3 | | | CDC20 | SYCP1 | MPL | PSMB2 |
| ST3GAL3 | TFAP2E | PTPRF | MPP6 | | | ELOVL1 | TSPAN2 | CDC20 | PTBP2 |
| ARTN | NEGR1 | KDM4A | DFNA5 | | | MED8 | RC3H1 | ELOVL1 | PBX1 |
| NEGR1 | DPYD | ST3GAL3 | KCNE3 | | | SZT2 | RABGAP1L | MED8 | OLA1 |
| ZNF644 | C1orf54 | ARTN | IREB2 | | | HYI | MRPS14 | SZT2 | SPATS2L |
| PTBP2 | ADAMTSL4 | IPO13 | HYKK | | | KLF17 | FOXN2 | HYI | SRGAP3 |
| DPYD | ADAMTSL4-AS1 | DPH2 | AC027228.1 | | | DMAP1 | OLA1 | PTPRF | THUMPD3 |
| MAEL | MCL1 | ATP6V0B | PSMA4 | | | ERI3 | FOXP1 | KDM4A | SETD5 |
| TNN | GOLPH3L | B4GALT2 | CHRNA5 | | | PDE4B | PIK3CA | ST3GAL3 | RNF123 |
| KIAA0040 | HORMAD1 | SLC6A9 | CHRNA3 | | | DPYD | KCNMB3 | ARTN | GMPPB |
| OTOF | CTSS | BARHL2 | CHRNB4 | | | SLC9C2 | PPP3CA | DPYD | FAM212A |
| DPYSL5 | CTSK | SDCCAG8 | ADAMTS7 | | | ANKRD45 | BANK1 | EXTL2 | TRAIP |
| MAPRE3 | ARNT | AKT3 | FMNL1 | | | KLHL20 | SLC39A8 | SDCCAG8 | CAMKV |
| TMEM214 | ANXA9 | ZNF804A | ARHGAP27 | | | CENPL | HIST1H2BD | AKT3 | MST1R |
| AGBL5 | FAM63A | PLCL2 | PLEKHM1 | | | DARS2 | HIST1H2BE | VRK2 | CTD-2330K9.3 |
| OST4 | PRUNE | BBX | CRHR1 | | | ZBTB37 | HIST1H4D | FANCL | MON1A |
| EMILIN1 | BNIPL | NDST3 | MAPT | | | SERPINC1 | HIST1H3D | SLC4A10 | RBM6 |
| KHK | C1orf56 | INPP4B | STH | | | RC3H1 | HIST1H2AD | DPP4 | RBM5 |
| CGREF1 | CDC42SE1 | FREM3 | KANSL1 | | | RABGAP1L | HIST1H2BF | FTCDNL1 | SEMA3F |
| ABHD1 | MLLT11 | DEPDC1B | ARL17B | | | GALNT2 | HIST1H4E | SPATS2L | HYAL3 |
| PREB | GABPB2 | ERCC8 | LRRC37A | | | ARV1 | HIST1H2BG | CUL3 | NAT6 |
| C2orf53 | SEMA6C | NDUFAF2 | LRRC37A2 | | | OTOF | HIST1H2AE | PLCL2 | RASSF1 |
| IFT172 | TNFAIP8L2 | SMIM15 | ARL17A | | | DPYSL5 | HIST1H3E | THRB | GPR62 |
| VRK2 | SCNM1 | HIST1H2BA | WNT3 | | | MAPRE3 | HIST1H1D | LSAMP | DUSP7 |
| FANCL | LYSMD1 | HIST1H4A | LRRC37A3 | | | TMEM214 | HIST1H3G | PCDH7 | POC1A |
| FBXO41 | TMOD4 | HIST1H3E | C17orf58 | | | AGBL5 | HIST1H2BI | DEPDC1B | PPM1M |
| ALMS1 | VPS72 | BTN3A2 | NCAN | | | OST4 | HIST1H4H | ELOVL7 | GLYCTK |
| NAT8 | LINGO4 | BTN2A2 | HAPLN4 | | | EMILIN1 | BTN3A2 | ERCC8 | SEMA3G |
| TPRKB | KLHL29 | BTN3A1 | TM6SF2 | | | KHK | PRSS16 | SLCO4C1 | NISCH |
| LMAN2L | ATAD2B | BTN3A3 | SUGP1 | | | CGREF1 | EYS | SLCO6A1 | STAB1 |
| CNNM4 | UBXN2A | BTN2A1 | MAU2 | | | ABHD1 | BAI3 | PAM | NT5DC2 |
| FAHD2B | MFSD2B | BTN1A1 | GATAD2A | | | PREB | DOPEY1 | REEP2 | SMIM4 |
| ANKRD36 | C2orf44 | HMGN4 | TSSK6 | | | C2orf53 | AL139333.1 | EGR1 | PBRM1 |
| CNTNAP5 | FAM228B | ABT1 | NDUFA13 | | | IFT172 | PGM3 | ETF1 | GNL3 |
| DYNC1I2 | FAM228A | ZNF322 | YJEFN3 | | | VRK2 | RWDD2A | HIST1H4A | GLT8D1 |
| AC068039.1 | TCF7L1 | HIST1H2BJ | CTC-260F20.3 | | | FANCL | ME1 | BTN2A2 | SPCS1 |
| SLC25A12 | RETSAT | HIST1H2AG | CILP2 | | | FBXO41 | PRSS35 | BTN3A3 | NEK4 |
| HAT1 | ELMOD3 | HIST1H2BK | PBX4 | | | ALMS1 | SDK1 | BTN2A1 | ITIH1 |
| METAP1D | CAPG | PRSS16 | LPAR2 | | | NAT8 | PRDM14 | BTN1A1 | ITIH3 |
| SLC39A10 | SH2D6 | POM121L2 | GMIP | | | TPRKB | C8orf87 | HMGN4 | ITIH4 |
| FTCDNL1 | MAT2A | ZNF391 | ATP13A1 | | | SLC4A10 | AC016885.1 | ABT1 | RP5-966M1.6 |
| C2orf69 | ARHGAP15 | ZNF184 | ZNF101 | | | DPP4 | LUZP2 | ZNF322 | SFMBT1 |
| TYW5 | GTDC1 | HIST1H2BL | ZNF536 | | | ZNF804A | SNX19 | HIST1H2BK | RFT1 |
| C2orf47 | FAM124B | HIST1H2AI | ITCH | | | FTCDNL1 | RLBP1 | PRSS16 | PPP2R3A |
| SPATS2L | CUL3 | HIST1H3H | DYNLRB1 | | | C2orf69 | FANCI | ZNF391 | MSL2 |
| PARD3B | FRMD4B | HIST1H2AJ | MAP1LC3A | | | TYW5 | POLG | HIST1H2BL | PCCB |
| CUL3 | CDS1 | HIST1H2BM | PIGU | | | C2orf47 | DPH1 | HIST1H2AI | STAG1 |
| GPC1 | WDFY3 | HIST1H4J | NCOA6 | | | SPATS2L | OVCA2 | HIST1H3H | CDS1 |
| CNTN4 | BANK1 | HIST1H4K | TP53INP2 | | | CREB1 | HIC1 | HIST1H2AJ | WDFY3 |
| PLCL2 | SLC39A8 | HIST1H2AK | ACSS2 | | | METTL21A | SMG6 | HIST1H2BM | EFNA5 |
| THRB | UGT8 | HIST1H2BN | GSS | | | ERBB4 | BCL2 | HIST1H4J | FBXL17 |
| FOXP1 | HCN1 | HIST1H2AL | MYH7B | | | CUL3 | KDSR | HIST1H4K | FER |
| BBX | RGMB | HIST1H1B | EDEM2 | | | SPHKAP | PSMG1 | HIST1H2AK | PJA2 |
| IFT57 | CHD1 | HIST1H3I | | | | DAZL | BRWD1 | HIST1H2BN | HIST1H2AC |
| OTOL1 | MAN2A1 | HIST1H4L | | | | PLCL2 | HMGN1 | HIST1H3I | HIST1H1E |
| NDUFB5 | AC011366.3 | HIST1H3J | | | | BBX | EP300 | HIST1H4L | HIST1H2BD |
| PEX5L | MFAP3 | HIST1H2AM | | | | SLC51A | L3MBTL2 | HIST1H3J | HIST1H2BE |
| TTC14 | GALNT10 | HIST1H2BO | | | | PCYT1A | CHADL | HIST1H2AM | HIST1H4D |
| CCDC39 | HIST1H1E | OR2B2 | | | | RP11-447L10.1 | RANGAP1 | HIST1H2BO | HIST1H3D |
| SOX2 | HIST1H2BD | OR2B6 | | | | TCTEX1D2 | AL035681.1 | ZKSCAN8 | HIST1H2AD |
| NDST3 | HIST1H2BE | ZNF165 | | | | TM4SF19 | ZC3H7B | ZSCAN9 | HIST1H2BF |
| PLK2 | HIST1H4D | ZSCAN16 | | | | TM4SF19 | TEF | ZSCAN31 | HIST1H4E |
| GAPT | HIST1H3D | ZKSCAN8 | | | | UBXN7 | TOB2 | ZKSCAN3 | HIST1H2BG |
| CTD-2117L12.1 | HIST1H2AD | ZSCAN9 | | | | PCDH7 | PHF5A | ZSCAN23 | HIST1H2AE |
| RAB3C | HIST1H2BF | ZKSCAN4 | | | | HNRNPD | ACO2 | QKI | HIST1H3E |
| DEPDC1B | HIST1H4E | NKAPL | | | | FREM3 | POLR3H | MAD1L1 | HIST1H1D |
| ERCC8 | HIST1H2BG | PGBD1 | | | | DEPDC1B | CSDC2 | AC110781.3 | HIST1H4F |
| KIAA0825 | HIST1H2AE | ZSCAN31 | | | | ELOVL7 | XRCC6 | CALN1 | HIST1H4G |
| SLCO4C1 | HIST1H3E | ZSCAN23 | | | | ERCC8 | MEI1 | NDUFB2 | HIST1H3F |
| SLCO6A1 | HIST1H1D | KIFC1 | | | | FAM114A2 | TNFRSF13C | BRAF | HIST1H2BH |
| PAM | HIST1H4F | PHF1 | | | | MFAP3 | CENPM | TMEM178B | HIST1H3G |
| FBXL17 | HIST1H4G | CUTA | | | | GALNT10 | NAGA | TAS2R5 | BTN3A2 |
| FER | HIST1H3G | SYNGAP1 | | | | SCGN | CYP2D6 | TNKS | ABT1 |
| BTN3A2 | HIST1H2BI | ZBTB9 | | | | HIST1H2BA | | MSRA | ZNF322 |
| BTN2A2 | HIST1H4H | BAK1 | | | | SLC17A4 | | PRSS55 | UBE3D |
| BTN2A1 | BTN3A2 | KMT2E | | | | SLC17A1 | | PTK2B | DOPEY1 |
| ZNF322 | BTN2A2 | SRPK2 | | | | SLC17A3 | | CHRNA2 | AL139333.1 |
| PRSS16 | BTN2A1 | CHRNA2 | | | | SLC17A2 | | EPHX2 | PGM3 |
| ZNF391 | BTN1A1 | EPHX2 | | | | TRIM38 | | CLU | RWDD2A |
| ZNF184 | HMGN4 | CLU | | | | HIST1H1A | | CCDC25 | PRSS35 |
| HIST1H2BL | ABT1 | CCDC25 | | | | HIST1H3A | | PXDNL | SNAP91 |
| HIST1H2AI | ZNF322 | MMP16 | | | | HIST1H4A | | PCMTD1 | MAD1L1 |
| HIST1H3H | PKIB | CLTA | | | | HIST1H4B | | AC090186.1 | AC110781.3 |
| HIST1H2AJ | MAD1L1 | CORO2A | | | | HIST1H3B | | MED27 | FTSJ2 |
| HIST1H2BM | AC110781.3 | TBC1D2 | | | | HIST1H2AB | | CDH23 | MPP6 |
| HIST1H4J | FTSJ2 | GABBR2 | | | | HIST1H2BB | | SORCS3 | DFNA5 |
| HIST1H4K | SDK1 | IKBKAP | | | | HIST1H3C | | ARHGAP1 | DGKI |
| HIST1H2AK | MPP6 | FAM206A | | | | HFE | | ZNF408 | BHLHE22 |
| HIST1H2BN | DFNA5 | CTNNAL1 | | | | HIST1H2BC | | F2 | CYP7B1 |
| HIST1H3I | RUSC2 | FRRS1L | | | | HIST1H2AC | | CKAP5 | DCAF12 |
| HIST1H4L | FAM166B | MED27 | | | | HIST1H2BD | | LRP4 | UBAP1 |
| HIST1H3J | TESK1 | SORCS3 | | | | HIST1H2BE | | C11orf49 | CHID1 |
| HIST1H2AM | CD72 | METTL15 | | | | HIST1H4D | | ARFGAP2 | BRSK2 |
| HIST1H2BO | CACNB2 | OR5AK2 | | | | HIST1H3D | | PACSIN3 | MOB2 |
| OR2B2 | NSUN6 | SLC43A1 | | | | HIST1H2AD | | DDB2 | DUSP8 |
| ZNF165 | RP11-139J15.7 | TIMM10 | | | | HIST1H2BF | | ACP2 | KRTAP5-1 |

ZSCAN16
ZKSCAN8
ZSCAN9
ZKSCAN4
PGBD1
ZSCAN31
ZKSCAN3
ZSCAN12
ZSCAN23
SCAND3
C6orf100
OR5V1
OR12D3
OR11A1
VPS52
RPS18
B3GALT4
WDR46
PFDN6
RGL2
TAPBP
ZBTB22
DAXX
KIFC1
PHF1
CUTA
SYNGAP1
ZBTB9
BACH2
CDK19
AMD1
GTF3C6
RPF2
SLC16A10
KIAA1919
REV3L
TRAF3IP2
SMLR1
EPB41L2
ZDHHC14
QKI
NPC1L1
DDX56
TMED4
OGDH
CALN1
KMT2E
SRPK2
PPP1R3A
UBN2
TNKS
CA8
NCALD
COL22A1
TSNARE1
RP11-295D22.1
CACNB2
KCNMA1
BTRC
CREB3L1
DGKZ
MDK
CHRM4
AMBRA1
HARBI1
ATG13
ARHGAP1
ZNF408
F2
PACSIN3
MADD
PTPMT1
C1QTNF4
OR4B1
OR4X2
OR4A47
TRIM64C
FOLH1
OR4C13
OR5AK2
SLC43A1
TIMM10
SERPING1
YPEL4
CLP1
ZDHHC5
MED19
TMX2
TMX2-CTNND1
C11orf31
RP11-691N7.6
BTBD18
CTNND1
OR5B17
CNTF
KCTD14
NDUFC2-KCTD14
THRSP
NDUFC2
ALG8
KCTD21
USP35
GAB2
GRM5
DRD2

ARL5B
ANO9
PTDSS2
RNH1
HRAS
LRRC56
C11orf35
TMEM80
EFCAB4A
DSCAML1
MYO1H
KCTD10
UBE3B
MMAB
MVK
SETDB2
RCBTB1
ARL11
EBPL
NPAS3
RLBP1
FANCI
POLG
ELMO3
TPPP3
RLTPR
ACD
PARD6A
ENKD1
C16orf86
GFOD2
RANBP10
TSNAXIP1
CENPT
THAP11
NUTF2
EDC4
NRN1L
PSKH1
CTRL
CTC-479C5.12
PSMB10
LCAT
SLC12A4
DPEP3
DPEP2
DUS2
DDX28
NFATC3
ESRP2
PLA2G15
SLC7A6
PRMT7
DPH1
OVCA2
HIC1
SMG6
EPB41L3
NCAN
HAPLN4
TM6SF2
SUGP1
MAU2
GATAD2A
TSSK6
NDUFA13
YJEFN3
CTC-260F20.3
CILP2
PBX4
LPAR2
ATP13A1
ZNF101
PTPRT
KCNJ6
DSCR4
DSCR8
KCNJ15
MCHR1
SLC25A17
EP300
L3MBTL2
CHADL
RANGAP1
AL035681.1
ZC3H7B
ACO2
POLR3H
CSDC2
MEI1
TNFRSF13C
WBP2NL
NAGA
CYP2D6

SERPING1
YPEL4
CLP1
ZDHHC5
MED19
TMX2
TMX2-CTNND1
C11orf31
RP11-691N7.6
BTBD18
CTNND1
OR5B17
CNTF
NCAM1
DRD2
TMPRSS5
EFTUD1
FAM154B
GOLGA6L9
GOLGA6L18
RPS17L
RP11-152F13.10
RP11-379H8.1
CPEB1
AP3B2
C15orf40
FURIN
FES
CLEC18A
VAC14
HYDIN
CALB2
MARVELD3
AP1G1
IST1
CDH13
DCC
TCF4
KDM4B
FAM83E
NTN5
FUT2
MAMSTR
RASIP1
IZUMO1
FUT1
FGF21
GNL3L

HIST1H4E
HIST1H2BG
HIST1H2AE
HIST1H3E
HIST1H4F
HIST1H3G
HIST1H2BI
BTN3A2
BTN2A2
BTN3A1
BTN3A3
BTN2A1
BTN1A1
HMGN4
ABT1
ZNF322
HIST1H2BJ
HIST1H2AG
HIST1H2BK
PRSS16
POM121L2
ZNF391
ZNF184
HIST1H2AK
ZKSCAN8
ZSCAN9
ZSCAN31
ZKSCAN3
ZSCAN23
C6orf100
ZNF311
OR2J2
OR14J1
UBD
GABBR1
OR2H2
VPS52
RPS18
WDR46
TAPBP
ZBTB22
DAXX
KIFC1
PHF1
CUTA
SYNGAP1
ZBTB9
BAK1
ITPR3
UQCC2
SBP1
IP6K3
LEMD2
MLN
QKI
CALN1
KMT2E
SRPK2
FAM49B
TSNARE1
CORO2A
TBC1D2
GABBR2
SORCS3
RPS13
PIK3C2A
NUCB2
NCR3LG1
KCNJ11
BDNF
METTL15
OR5AK2
SLC43A1
TIMM10
SMTNL1
SERPING1
YPEL4
CLP1
ZDHHC5
MED19
TMX2
TMX2-CTNND1
C11orf31
RP11-691N7.6
BTBD18
CTNND1
OR5B17
MACROD1
NUDT22
TEX40
NCAM1
TTC12
ANKK1
DRD2
TMPRSS5
IGSF9B
CACNA1C
SP7
SP1
PRR13
PCBP2
MAP3K12
RP11-793H13.10
ATF7
ATP5G2

NR1H3
NCAM1
DRD2
TMPRSS5
GLIPR1L2
BCL11B
APOPT1
XRCC3
ZFYVE21
PPP1R13B
TDRD9
ADAM10
FAM63B
LINGO1
CIB2
IREB2
HYKK
AC027228.1
PSMA4
CHRNA5
CHRNA3
CHRNB4
ADAMTS7
CHD2
ACADVL
SLC2A4
YBX2
SPEM1
SLC35G6
TNFSF12
EIF4A1
CHD3
KCNAB3
RP11-1099M24.7
TRAPPC1
CNTROB
GUCY2D
ALOXE3
RAI1
SREBF1
TOM1L2
LRRC48
ATPAF2
GID4
DRG2
MYO15A
ALKBH5
RHBDL3
C17orf75
ZNF207
PSMD11
CDK5R1
MYO1D
HSCB
CCDC117
XBP1
ZNRF3
EP300
CSDC2
DESI1
SREBF2
TNFRSF13C
CENPM
SEPT3
NAGA
SMDT1
NDUFA6
CYP2D6
TCF20
GNL3L

KRTAP5-2
KRTAP5-3
KRTAP5-4
KRTAP5-5
CTD-2210P24.4
PEX16
GYLTL1B
PHF21A
ANKRD42
CCDC90B
DLG2
WSCD2
BUB1B
PAK6
RP11-133K1.2
PLCB2
ANKRD63
KNSTRN
C15orf57
RLBP1
YWHAE
CRK
MYO1C
FMNL1
ARHGAP27
PLEKHM1
CRHR1
SPPL2C
MAPT
STH
KANSL1
ARL17B
LRRC37A
LRRC37A2
ARL17A
WNT3
LRRC37A3
C17orf58
TOMM34
STK4
KCNS1
EEF1A2
PSMG1
BRWD1
HMGN1
EP300
ZC3H7B
TEF
PHF5A
ACO2
POLR3H
CSDC2
NAGA

TMPRSS5
CACNA1C
TMEM117
LRP1
NXPH4
SHMT2
NDUFA4L2
MBD6
XRCC6BP1
ATXN7L3B
GLIPR1L2
TCHP
GIT2
ANKRD13A
C12orf76
IFT81
ATP2A2
ANAPC7
ARPC3
GPN3
FAM216A
VPS29
RAD9B
PPTC7
TCTN1
HVCN1
PPP1CC
CCDC63
BCL7A
LRRC43
B3GNT4
DIABLO
RP11-512M8.5
VPS33A
CLIP1
POSTN
TRPC4
NAA16
RBM26
NDFIP2
NALCN
RASA3
CDC16
UPF3A
BAZ1A
SRP54
FAM177A1
PPP2R3C
KIAA0391
KIAA0391
PSMA6
L2HGDH
RTN1
PPP2R5E
RGS6
AC005477.1
AREL1
AC007956.1
FCF1
YLPM1
PROX2
DLST
RPS6KL1
NEK9
SPATA7
KLC1
APOPT1
RP11-73M18.2
XRCC3
AL049840.1
ZFYVE21
PPP1R13B
TDRD9
KIF26A
FKSG62
SEMA6D
ADAM10
FAM63B
RNF111
HOMER2
FAM103A1
C15orf40
BTBD1
TM6SF1
HDGFRP3
BNC1
FURIN
FES
GRIN2A
BOLA2
SPN
TMEM219
TAOK2
HIRIP3
INO80E
DOC2A
C16orf92
FAM57B
ALDOA
PPP4C
TBX6
YPEL3
GDPD3
MAPK3
NDRG4

GLTP
TCHP
GIT2
ANKRD13A
C12orf76
VPS29
PPP1CC
RBM26
NDFIP2
DNAJC3
UGGT2
HS6ST3
RASA3
CDC16
UPF3A
RTN1
RGS6
AC005477.1
BCL11B
APOPT1
XRCC3
AL049840.1
ZFYVE21
PPP1R13B
KIF26A
FBXO22
ISL2
SCAPER
FURIN
FES
RBFOX1
SPN
ASPHD1
KCTD13
TMEM219
TAOK2
HIRIP3
INO80E
DOC2A
C16orf92
FAM57B
ALDOA
PPP4C
TBX6
YPEL3
GDPD3
MAPK3
CNOT1
ACADVL
SLC2A4
YBX2
SPEM1
SLC35G6
TNFSF12
EIF4A1
CHD3
KCNAB3
RP11-1099M24.7
TRAPPC1
CNTROB
GUCY2D
ALOXE3
FZD2
DBF4B
NMT1
FMNL1
ARHGAP27
PLEKHM1
CRHR1
SPPL2C
MAPT
STH
KANSL1
ARL17B
LRRC37A
LRRC37A2
ARL17A
NSF
WNT3
LRRC37A3
C17orf58
SLC26A11
RPTOR
TCF4
FAM83E
NTN5
FUT2
MAMSTR
RASIP1
IZUMO1
FUT1
FGF21
SRSF6
HSCB
CCDC117
XBP1
ZNRF3
ADSL
MCHR1
SLC25A17
ST13
EP300
TEF
CSDC2
PMM1

*SETD6*
*CNOT1*
*ANKRD11*
*SPG7*
*RPL13*
*CPNE7*
*DPEP1*
*CHMP1A*
*SPATA33*
*CDK10*
*RP11-368I7.4*
*VPS9D1*
*ZNF276*
*FANCA*
*SPIRE2*
*MC1R*
*GAS8*
*C16orf3*
*RHBDL3*
*C17orf75*
*ZNF207*
*PSMD11*
*CDK5R1*
*MYO1D*
*DBF4B*
*NMT1*
*FMNL1*
*ARHGAP27*
*PLEKHM1*
*CRHR1*
*SPPL2C*
*MAPT*
*STH*
*KANSL1*
*ARL17B*
*LRRC37A*
*LRRC37A2*
*ARL17A*
*NSF*
*WNT3*
*TTLL6*
*ATP5G1*
*UBE2Z*
*SNF8*
*GIP*
*IGF2BP1*
*LRRC37A3*
*C17orf58*
*NOL4*
*TCF4*
*PIGN*
*CACNG8*
*CACNG6*
*TFPT*
*PPP1R16B*
*SRSF6*
*KCNB1*
*DYRK1A*
*TANGO2*
*DGCR8*
*TRMT2A*
*RANBP1*
*ZDHHC8*
*AC006547.14*
*HSCB*
*CCDC117*
*XBP1*
*ZNRF3*
*MCHR1*
*EP300*
*L3MBTL2*
*CHADL*
*RANGAP1*
*SREBF2*
*TNFRSF13C*
*CENPM*
*SEPT3*
*WBP2NL*
*NAGA*
*FAM109B*
*SMDT1*
*NDUFA6*
*CYP2D6*
*TCF20*
*CYB5R3*
*A4GALT*
*ARFGAP3*
*PACSIN2*
*TTLL1*
*BRD1*
*ZBED4*
*ALG12*
*CRELD2*
*GNL3L*

*DESI1*
*XRCC6*
*NHP2L1*
*MEI1*
*SREBF2*
*TNFRSF13C*
*CENPM*
*SEPT3*
*NAGA*
*SMDT1*
*NDUFA6*
*CYP2D6*
*TCF20*
*GNL3L*

**Table S14. GO enrichment on biological processes of pleiotropic loci between schizophrenia-substance use disorders and schizophrenia-addiction factor pairs.**

| Enrichment ID | Description | GeneRatio | BgRatio | RichFactor | FoldEnrichment | zScore | pvalue | p.adjust | geneID | Count |
|---|---|---|---|---|---|---|---|---|---|---|
| SCZ - AF | | | | | | | | | | |
| GO:0043543 | protein acylation | 11/462 | 101/18888 | 0,10891089 | 4,45261669 | 5,50893032 | 3,6534E-05 | 0,01769237 | EP300/NAT8/HAT1/ZDHHC14/OGDH/ZDHHC5/NAA16/DLST/NMT1 | 11 |
| GO:0033044 | regulation of chromosome organization | 18/462 | 247/18888 | 0,07287449 | 2,979336453 | 4,9581589 | 3,9938E-05 | 0,01769237 | CUL3/PKIB/MAD1L1/SETDB2/ACD/SMG6/TNKS/ANAPC7/BCL7A/CD | 18 |
| SCZ - CanUD | | | | | | | | | | |
| GO:0048670 | regulation of collateral sprouting | 4/156 | 21/18888 | 0,19047619 | 23,06227106 | 9,23124237 | 2,4017E-05 | 0,01541892 | EPHA7/WNT3/IST1/DCC | 4 |
| GO:0035094 | response to nicotine | 5/156 | 50/18888 | 0,1 | 12,10769231 | 7,17700356 | 5,658E-05 | 0,01816203 | CHRNA5/CHRNA3/CHRNB4/CHRNA2/DRD2 | 5 |
| GO:0007271 | synaptic transmission, cholinergic | 4/156 | 30/18888 | 0,13333333 | 16,14358974 | 7,57518707 | 0,00010379 | 0,02221112 | CHRNA5/CHRNA3/CHRNB4/CHRNA2 | 4 |
| GO:0050804 | modulation of chemical synaptic transmission | 13/156 | 493/18888 | 0,02636917 | 3,192697769 | 4,5019851 | 0,00023347 | 0,03599902 | EPHA7/CHRNA5/CHRNA3/CHRNB4/MAPT/SLC6A9/PLCL2/SYNGAP1 | 13 |
| GO:0061387 | regulation of extent of cell growth | 6/156 | 109/18888 | 0,05504587 | 6,664784757 | 5,41267742 | 0,00028983 | 0,03599902 | EPHA7/MAPT/WNT3/BARHL2/IST1/DCC | 6 |
| GO:0050851 | antigen receptor-mediated signaling pathway | 8/156 | 208/18888 | 0,03846154 | 4,656804734 | 4,83945791 | 0,00034065 | 0,03599902 | BANK1/PLCL2/BTN3A2/BTN2A2/BTN3A1/BTN3A3/BTN2A1/BTN1A | 8 |
| GO:0095500 | acetylcholine receptor signaling pathway | 4/156 | 42/18888 | 0,0952381 | 11,53113553 | 6,23507912 | 0,00039251 | 0,03599902 | CHRNA5/CHRNA3/CHRNB4/CHRNA2 | 4 |
| GO:1905144 | response to acetylcholine | 4/156 | 46/18888 | 0,08695652 | 10,52842809 | 5,90457158 | 0,00055779 | 0,0447625 | CHRNA5/CHRNA3/CHRNB4/CHRNA2 | 4 |
| SCZ - OUD | | | | | | | | | | |
| GO:0048278 | vesicle docking | 1/1 | 64/18888 | 0,015625 | 295,125 | 17,1500729 | 0,00338839 | 0,01162643 | TSNARE1 | 1 |
| GO:0022406 | membrane docking | 1/1 | 92/18888 | 0,01086957 | 205,3043478 | 14,2935072 | 0,00487082 | 0,01162643 | TSNARE1 | 1 |
| GO:0006906 | vesicle fusion | 1/1 | 119/18888 | 0,00840336 | 158,7226891 | 12,5587694 | 0,0063003 | 0,01162643 | TSNARE1 | 1 |
| GO:0048284 | organelle fusion | 1/1 | 159/18888 | 0,00628931 | 118,7924528 | 10,8532232 | 0,00841804 | 0,01162643 | TSNARE1 | 1 |
| GO:0061025 | membrane fusion | 1/1 | 183/18888 | 0,00546448 | 103,2131148 | 10,1100502 | 0,00968869 | 0,01162643 | TSNARE1 | 1 |
| GO:0016050 | vesicle organization | 1/1 | 371/18888 | 0,00269542 | 50,91105121 | 7,06477538 | 0,0196421 | 0,0196421 | TSNARE1 | 1 |
| SCZ - PAU | | | | | | | | | | |
| GO:0015747 | urate transport | 4/295 | 11/18888 | 0,36363636 | 23,2825886 | 9,31136305 | 1,7647E-05 | 0,01058959 | SLC17A4/SLC17A1/SLC17A3/SLC17A2 | 4 |
| GO:0050851 | antigen receptor-mediated signaling pathway | 13/295 | 208/18888 | 0,0625 | 4,001694915 | 5,4831174 | 2,4975E-05 | 0,01058959 | RC3H1/FOXP1/PIK3CA/BANK1/BTN3A2/BCL2/PDE4B/PLCL2/BTN2A | 13 |
| GO:0045026 | plasma membrane fusion | 5/295 | 33/18888 | 0,15151515 | 9,701078582 | 6,30136383 | 0,00014896 | 0,04210464 | TIE1/C16orf92/SPPL2C/NSF/IZUMO1 | 5 |
| GO:0031330 | negative regulation of cellular catabolic process | 12/295 | 224/18888 | 0,05357143 | 3,430024213 | 4,60839833 | 0,00022003 | 0,04664629 | CSDE1/PIK3CA/BCL2/EP300/DAZL/HNRNPD/ZKSCAN3/QKI/UPF3A/ | 12 |
| SCZ - TUD | | | | | | | | | | |
| GO:0045839 | negative regulation of mitotic nuclear division | 6/231 | 56/18888 | 0,10714286 | 8,760667904 | 6,47159699 | 6,109E-05 | 0,02662964 | MAD1L1/BUB1B/CDC20/XRCC3/TOM1L2/ZNF207 | 6 |
| GO:0010965 | regulation of mitotic sister chromatid separation | 6/231 | 59/18888 | 0,10169492 | 8,315210214 | 6,26189474 | 8,2229E-05 | 0,02662964 | MAD1L1/BUB1B/CDC20/CUL3/XRCC3/ZNF207 | 6 |
| GO:0051784 | negative regulation of nuclear division | 6/231 | 63/18888 | 0,0952381 | 7,787260359 | 6,00432049 | 0,00011906 | 0,02662964 | MAD1L1/BUB1B/CDC20/XRCC3/TOM1L2/ZNF207 | 6 |
| GO:0095500 | acetylcholine receptor signaling pathway | 5/231 | 42/18888 | 0,11904762 | 9,734075448 | 6,30519562 | 0,00015412 | 0,02662964 | CHRNA2/CHRNA5/CHRNA3/CHRNB4/CDK5R1 | 5 |
| GO:0071173 | spindle assembly checkpoint signaling | 5/231 | 46/18888 | 0,10869565 | 8,887634105 | 5,95975967 | 0,00023868 | 0,02662964 | MAD1L1/BUB1B/CDC20/XRCC3/ZNF207 | 5 |
| GO:0071174 | mitotic spindle checkpoint signaling | 5/231 | 46/18888 | 0,10869565 | 8,887634105 | 5,95975967 | 0,00023868 | 0,02662964 | MAD1L1/BUB1B/CDC20/XRCC3/ZNF207 | 5 |
| GO:1905144 | response to acetylcholine | 5/231 | 46/18888 | 0,10869565 | 8,887634105 | 5,95975967 | 0,00023868 | 0,02662964 | CHRNA2/CHRNA5/CHRNA3/CHRNB4/CDK5R1 | 5 |
| GO:1905818 | regulation of chromosome separation | 6/231 | 74/18888 | 0,08108108 | 6,62969463 | 5,39916628 | 0,00029032 | 0,02769256 | MAD1L1/BUB1B/CDC20/CUL3/XRCC3/ZNF207 | 6 |
| GO:0035094 | response to nicotine | 5/231 | 50/18888 | 0,1 | 8,176623377 | 5,65398088 | 0,00035458 | 0,02769256 | CHRNA2/DRD2/CHRNA5/CHRNA3/CHRNB4 | 5 |
| GO:0051985 | negative regulation of chromosome segregation | 5/231 | 50/18888 | 0,1 | 8,176623377 | 5,65398088 | 0,00035458 | 0,02769256 | MAD1L1/BUB1B/CDC20/XRCC3/ZNF207 | 5 |
| GO:0007271 | synaptic transmission, cholinergic | 4/231 | 30/18888 | 0,13333333 | 10,9021645 | 6,03961872 | 0,00046529 | 0,03083603 | CHRNA2/CHRNA5/CHRNA3/CHRNB4 | 4 |
| GO:0051304 | chromosome separation | 6/231 | 81/18888 | 0,07407407 | 6,056758057 | 5,07482998 | 0,00047379 | 0,03083603 | MAD1L1/BUB1B/CDC20/CUL3/XRCC3/ZNF207 | 6 |
| GO:0050770 | regulation of axonogenesis | 8/231 | 156/18888 | 0,05128205 | 4,193140193 | 4,45609853 | 0,00068172 | 0,0409557 | SEMA3F/SEMA3G/EFNA5/BRSK2/MAPT/WNT3/BRAF/LRP4 | 8 |
| GO:0030212 | hyaluronan metabolic process | 4/231 | 34/18888 | 0,11764706 | 9,619556914 | 5,59743711 | 0,00075784 | 0,04227644 | HYAL3/ITIH1/ITIH3/ITIH4 | 4 |
| GO:0050807 | regulation of synapse organization | 10/231 | 243/18888 | 0,04115226 | 3,364865587 | 4,12852557 | 0,00084819 | 0,04286475 | SETD5/CAMKV/SEMA3F/EFNA5/CDC20/PTK2B/LRP4/DRD2/ADAM1 | 10 |
| GO:0007626 | locomotory behavior | 9/231 | 202/18888 | 0,04455446 | 3,643050019 | 4,20232338 | 0,00087815 | 0,04286475 | PAK6/SLC4A10/DPP4/EGR1/CDH23/DRD2/CHRNA3/CHRNB4/MYO1 | 9 |
| GO:0050803 | regulation of synapse structure or activity | 10/231 | 249/18888 | 0,04016064 | 3,283784489 | 4,03654746 | 0,0010208 | 0,04317187 | SETD5/CAMKV/SEMA3F/EFNA5/CDC20/PTK2B/LRP4/DRD2/ADAM1 | 10 |
| GO:0006633 | fatty acid biosynthetic process | 8/231 | 166/18888 | 0,04819277 | 3,940541386 | 4,23420416 | 0,00102148 | 0,04317187 | ELOVL1/ELOVL7/QKI/NR1H3/ACADVL/ALOXE3/XBP1/CYP2D6 | 8 |
| GO:0043161 | proteasome-mediated ubiquitin-dependent protein ca | 13/231 | 389/18888 | 0,03341902 | 2,732547658 | 3,84195646 | 0,00105028 | 0,04317187 | PSMB2/FBXL17/PJA2/UBE3D/DCAF12/CDC20/CUL3/ERCC8/CLU/PS | 13 |

**Table S15. List of convergent coding genes across schizophrenia and substance use disorder pairs.**

| SCZ-CanUD, SCZ-PAU | SCZ-CanUD, SCZ-TUD | SCZ-CanUD, SCZ-PAU, SCZ-TUD | SCZ-OUD, SCZ-PAU | SCZ-PAU, SCZ-TUD |
|---|---|---|---|---|
| *BANK1* | *PBX1* | *FMNL1* | *TSNARE1* | *DPYD* |
| *SLC39A8* | *MPP6* | *ARHGAP27* | | *OLA1* |
| *FMNL1* | *DFNA5* | *PLEKHM1* | | *HIST1H2BD* |
| *ARHGAP27* | *IREB2* | *CRHR1* | | *HIST1H2BE* |
| *PLEKHM1* | *HYKK* | *MAPT* | | *HIST1H4D* |
| *CRHR1* | *AC027228.1* | *STH* | | *HIST1H3D* |
| *MAPT* | *PSMA4* | *KANSL1* | | *HIST1H2AD* |
| *STH* | *CHRNA5* | *ARL17B* | | *HIST1H2BF* |
| *KANSL1* | *CHRNA3* | *LRRC37A* | | *HIST1H4E* |
| *ARL17B* | *CHRNB4* | *LRRC37A2* | | *HIST1H2BG* |
| *LRRC37A* | *ADAMTS7* | *ARL17A* | | *HIST1H2AE* |
| *LRRC37A2* | *FMNL1* | *WNT3* | | *HIST1H3E* |
| *ARL17A* | *ARHGAP27* | *LRRC37A3* | | *HIST1H1D* |
| *WNT3* | *PLEKHM1* | *C17orf58* | | *HIST1H3G* |
| *LRRC37A3* | *CRHR1* | *PLCL2* | | *BTN3A2* |
| *C17orf58* | *MAPT* | *DEPDC1B* | | *PRSS16* |
| *ZNF804A* | *STH* | *ERCC8* | | *DOPEY1* |
| *PLCL2* | *KANSL1* | *HIST1H4A* | | *AL139333.1* |
| *BBX* | *ARL17B* | *HIST1H3E* | | *PGM3* |
| *FREM3* | *LRRC37A* | *BTN3A2* | | *RWDD2A* |
| *DEPDC1B* | *LRRC37A2* | *BTN2A2* | | *PRSS35* |
| *ERCC8* | *ARL17A* | *BTN3A3* | | *RLBP1* |
| *HIST1H2BA* | *WNT3* | *BTN2A1* | | *PSMG1* |
| *HIST1H4A* | *LRRC37A3* | *BTN1A1* | | *BRWD1* |
| *HIST1H3E* | *C17orf58* | *HMGN4* | | *HMGN1* |
| *BTN3A2* | *PTPRF* | *ABT1* | | *EP300* |
| *BTN2A2* | *KDM4A* | *ZNF322* | | *ZC3H7B* |
| *BTN3A1* | *ST3GAL3* | *HIST1H2BK* | | *TEF* |
| *BTN3A3* | *ARTN* | *PRSS16* | | *PHF5A* |
| *BTN2A1* | *SDCCAG8* | *ZNF391* | | *ACO2* |
| *BTN1A1* | *AKT3* | *HIST1H2AK* | | *POLR3H* |
| *HMGN4* | *PLCL2* | *ZKSCAN8* | | *CSDC2* |
| *ABT1* | *DEPDC1B* | *ZSCAN9* | | *TNFRSF13C* |
| *ZNF322* | *ERCC8* | *ZSCAN31* | | *CENPM* |
| *HIST1H2BJ* | *HIST1H4A* | *ZSCAN23* | | *NAGA* |
| *HIST1H2AG* | *HIST1H3E* | *SORCS3* | | *CYP2D6* |
| *HIST1H2BK* | *BTN3A2* | *NCAM1* | | *RERE* |
| *PRSS16* | *BTN2A2* | *DRD2* | | *C1orf210* |
| *POM121L2* | *BTN3A3* | *TMPRSS5* | | *TIE1* |
| *ZNF391* | *BTN2A1* | *GNL3L* | | *MPL* |
| *ZNF184* | *BTN1A1* | | | *CDC20* |
| *HIST1H2AK* | *HMGN4* | | | *ELOVL1* |
| *ZKSCAN8* | *ABT1* | | | *MED8* |
| *ZSCAN9* | *ZNF322* | | | *SZT2* |
| *ZSCAN31* | *HIST1H2BK* | | | *HYI* |
| *ZSCAN23* | *PRSS16* | | | *VRK2* |
| *KIFC1* | *ZNF391* | | | *FANCL* |
| *PHF1* | *HIST1H2BL* | | | *SLC4A10* |
| *CUTA* | *HIST1H2AI* | | | *DPP4* |
| *SYNGAP1* | *HIST1H3H* | | | *FTCDNL1* |
| *ZBTB9* | *HIST1H2AJ* | | | *SPATS2L* |
| *BAK1* | *HIST1H2BM* | | | *CUL3* |
| *KMT2E* | *HIST1H4J* | | | *PLCL2* |
| *SRPK2* | *HIST1H4K* | | | *PCDH7* |
| *CORO2A* | *HIST1H2AK* | | | *DEPDC1B* |
| *TBC1D2* | *HIST1H2BN* | | | *ELOVL7* |
| *GABBR2* | *HIST1H3I* | | | *ERCC8* |
| *SORCS3* | *HIST1H4L* | | | *HIST1H4A* |
| *METTL15* | *HIST1H3J* | | | *HIST1H2AC* |
| *OR5AK2* | *HIST1H2AM* | | | *HIST1H4F* |
| *SLC43A1* | *HIST1H2BO* | | | *BTN2A2* |
| *TIMM10* | *ZKSCAN8* | | | *BTN3A3* |

| | | |
|---|---|---|
| *SERPING1* | *ZSCAN9* | *BTN2A1* |
| *YPEL4* | *ZSCAN31* | *BTN1A1* |
| *CLP1* | *ZSCAN23* | *HMGN4* |
| *ZDHHC5* | *CHRNA2* | *ABT1* |
| *MED19* | *EPHX2* | *ZNF322* |
| *TMX2* | *CLU* | *HIST1H2BK* |
| *TMX2-CTNND1* | *CCDC25* | *ZNF391* |
| *C11orf31* | *MED27* | *HIST1H2AK* |
| *RP11-691N7.6* | *SORCS3* | *ZKSCAN8* |
| *BTBD18* | *NCAM1* | *ZSCAN9* |
| *CTNND1* | *DRD2* | *ZSCAN31* |
| *OR5B17* | *TMPRSS5* | *ZKSCAN3* |
| *NCAM1* | *GNL3L* | *ZSCAN23* |
| *DRD2* | | *QKI* |
| *TMPRSS5* | | *CALN1* |
| *FURIN* | | *SORCS3* |
| *FES* | | *NCAM1* |
| *TCF4* | | *DRD2* |
| *FAM83E* | | *TMPRSS5* |
| *NTN5* | | *BCL11B* |
| *FUT2* | | *APOPT1* |
| *MAMSTR* | | *XRCC3* |
| *RASIP1* | | *ZFYVE21* |
| *IZUMO1* | | *PPP1R13B* |
| *FUT1* | | *ACADVL* |
| *FGF21* | | *SLC2A4* |
| *GNL3L* | | *YBX2* |
| | | *SPEM1* |
| | | *SLC35G6* |
| | | *TNFSF12* |
| | | *EIF4A1* |
| | | *CHD3* |
| | | *KCNAB3* |
| | | *RP11-1099M24.7* |
| | | *TRAPPC1* |
| | | *CNTROB* |
| | | *GUCY2D* |
| | | *ALOXE3* |
| | | *FMNL1* |
| | | *ARHGAP27* |
| | | *PLEKHM1* |
| | | *CRHR1* |
| | | *SPPL2C* |
| | | *MAPT* |
| | | *STH* |
| | | *KANSL1* |
| | | *ARL17B* |
| | | *LRRC37A* |
| | | *LRRC37A2* |
| | | *ARL17A* |
| | | *WNT3* |
| | | *LRRC37A3* |
| | | *C17orf58* |
| | | *HSCB* |
| | | *CCDC117* |
| | | *XBP1* |
| | | *ZNRF3* |
| | | *DESI1* |
| | | *SREBF2* |
| | | *SEPT3* |
| | | *SMDT1* |
| | | *NDUFA6* |
| | | *TCF20* |
| | | *GNL3L* |

**Table S16. GO enrichment on biological processes of convergent coding genes across schizophrenia and substance use disorder pairs.**

| Enrichment ID | Description | GeneRatio | BgRatio | RichFactor | FoldEnrichment | zScore | pvalue | p.adjust | geneID | Count |
|---|---|---|---|---|---|---|---|---|---|---|
| SCZ -CanUD, SCZ-PAU | | | | | | | | | | |
| GO:0050851 | antigen receptor-mediated signaling pathway | 8/73 | 208/18888 | 0,03846154 | 9,951527924 | 8,08595297 | 1,3757E-06 | 0,00062733 | BANK1/PLCL2/BTN3A2/BTN2A2/BTN3A1/BTN3A3/BTN2A1/BTN1A | 8 |
| SCZ -CanUD, SCZ-TUD | | | | | | | | | | |
| GO:0035094 | response to nicotine | 5/50 | 50/18888 | 0,1 | 37,776 | 13,414707 | 2,0502E-07 | 8,57E-05 | CHRNA5/CHRNA3/CHRNB4/CHRNA2/DRD2 | 5 |
| GO:0007271 | synaptic transmission, cholinergic | 4/50 | 30/18888 | 0,13333333 | 50,368 | 13,9414338 | 1,1317E-06 | 0,00023652 | CHRNA5/CHRNA3/CHRNB4/CHRNA2 | 4 |
| GO:0095500 | acetylcholine receptor signaling pathway | 4/50 | 42/18888 | 0,0952381 | 35,97714286 | 11,6909143 | 4,5152E-06 | 0,00062912 | CHRNA5/CHRNA3/CHRNB4/CHRNA2 | 4 |
| GO:1905144 | response to acetylcholine | 4/50 | 46/18888 | 0,08695652 | 32,84869565 | 11,1418206 | 6,5317E-06 | 0,00068256 | CHRNA5/CHRNA3/CHRNB4/CHRNA2 | 4 |
| GO:0060079 | excitatory postsynaptic potential | 5/50 | 107/18888 | 0,04672897 | 17,65233645 | 8,89931963 | 9,186E-06 | 0,00076795 | CHRNA5/CHRNA3/CHRNB4/CHRNA2/DRD2 | 5 |
| GO:0050851 | antigen receptor-mediated signaling pathway | 6/50 | 208/18888 | 0,02884615 | 10,89692308 | 7,39422263 | 1,7608E-05 | 0,00122666 | PLCL2/BTN3A2/BTN2A2/BTN3A3/BTN2A1/BTN1A1 | 6 |
| GO:0030534 | adult behavior | 5/50 | 144/18888 | 0,03472222 | 13,11666667 | 7,51938575 | 3,8621E-05 | 0,00230032 | CHRNA5/CHRNA3/CHRNB4/CRHR1/DRD2 | 5 |
| GO:0060078 | regulation of postsynaptic membrane potential | 5/50 | 148/18888 | 0,03378378 | 12,76216216 | 7,40086229 | 4,4025E-05 | 0,00230032 | CHRNA5/CHRNA3/CHRNB4/CHRNA2/DRD2 | 5 |
| GO:0007588 | excretion | 3/50 | 30/18888 | 0,1 | 37,776 | 10,3854758 | 6,7385E-05 | 0,00312964 | CHRNA3/CHRNB4/DRD2 | 3 |
| GO:0050804 | modulation of chemical synaptic transmission | 7/50 | 493/18888 | 0,01419878 | 5,363732252 | 5,05802248 | 0,00029901 | 0,0124988 | CHRNA5/CHRNA3/CHRNB4/MAPT/PLCL2/SORCS3/DRD2 | 7 |
| GO:1901386 | negative regulation of voltage-gated calcium channel a | 2/50 | 15/18888 | 0,13333333 | 50,368 | 9,85416407 | 0,00070541 | 0,02680561 | CRHR1/DRD2 | 2 |
| SCZ -CanUD, SCZ-PAU, SCZ-TUD | | | | | | | | | | |
| GO:0050851 | antigen receptor-mediated signaling pathway | 6/31 | 208/18888 | 0,02884615 | 17,57568238 | 9,7463204 | 9,7099E-07 | 0,00028547 | PLCL2/BTN3A2/BTN2A2/BTN3A3/BTN2A1/BTN1A1 | 6 |
| GO:0050805 | negative regulation of synaptic transmission | 3/31 | 54/18888 | 0,05555556 | 33,84946237 | 9,80121463 | 9,3819E-05 | 0,01379138 | MAPT/SORCS3/DRD2 | 3 |
| GO:1901386 | negative regulation of voltage-gated calcium channel a | 2/31 | 15/18888 | 0,13333333 | 81,23870968 | 12,604776 | 0,00027011 | 0,02647097 | CRHR1/DRD2 | 2 |
| SCZ -OUD, SCZ-PAU | | | | | | | | | | |
| GO:0048278 | vesicle docking | 1/1 | 64/18888 | 0,015625 | 295,125 | 17,1500729 | 0,00338839 | 0,01162643 | TSNARE1 | 1 |
| GO:0022406 | membrane docking | 1/1 | 92/18888 | 0,01086957 | 205,3043478 | 14,2935072 | 0,00487082 | 0,01162643 | TSNARE1 | 1 |
| GO:0006906 | vesicle fusion | 1/1 | 119/18888 | 0,00840336 | 158,7226891 | 12,5587694 | 0,0063003 | 0,01162643 | TSNARE1 | 1 |
| GO:0048284 | organelle fusion | 1/1 | 159/18888 | 0,00628931 | 118,7924528 | 10,8532232 | 0,00841804 | 0,01162643 | TSNARE1 | 1 |
| GO:0061025 | membrane fusion | 1/1 | 183/18888 | 0,00546448 | 103,2131148 | 10,1100502 | 0,00968869 | 0,01162643 | TSNARE1 | 1 |
| GO:0016050 | vesicle organization | 1/1 | 371/18888 | 0,00269542 | 50,91105121 | 7,06477538 | 0,0196421 | 0,0196421 | TSNARE1 | 1 |
| SCZ -PAU, SCZ-TUD | | | | | | | | | | |
| GO:0006633 | fatty acid biosynthetic process | 7/91 | 166/18888 | 0,04216867 | 8,752548656 | 6,98040048 | 1,5541E-05 | 0,00850085 | CYP2D6/ELOVL1/ELOVL7/QKI/ACADVL/ALOXE3/XBP1 | 7 |
