## Supplementary figures and images for "Pleiotropic genetic architecture linking schizophrenia and substance use disorders"

### Figure S1

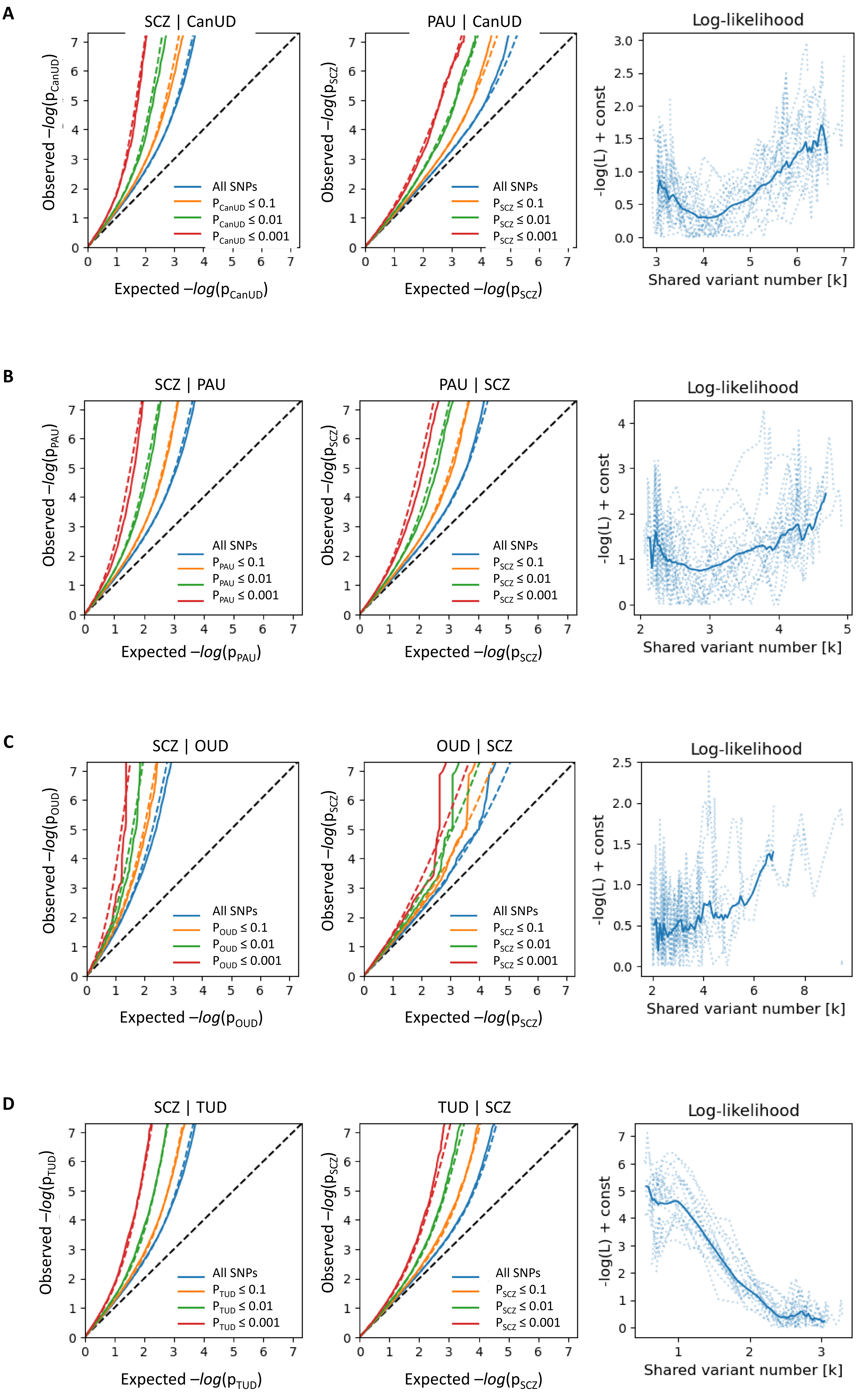

### Figure S2

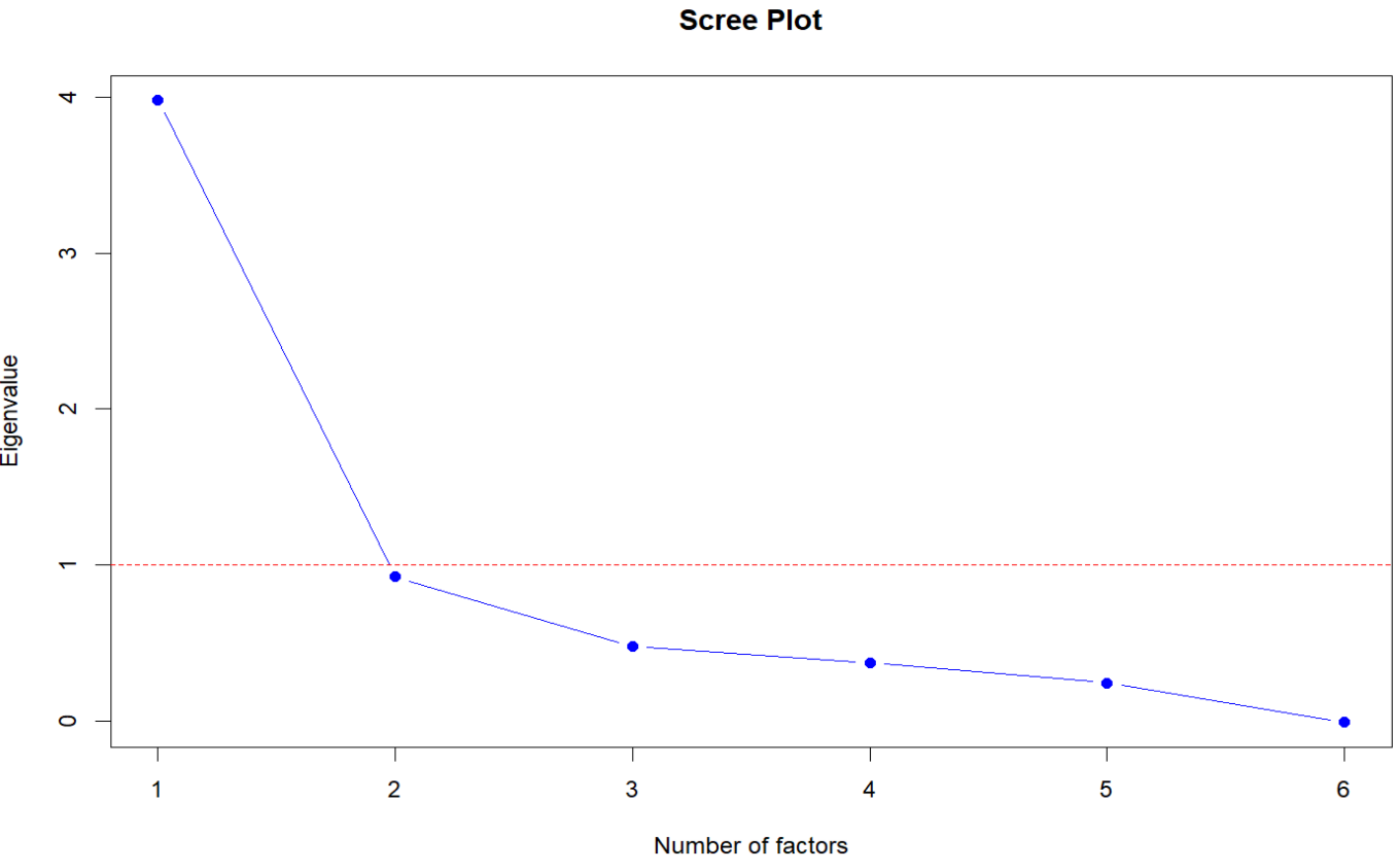
